# A second update on mapping the human genetic architecture of COVID-19

**DOI:** 10.1101/2022.12.24.22283874

**Authors:** The COVID-19 Host Genetics Initiative, Andrea Ganna

**Affiliations:** University of Helsinki

## Abstract

Investigating the role of host genetic factors in COVID-19 severity and susceptibility can inform our understanding of the underlying biological mechanisms that influence adverse outcomes and drug development^1,2^. Here we present a second updated genome-wide association study (GWAS) on COVID-19 severity and infection susceptibility to SARS-CoV-2 from the COVID-19 Host Genetic Initiative (data release 7). We performed a meta-analysis of up to 219,692 cases and over 3 million controls, identifying 51 distinct genome-wide significant loci—adding 28 loci from the previous data release^2^. The increased number of candidate genes at the identified loci helped to map three major biological pathways involved in susceptibility and severity: viral entry, airway defense in mucus, and type I interferon.

## Main Text

We conducted a meta-analysis for three phenotypes across 82 studies from 35 countries, including 36 studies with non-European ancestry (**Fig. 1, Supplementary Fig. 1, 2, Supplementary Table 1**): critical illness (respiratory support or death; 21,194 cases), hospitalization (49,033 cases), and SARS-CoV-2 infection (219,692 cases). Most of the studies were collected before the widespread introduction of COVID-19 vaccination. We found 30, 40, and 21 loci associated with critical illness, hospitalization, and infection due to SARS-CoV-2 respectively, for a total of 51 distinct genome-wide significant loci across all three phenotypes (*P* < 5 × 10^−8^; **Fig. 2, Supplementary Fig. 3, Supplementary Table 2**), adding 28 genome-wide significant loci to the 23 previously identified by the COVID-19 HGI (data release 6)^1,2^. We observed a median increase of 2.9-fold in statistical power across lead variants owing to a median increase of 1.6-fold in effective sample sizes from the previous release (**Supplementary Table 3**). After correcting for the number of phenotypes examined, 46 loci remain significant (*P* < 1.67 × 10^−8^). Of the 28 additional loci, six loci were originally reported by the GenOMICC^3^ which also contributed to the current meta-analysis; and nine other loci were identified by the new GenOMICC meta-analysis^4^ during the preparation of this manuscript. We found nine more loci reached genome-wide significance, but excluded them due to being likely false positives identified via newly developed leave-most-significant-biobank-out analysis (**Supplementary Table 4, Supplementary Note**). Comparing the effect sizes and statistical significance between the previous^2^ and current analysis indicated that all the previously identified loci were replicated and showed an increase in statistical significance (**Supplementary Fig. 4**). Using our previously developed two-class Bayesian model for classifying loci as being more likely involved in infection susceptibility or severity^2^, we determined that 36 loci are substantially more likely (> 99% posterior probability) to impact disease severity (hospitalization) and 9 loci to influence susceptibility to SARS-CoV-2 infection, while the remaining 6 loci could not be classified (**Supplementary Fig. 5, Supplementary Table 5, Supplementary Note**). We observed that 1q22 locus (lead variant: rs12752585:G>A) showed significant effect size heterogeneity across ancestries (*P*_het_ < 9.80 × 10^−4^ = 0.05 / 51), while the previously reported heterogenous locus (*FOXP4*) remained at the same level of significance as before^2^ despite increase in sample size (*P*_het_ = 2.01 × 10^−3^; **Supplementary Fig. 6, Supplementary Table 6**). We found significant observed-scale SNP heritabilities of all the three phenotypes (1.2–8.2%, *P* < 0.0001). We also estimated liability-scale heritabilities for a range of population prevalences (**Supplementary Fig. 7, Supplementary Table 7, Supplementary Note**).

**Fig. 1.**
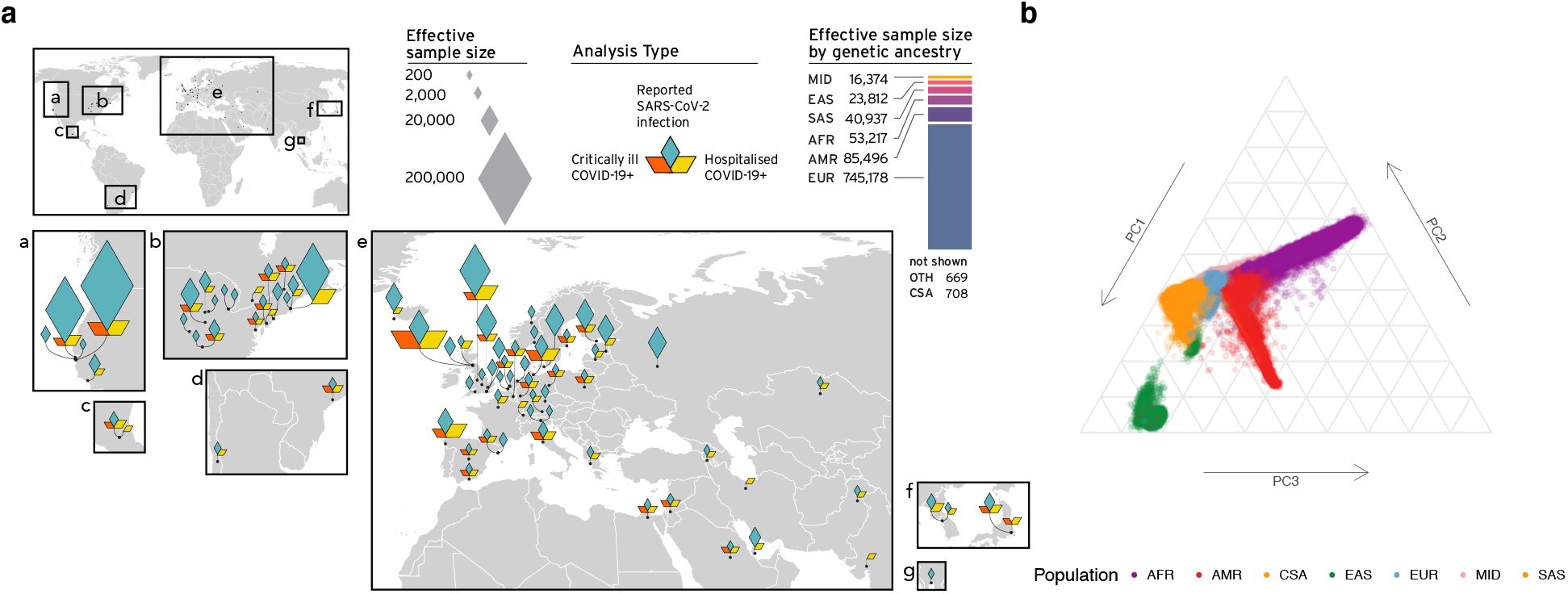
Overview of contributing studies in Host Genetics Initiative data release 7. **a**. Geographical overview of the contributing studies to the COVID-19 HGI and composition by major ancestry groups. Populations are defined as Middle Eastern (MID), South Asian (SAS), East Asian (EAS), African (AFR), Admixed American (AMR), European (EUR). **b**. Principal components analysis highlights the population structure and the sample ancestry of the individuals participating in the COVID-19 HGI. Per-cohort PCA results are available in **Supplementary Figure 2**. This figure is an updated version of the figure published by the COVID-19 Host Genetics Initiative^1,2^.

**Fig. 2.**
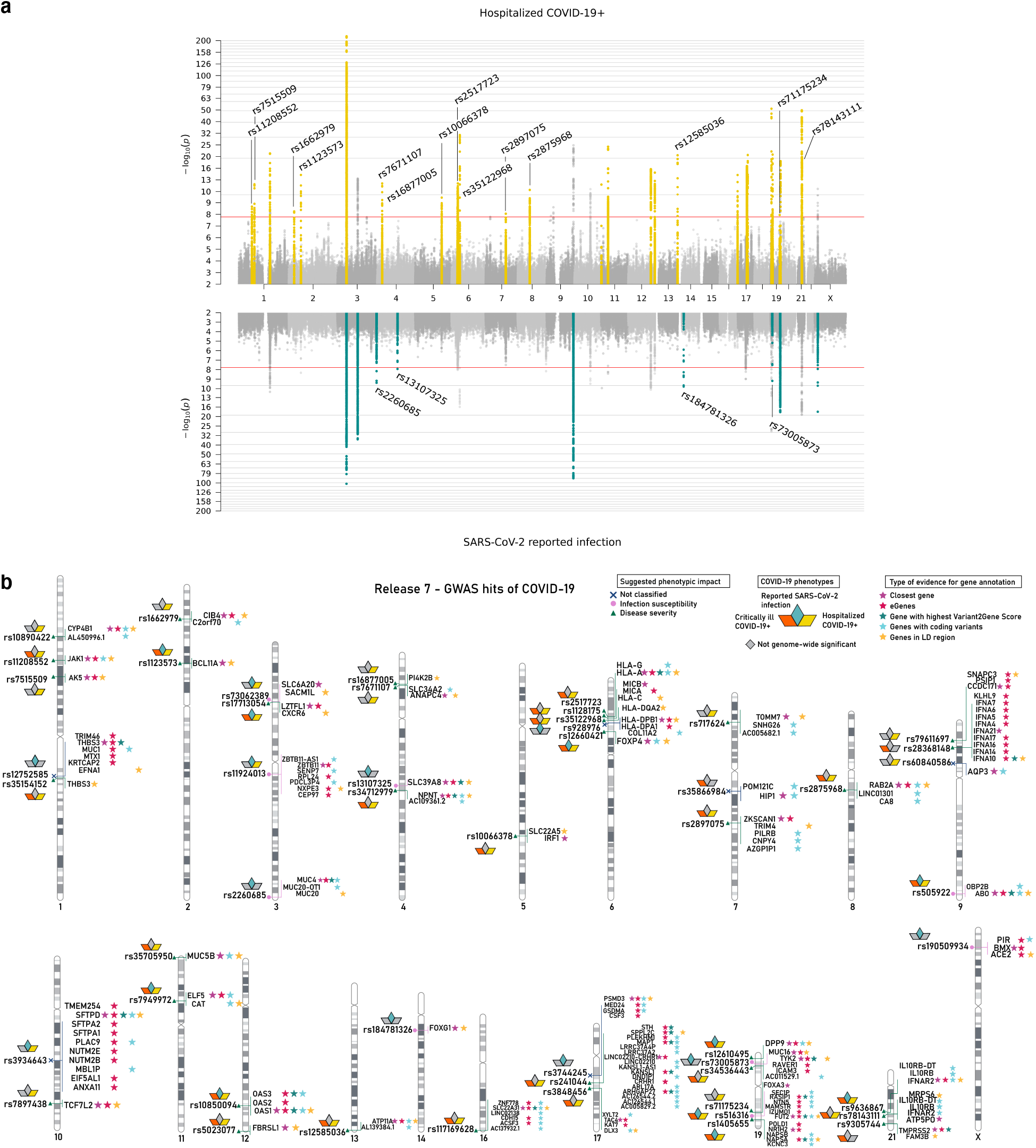
Genome-wide association results for COVID-19. **a**. Top panel shows results of genome-wide association study of hospitalized COVID-19 (*n* = 49,033 cases and *n* = 3,393,109 controls), and bottom panel the results of reported SARS-CoV-2 infection (*n* =219,692 cases and *n* = 3,001,905 controls). Loci highlighted in yellow (top panel) represent regions associated with severity of COVID-19 manifestation. Loci highlighted in green (bottom panel) are regions associated with susceptibility to SARS-CoV-2 infection. Lead variants for the loci newly identified in this data release are annotated with their respective rsID. **b**. Results of gene prioritization using different evidence measures of gene annotation. Genes in linkage disequilibrium (LD) region, genes with coding variants and eGenes (fine-mapped *cis*-eQTL variant PIP > 0.1 in GTEx Lung) are annotated if in LD with a COVID-19 lead variant (*r*^2^ > 0.6). V2G: Highest gene prioritized by OpenTargetGenetics’ V2G score. The pink circle indicates SARS-CoV-2 infection susceptibility, green triangle indicates COVID-19 severity, and blue cross indicates unclassified. This figure is an updated version of the figure published by the COVID-19 Host Genetics Initiative^1,2^.

To better understand the biological mechanisms underlying COVID-19 susceptibility and severity, we further characterized candidate causal genes by mapping them onto biological pathways and performing a phenome-wide association analysis (**Fig. 3, Supplementary Fig. 8, Supplementary Tables 2, 8, 9**). 15 out of 51 loci could be linked to three major pathways involved in susceptibility and severity defined by expert-driven classification (**Supplementary Note**): a) viral entry, b) entry defense in airway mucus, and c) type I interferon response. In addition, the phenome-wide association analysis identified nine loci involved in the upkeep of healthy lung tissue.

**Fig. 3.**
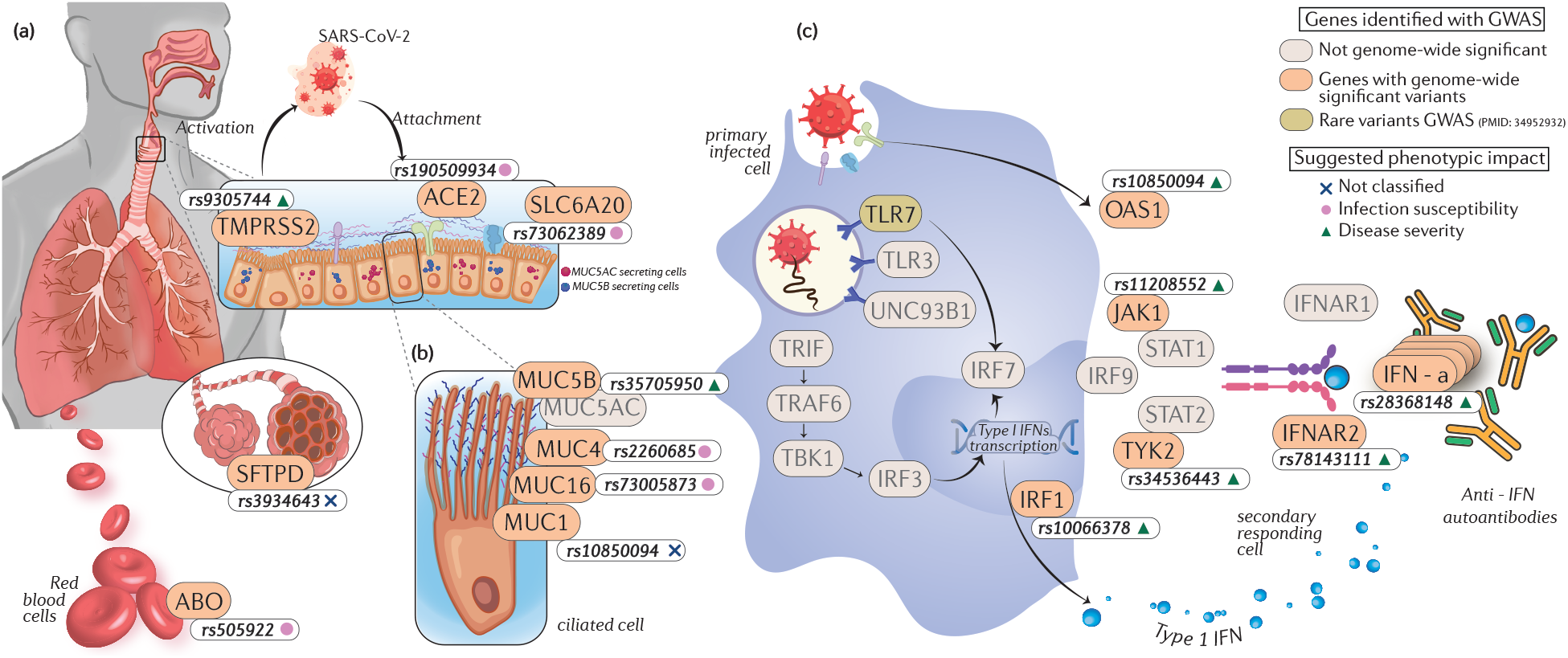
Major COVID-19 biological pathways mapped with susceptibility and severity GWAS loci. Genome-wide significant variants associated with COVID-19 (white boxes) and the annotated genes (peach boxes) are mapped on to pathways known to be involved in (**a**) viral entry and innate immunity, (**b**) entry defense in airway mucus, and (**c**) type I interferon. The suggested phenotypic impact of the significant variants using the Bayesian approach^2^ are denoted with shapes; COVID-19 susceptibility (pink circles), COVID-19 disease severity (green triangles), and unclassified variants (blue cross). Other genes known to be involved in the aforementioned pathways are shown using gray boxes. Detailed list of references to studies used to design this pathway can be found in **Supplementary Note**.

First, five loci include candidate causal genes involved in viral entry pathway (**Fig. 3a**), such as previously reported *SLC6A20* (3p21.31), *ABO* (9q34.2), *SFTPD* (10q22.3), and *ACE2* (Xp22.2), as well as *TMPRSS2* (21q22.3) which was additionally identified in the data release 7. We found a lead variant rs9305744:G>A, an intronic variant of *TMPRSS2*, is protective against critical illness (OR = 0.92, 95% CI = 0.89–0.95, *P* = 1.4 × 10^−8^) and in LD with a missense variant rs12329760:C>T (p.Val197Met; *r*^2^ = 0.68). SARS-CoV-2 employs the serine protease TMPRSS2 for viral spike (S) protein priming, as well as the previously reported ACE2 for host cell entry which functionally interacts with SLC6A20^5,6^. Of note, the previously reported association between ABO blood groups and susceptibility could be attributed to the interference of anti-A and anti-B antibodies with the S protein, potentially interfering with viral entry^7^. In addition, the previously reported *SFTPD* encodes pulmonary surfactant protein D (SP-D) which contributes dually to the lung’s innate immune molecules, and viral entry response in pulmonary epithelia^8,9^ along with other genes for airway defense.

Second, four loci contain candidate causal genes for entry defense in airway mucus (**Fig. 3b**), such as previously reported *MUC1*/*THBS3* (1q22) and *MUC5B* (11p15.5) as well as novel *MUC4* (3q29) and *MUC16* (19p13.2). We found that novel lead variants rs2260685:T>C in *MUC4* (intronic variant; in LD [*r*^2^ = 0.65] with a missense variant rs2259292:C>T [p.Gly4324Asp]) and rs73005873:G>A in *MUC16* (intronic variant) increase risk for SARS-CoV-2 infection (OR = 1.03 and 1.03, 95% CI = 1.02–1.04 and 1.02–1.04, *P* = 4.1 × 10^−8^ and 6.4 × 10^−10^, respectively). In addition, the previously reported locus 1q22 contains an intergenic lead variant rs12752585:G>A that decreases risk for the infection (OR = 0.98, 95% CI = 0.97–0.98, *P* = 1.5 × 10^−11^) and increases *MUC1* expression in esophagus mucosa in GTEx v8 (*P* = 5.2 × 10^−9^). Notably, the 1q22 locus also harbors an independent lead variant rs35154152:T>C, a missense variant (p.Ser279Gly) of *THBS3*, that decreases risk for hospitalization (OR = 0.88, 95% CI = 0.86–0.90, *P* = 5.6 × 10^−22^) but not infection (*P* = 5.7 × 10^−4^), suggesting potential distinct mechanisms in the locus. Consistent with these association patterns, MUC1, MUC4 and MUC16 are three known major transmembrane mucins of the respiratory tracts that prevent microbial invasion, while previously reported MUC5B, together with nearby MUC5AC, are primary structural components of airways mucus that enable mucociliary clearance of pathogens^10^.

Third, six loci harbor candidate causal genes that are linked to the type I interferon pathway (**Fig. 3c**), such as previously reported *IFNAR2* (21q22.11), *OAS1* (12q24.13), and *TYK2* (19p13.2), as well as additionally identified *JAK1* (1p31.3), *IRF1* (5q31.1) and IFN-α genes (9p21.3). Previous studies have reported additional genes in this pathway: *TLR7* (ref. ^11,12^) and *DOCK2* (ref. ^13^). In the present study, we found that a lead variant rs28368148:C>G, a missense variant (p.Trp164Cys) of *IFNA10* located within the IFN-α gene cluster, increases risk for critical illness (OR = 1.56, 95% CI = 1.38–1.77, *P* = 3.7 × 10^−12^). The IFN-α is one of the type I interferons that binds specifically to the IFN-α receptor consisting of IFNAR1/IFNAR2 chains, whose mutations are also known for increasing risk of hospitalization and critical illness. On the genes that enable signaling downstream of IFNAR, we identified a lead variant rs11208552:G>T, an intronic variant of *JAK1*, is protective against critical illness and hospitalization (OR = 0.92 and 0.95, 95% CI = 0.89–0.94 and 0.93–0.96, *P* = 5.5 × 10^−10^ and 2.2 × 10^−9^, respectively). This variant is previously reported to decrease lymphocyte counts^14^ (*β* = –0.016, *P* = 5.5 × 10^−15^) and increase the *JAK1* expression in thyroid in GTEx^15^ (*P* = 6.1 × 10^−23^). JAK1 and previously reported TYK2 are Janus kinases (JAKs) required for the type I interferon induced JAK-STAT signaling. JAK inhibitors are used to treat severe COVID-19 patients^16^. In addition, on the downstream of the JAK-STAT signaling, we found that a lead variant rs10066378:T>C, located 67 kb upstream of *IRF1*, increases risk for critical illness and hospitalization (OR = 1.09 and 1.07, 95% CI = 1.06–1.13 and 1.05–1.09, *P* = 2.7 × 10^−9^ and 3.74 × 10^−10^, respectively).

Furthermore, the phenome-wide association analysis identified nine loci previously associated with lung function and respiratory diseases. There loci contain genes involved in the upkeep of healthy lung tissue such as previously reported *FOXP4* (6p21.1), *SFTPD* (10q22.3), *MUC5B* (11p15.5), and *DPP9* (19p13.3), as well as additionally identified *CIB4* (2p23.3), *NPNT* (4q24), *ZKSCAN1* (7q22.1), *ATP11A* (13q34), and *PSMD3* (17q21.1). For example, we found that three lead variants rs1662979:G>T (intronic variant of *CIB4*), rs34712979:G>A (splice region variant of *NPNT*), and rs2897075:C>T (intronic variant of *ZKSCAN1*) significantly associated with hospitalization (OR = 1.05, 0.94, and 1.05, 95% CI = 1.03–1.07, 0.92–0.96 and 1.03–1.07, *P* = 5.6 × 10^−9^, 3.8 × 10^−8^ and 8.9 × 10^−9^, respectively) and lung function (FEV1/FVC)^17^, similar to the previously reported lead variant rs3934643:G>A (intronic variant of *SFTPD*). Notably, while the COVID-19 severity risk-increasing alleles of rs1662979 and rs3934643 decrease lung function (*β* = –0.013 and –0.025, *P* = 5.3 × 10^−8^ and 6.3 × 10^−10^), those of rs34712979 and rs2897075 increase it (*β* = 0.068 and 0.023, *P* = 4.2 × 10^−134^ and 1.6 × 10^−20^, respectively). Likewise, we found lead variants that significantly associated with hospitalization and idiopathic pulmonary fibrosis (IPF)^18,19^, such as the aforementioned rs2897075 and rs12585036:C>T (intronic variant of *ATP11A*; OR = 1.10, 95% CI = 1.08–1.12, *P* = 3.2 × 10^−21^), in addition to the previously reported rs35705950:G>T (promoter variant of *MUC5B*). While the COVID-19 severity risk-increasing alleles of rs2897075 and rs12585036 increase risk for IPF (OR = 1.12 and 1.27, *P* = 3.0 × 10^−14^ and 7.0 × 10^−9^, respectively), those of rs35705950 decreases (OR = 0.50, *P* = 3.9 × 10^−80^). These results highlight the complex pleiotropic relationships between COVID-19 severity, lung function, and respiratory diseases.

We applied genetic correlations and Mendelian randomization (MR) analyses to identify potential causal effects of risk factors on COVID-19 phenotypes (**Supplementary Fig. 9, Supplementary Tables 10, 11**). Fourteen novel genetic correlations and 10 novel robust exposure-COVID-19 trait pairs showed evidence of causal associations (**Supplementary Note**). In particular, smoking initiation and the number of cigarettes per day were positively correlated with severity and susceptibility phenotypes; with MR indicating that smoking was causally associated with increased risk for COVID-19 and further highlighting the role of the healthy lung tissue in COVID-19 severity. Additionally, genetically instrumented higher glomerular filtration rate (eGFR; indicative of better kidney function) was associated with lower risk of COVID-19 critical illness, while genetically predicted chronic kidney disease was associated with increased risk of COVID-19 critical illness, suggesting that better kidney function would be beneficial for lower risk of COVID-19 severity.

In summary, we have substantially expanded the current knowledge of host genetics for COVID-19 susceptibility and severity, by further doubling the case numbers from the previous data release^2^ and identifying 28 additional loci. The increased number of loci allows us to map genes to pathways involved in viral entry, airway defense and immune system response. Intriguingly, we observed severity loci mapped to type I interferon pathway, while susceptibility loci mapped to viral entry and airway defense pathways, with notable exceptions for severity-classified *TMPRSS2* and *MUC5B* loci. Further investigation of how such susceptibility and severity loci map to different pathways would provide mechanistic insights into the human genetic architecture of COVID-19.

## Data Availability

Summary statistics generated by COVID-19 HGI are available online, including per-ancestry summary statistics for African, Admixed American, East Asian, European, and South Asian ancestries　(https://www.covid19hg.org/results/r7/). The analyses described here utilize the data release 7. If available, individual-level data can be requested directly from contributing studies, listed in Supplementary Table 1. We used publicly available data from GTEx (https://gtexportal.org/home/), the Neale lab (http://www.nealelab.is/uk-biobank/), Finucane lab (https://www.finucanelab.org), FinnGen Freeze 4 cohort (https://www.finngen.fi/en/access_results), and eQTL catalogue release 3 (http://www.ebi.ac.uk/eqtl/).

## Data availability

Summary statistics generated by COVID-19 HGI are available online, including per-ancestry summary statistics for African, Admixed American, East Asian, European, and South Asian ancestries (https://www.covid19hg.org/results/r7/). The analyses described here utilize the data release 7. If available, individual-level data can be requested directly from contributing studies, listed in **Supplementary Table 1**. We used publicly available data from GTEx (https://gtexportal.org/home/), the Neale lab (http://www.nealelab.is/uk-biobank/), Finucane lab (https://www.finucanelab.org), FinnGen Freeze 4 cohort (https://www.finngen.fi/en/access_results), and eQTL catalogue release 3 (http://www.ebi.ac.uk/eqtl/).

## Code Availability

The code for summary statistics lift-over, the projection PCA pipeline including precomputed loadings and meta-analyses are available on GitHub (https://github.com/covid19-hg/); the code for custom analyses are available on GitHub—heritability estimation: https://github.com/AndrewsLabUCSF/COVID19_heritability; Mendelian randomization and genetic correlation: https://github.com/marcoralab/MRcovid; subtype analyses: https://github.com/mjpirinen/covid19-hgi_subtypes.

## Acknowledgments

We acknowledge M. O’Reilly and B. Cooley from the Pattern team at the Broad Institute of MIT and Harvard for designing the geographical map in **Fig. 1**. A full list of acknowledgement is available in **Supplementary Table 12**.

## Author contributions

Detailed author contributions are integrated in [the authorship list](https://docs.google.com/spreadsheets/d/1cp9pFeFUxXz5WMjRFv4X-AM1Hlc0iXYJa1rorSSj2Dc/edit#gid=2105248278).

