## Supplementary Information for "A second update on mapping the human genetic architecture of COVID-19"

### Supplementary Note

#### Online Methods

Ethical statements for each contributing study are given in **Supplementary Table 1**. Methods used in this study are described in our manuscripts from the previous releases[1,2], unless otherwise noted.

##### Identification of candidate false positive loci via leave-most-significant-out analysis

We implemented an additional quality control step on the meta-analysis results in order to identify potential false positive loci. For each genome-wide significant locus in any of the three analyses, we re-run a meta-analysis excluding the study with the smallest p-value. We called this approach “leave-most-significant-out”. Loci that were genome-wide significant in only one of the three phenotypes and had a “leave-most-significant-out” p-value > 0.001 were considered potential false positive (**Supplementary Table 4**), as their association was likely driven by one single study (the excluded, most significant one), and therefore excluded from the main results.

##### Loci with predicted classification in the previous release but not classified in the current one

Six loci could not be classified using our two-class Bayesian model[2] and setting a posterior probability threshold > 99%. Of these, 3 have been identified and classified in the previous release[2]: rs12752585:G>A and rs928976:C>T were classified as susceptibility variants in the previous release but showed a posterior probability for susceptibility of 98% and 58%, respectively, in the current one. rs3934643:G>A (*SFTPD*) was previously classified as a severity variant, but now reported a posterior probability for severity of 91%.

##### Conceptualization and design of the major COVID-19 biological pathways

The following studies were used to conceptualize and design major pathways of COVID-19 (**Fig. 3**) by Dr. Gita Pathak (Yale University). We then received expert guidance from immunologist Dr. Akiko Iwasaki (Yale University) to validate the diagrammatic representation of the pathways.

Briefly, ACE2 serves as the entry receptor for the SARS-CoV-2, while the TMPRSS2 serine protease primes the S protein[3]. *SFTPD* encodes pulmonary surfactant protein D (SP-D), which is an innate immune molecule of the pulmonary epithelia and has previously been known to aid in clearance of the mucosal entry points for viruses[4,5]. The ACE2 receptor and S protein during viral entry can be affected by natural anti-A and anti-B antibodies[6–9]. Mucin genes have a strong role in lung function by providing immune defenses to chronic respiratory conditions[10–12]. Lastly, we observed several genetic variants map to genes in the interferon pathway[13–16].

*Innate immunity*

- Goel et al., ABO blood group and COVID-19: a review on behalf of the ISBT COVID-19 Working Group (2021)[6]
- Bullerdiek et al., ABO blood groups and the risk of SARS-CoV-2 infection (2022)[7]
- Pendu et al., ABO Blood Types and COVID-19: Spurious, Anecdotal, or Truly Important Relationships? A Reasoned Review of Available Data (2021)[8]
- Tamayo-Velasco et al., ABO Blood System and COVID-19 Susceptibility: Anti-A and Anti-B Antibodies Are the Key Points (2022)[9]

- Meffre & Iwasaki, Interferon deficiency can lead to severe COVID (2021)[13]
- Zhou et al., Revisiting IRF1-mediated antiviral innate immunity (2022)[14]
- Merad & Martin, Pathological inflammation in patients with COVID-19: a key role for monocytes and macrophages (2020)[15]
- Gray et al., Severe COVID-19 represents an undiagnosed primary immunodeficiency in a high proportion of infected individuals (2022)[16]

*Mucin Biology*

- Denneny et al., Mucins and their receptors in chronic lung disease (2020)[10]
- Chatterjee et al., Defensive Properties of Mucin Glycoproteins during Respiratory Infections—Relevance for SARS-CoV-2 (2020)[11]
- Ridley & Thornton, Mucins: The frontline defence of the lung (2018)[12]

**Observed- and liability-scale SNP heritability estimates**

SNP heritability was estimated via the GenomicSEM implementation of LDSC using only EUR summary statistics, a EUR LD reference panel, the sum of the effective sample size (the sample size for an equivalently powered GWAS within a balanced sample—*i.e.*, 50% cases and 50% controls) across the contributing cohorts, a sample prevalence of 0.5, and across a range of population-based prevalence (1%–90%)[17]. Using the summation of the effective sample size across cohorts, rather than total sample size or effective sample size calculated using total sample prevalence, accounts for differences in sample prevalence across contributing cohorts in a meta-analysis. Estimating heritability without accounting for differences in ascertainment can result in a downward bias by as much as 30% (ref. [17]). SNP heritability for all three COVID-19 related phenotypes was significant on the observed-scale for all the three phenotypes (1.2–8.2%, $P <$ 0.0001). Liability-scale heritabilities were estimated to range between 0.0066–0.013, 0.032–0.061, and 0.045–0.086 for reported infection, hospitalization, and critical illness, respectively (**Supplementary Table 7**).

**Genetic Correlations and Mendelian Randomization**

The genetic correlation, heritability estimates, and Mendelian Randomization were performed as per methods described in data release 5 (ref. [1]).

To understand which traits are genetically correlated and/or potentially causally related to the three phenotypes, we first estimated genetic correlations with 38 traits (**Supplementary Table 10**). In addition to what was previously reported, novel positive genetic correlations were observed between critical illness and cigarettes per day, idiopathic pulmonary fibrosis, C-Reactive protein levels, and depression; and negative genetic correlations with autism spectrum disorder. For hospitalization, positive genetic correlations were observed for rheumatoid arthritis, risk tolerance, and idiopathic pulmonary fibrosis; and negative genetic correlations with high density lipoprotein levels and eGFR. Finally, for SARS-CoV-2 infection, positive genetic correlations were observed with cigarettes per day, rheumatoid arthritis, and depression; and negative genetic correlations with red blood cell count.

We next applied two-sample Mendelian Randomization (MR) to infer potential causal relationships between COVID-19 related phenotypes and their genetically correlated traits. After correcting for multiple testing and were robust to potential violations of the underlying assumptions of MR 10 novel causal associations were observed (**Supplementary Table 11**). Cigarettes per day, eGFR, chronic kidney disease, and ADHD with critical illness; smoking initiation and cigarettes per day with hospitalization; and red blood cell count, white blood cell count, CRP, and smoking initiation with reported infection.

#### Supplementary Figures

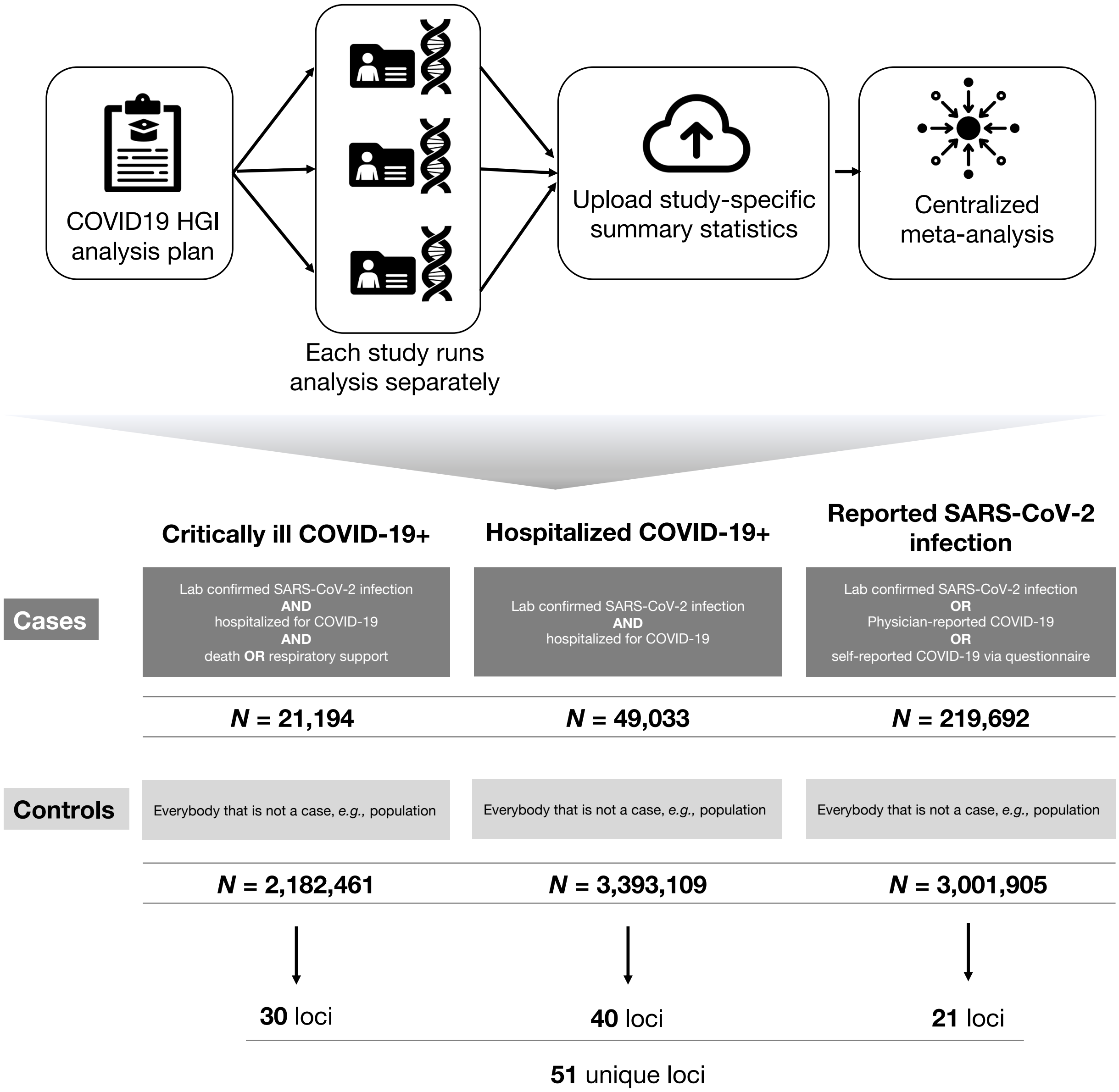


**Supplementary Fig. 1 | Analytical summary of the COVID-19 HGI meta-analysis**. Using the analytical plan set by the COVID-19 HGI[1], each individual study runs their analyses and uploads the results to the Initiative, who then runs the meta-analysis. There are three main analyses that each study can contribute summary statistics to: critically ill COVID-19, hospitalized COVID-19 and reported SARS-CoV-2 infection. The phenotypic criteria used to define cases are listed in the dark gray boxes, along with the numbers of cases included in the final all ancestries meta-analysis. Controls were defined in the same way across all three analyses: as everybody that is not a case *e.g.,* population controls (light gray box). Sample number of controls differed between the analyses due to the difference in the number of studies contributing data to these.

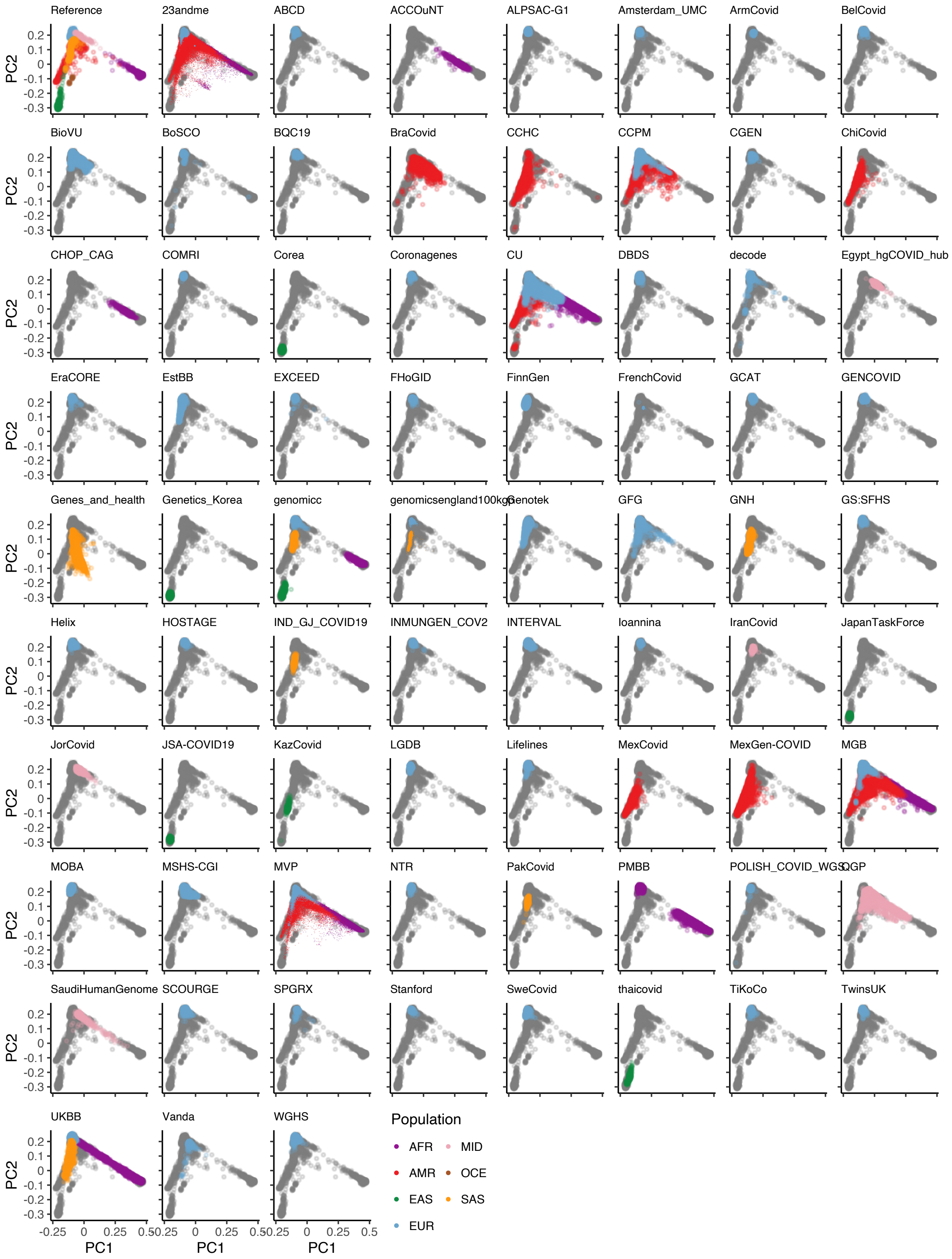

Reference
23andme
ABCD
ACCOuNT
ALPSAC-G1
Amsterdam_UMC
ArmCovid
BelCovid
BioVU
BoSCO
BQC19
BraCovid
CCHC
CCPM
CGEN
ChiCovid
CHOP_CAG
COMRI
Corea
Coronagenes
CU
DBDS
decode
Egypt_hgCOVID_hub
EraCORE
EstBB
EXCEED
FHoGID
FinnGen
FrenchCovid
GCAT
GENCOVID
Genes_and_health
Genetics_Korea
genomicc
genomicsengland100kgp
Genotek
GFG
GNH
GS:SFHS
Helix
HOSTAGE
IND_GJ_COVID19
INMUNGEN_COV2
INTERVAL
Ioannina
IranCovid
JapanTaskForce
JorCovid
JSA-COVID19
KazCovid
LGDB
Lifelines
MexCovid
MexGen-COVID
MGB
MOBA
MSHS-CGI
MVP
NTR
PakCovid
PMBB
POLISH_COVID_WGS
QGP
SaudiHumanGenome
SCOURGE
SPGRX
Stanford
SweCovid
thaicovid
TiKoCo
TwinsUK
UKBB
Vanda
WGHS
PC2
0.2
0.1
0
-0.1
-0.2
-0.3
PC1
-0.25
0
0.25
0.5
Population
AFR
MID
AMR
OCE
EAS
SAS
EUR

**Supplementary Fig. 2 | Projection of contributed samples from participating studies into the same PC space**. We asked participating studies to perform PC projection using the 1000 Genomes Project and Human Genome Diversity Project as a reference, with a common set of variants. For each panel (except for the reference), colored points correspond to contributed samples from each cohort, whereas gray points correspond to the 1000 Genomes reference samples. Color represents a genetic population that each cohort specified. Since 23andme, FrenchCovid, genomicsengland100kgp, and MVP only submitted PCA images, we overlaid their submitted transparent images using the same coordinates, instead of directly plotting them. We excluded studies that were not able to submit PCA results.

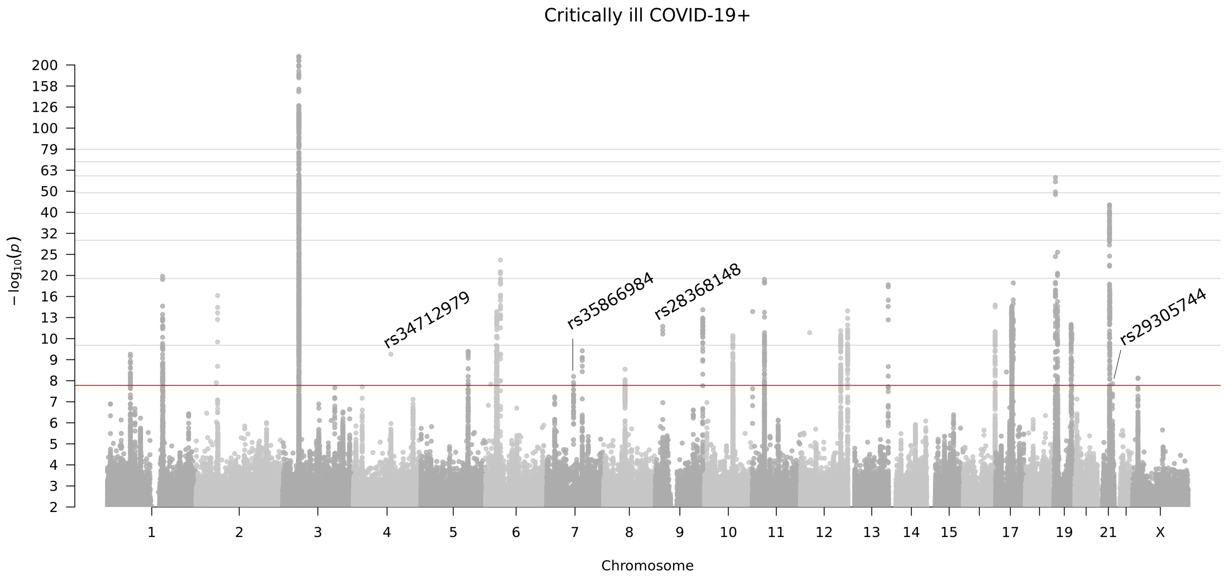


**Supplementary Fig. 3 | Genome-wide association results for COVID-19 critical illness (Release 7).** Results of genome-wide association study of critically ill COVID-19 cases vs population controls (21,194 cases and 2,182,461 controls). Critically ill COVID-19 cases defined as those who required respiratory support in hospital or who were deceased due to the disease.

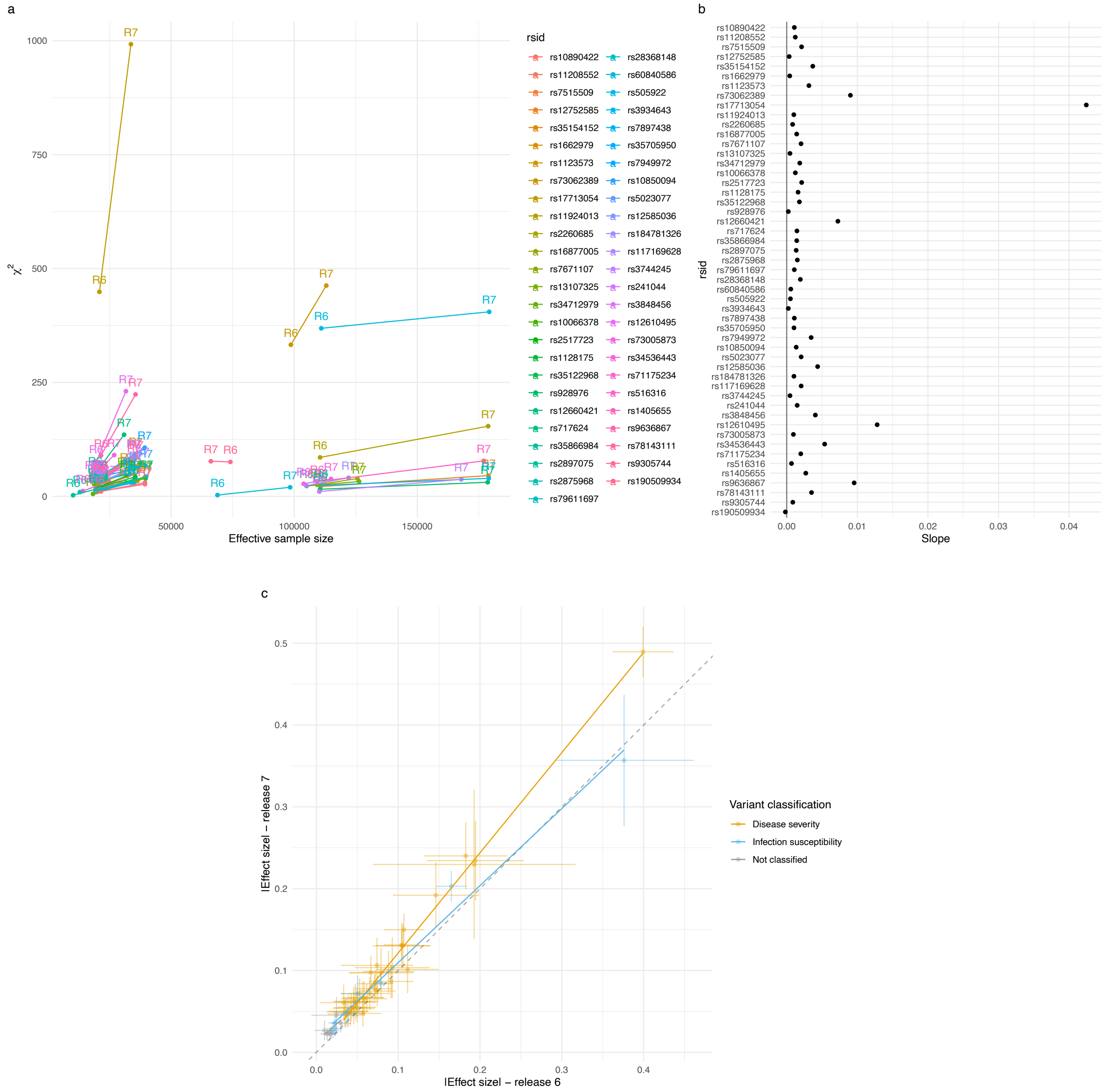


**Supplementary Fig. 4 | Comparison of associations between release 6 and release 7. a**. $\chi2$ statistics (y-axis) and effective sample size (x-axis) in release 6 (dots labeled as R6) and release 7 (dots labeled as R7), for each variant reported in release 7. **b**. Slope of the lines represented in panel **a**, that is, the change in $\chi2$ statistics for each additional unit in the effective sample size, for each variant reported in release 7. **c**. Effect size absolute values in release 6 (x-axis) and release 7 (y-axis) and standard errors, for each variant reported in release 7.

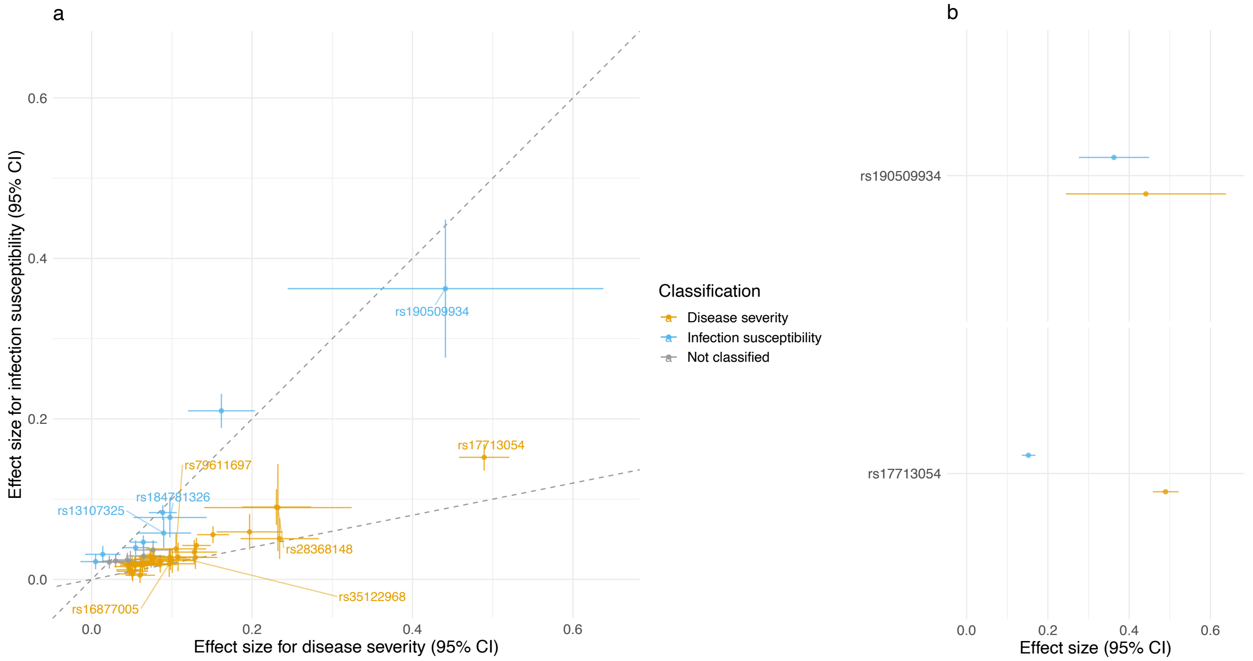


**Supplementary Fig. 5 | The estimated log odds ratios for 51 lead variants shown for COVID-19 severity GWAS on x-axis and SARS-CoV-2 GWAS on y-axis. a**. Effect sizes, 95% confidence intervals and classification for all reported loci. Classification of variants affecting infection susceptibility or disease severity is shown by colors and corresponds to **Supplementary Table 5**. The two lines show the expected relationship between effect sizes of variants affecting susceptibility to infection (line y = x) or disease severity (line y = 0.2x). RsIDs are reported only for newly discovered loci. **b**. Effect sizes for rs190509934 and rs17713054 to illustrate the principle beyond the model used for the classification. Effect sizes for the disease severity and infection susceptibility analyses overlap in the case of a susceptibility variant (rs190509934), while for a severity variant (rs17713054) the effect size for the infection susceptibility analysis is neglectable compared to the one for the disease severity analysis.

Figures enclosed in a separate PDF.

**Supplementary Fig. 6 | Forest plots for highly heterogeneous loci across ancestries**. Forest plots for 1q22 locus (lead variant: rs12752585:G>A) and *FOXP4* locus (lead variant: rs12660421:G>A). The 1q22 locus showed a significant heterogeneous effect across studies ($P_{het}$ < $9.80 \times 10^{-4}$ = 0.05 / 51), while the FOXP4 locus remained at the same level of significance as before[2] ($P_{het}$ = $2.01 \times 10^{-3}$; **Supplementary Table 6**). For each of the loci, effect sizes and confidence intervals are reported for each contributing study. Studies are grouped by ancestry, with summary effects reported for each ancestry subgroup. The summary effect size across all studies is also reported in the bottom panel for each locus.

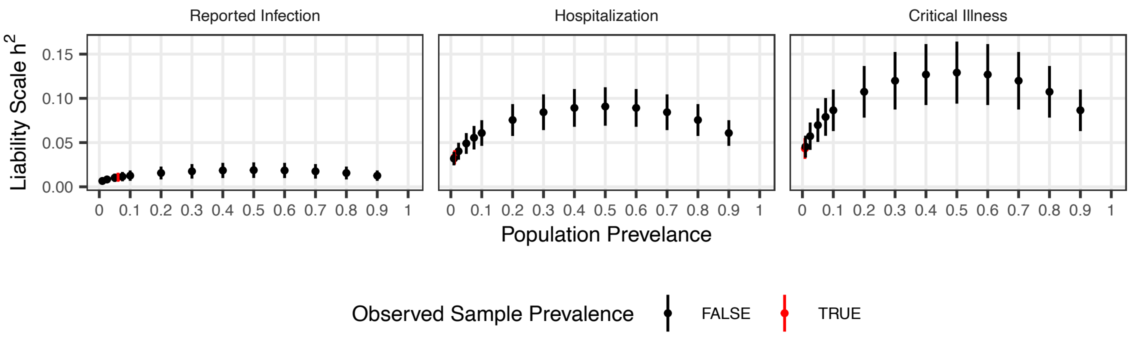


**Supplementary Fig. 7 | Estimated liability-scale SNP heritabilities of the three COVID-19 phenotypes across a range of population prevalences.** Liability-scale $h^2$ estimates and 95% confidence intervals. SNP-heritability was estimated using the summation of the effective sample sizes across contributing cohorts. Population prevalence was estimated using the total sample prevalence (red) or across a range of population prevalence (black; 1%–90%).

Figures enclosed in a separate PDF.

**Supplementary Fig. 8 | LocusZoom plots to visualize the meta-analysis results at the loci passing genome-wide significance**. For each genome-wide significant locus in three meta-analyses: meta-analysis of critical illness, hospitalization, and reported infection, we showed 1) a Manhattan plot of each locus where a color represents a weighted-average r2 value (see COVID-19 Host Genetics Initiative, 2021) to a lead variant (unadjusted P-values from the two-tailed inverse variance weighted meta-analysis); 2) r2 values to a lead variant across gnomAD v2 populations, i.e., African/African-American (AFR), Latino/Admixed American (AMR), Ashkenazi Jewish (ASJ), East Asian (EAS), Estonian (EST), Finnish (FIN), Non-Finish Europeans (NFE), North-Western Europeans (NWE), and Southern Europeans (SEU); 3) genes at a locus; and 4) genes prioritized by each gene prioritization metric where a size of circles represents a rank in each metric. Note that the COVID-19 lead variants were chosen across all the meta-analyses (**Supplementary Table 2**) and were not necessarily a variant with the most significant P-value from each inverse variance weighted meta-analysis.

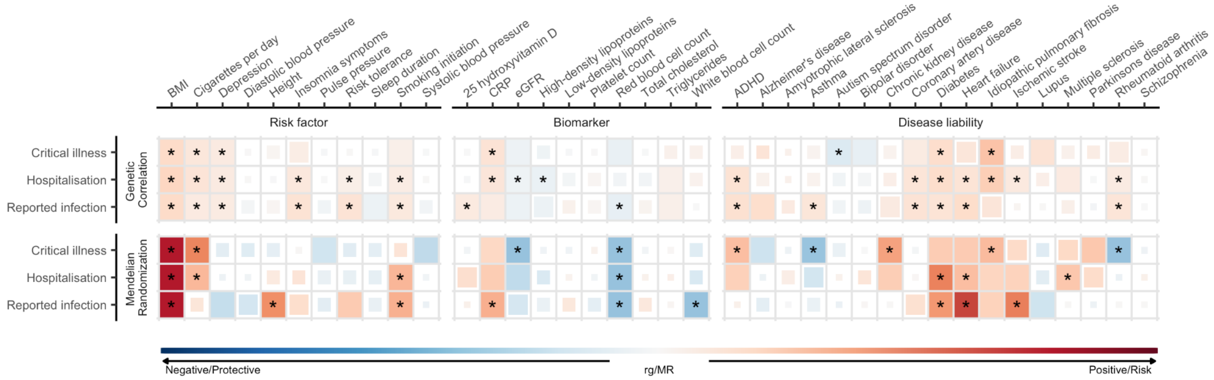


**Supplementary Fig. 9 | Genetic correlations and Mendelian randomization causal estimates between 38 traits and COVID-19 critical illness, hospitalization, and SARS-CoV-2 reported infection.** Larger squares correspond to more significant P-values, with genetic correlations or MR causal estimates significantly different from zero at a $P < 0.05$ shown as a full-sized square. Genetic correlations or causal estimates that are significantly different from zero at a false discovery rate (FDR) of 5% are marked with an asterisk. Two-sided *P*-values were calculated using LDSC for genetic correlations and Inverse variance weighted analysis for MR.
