## Supplementary figures and images for "A second update on mapping the human genetic architecture of COVID-19"

### Supplementary Fig. 6

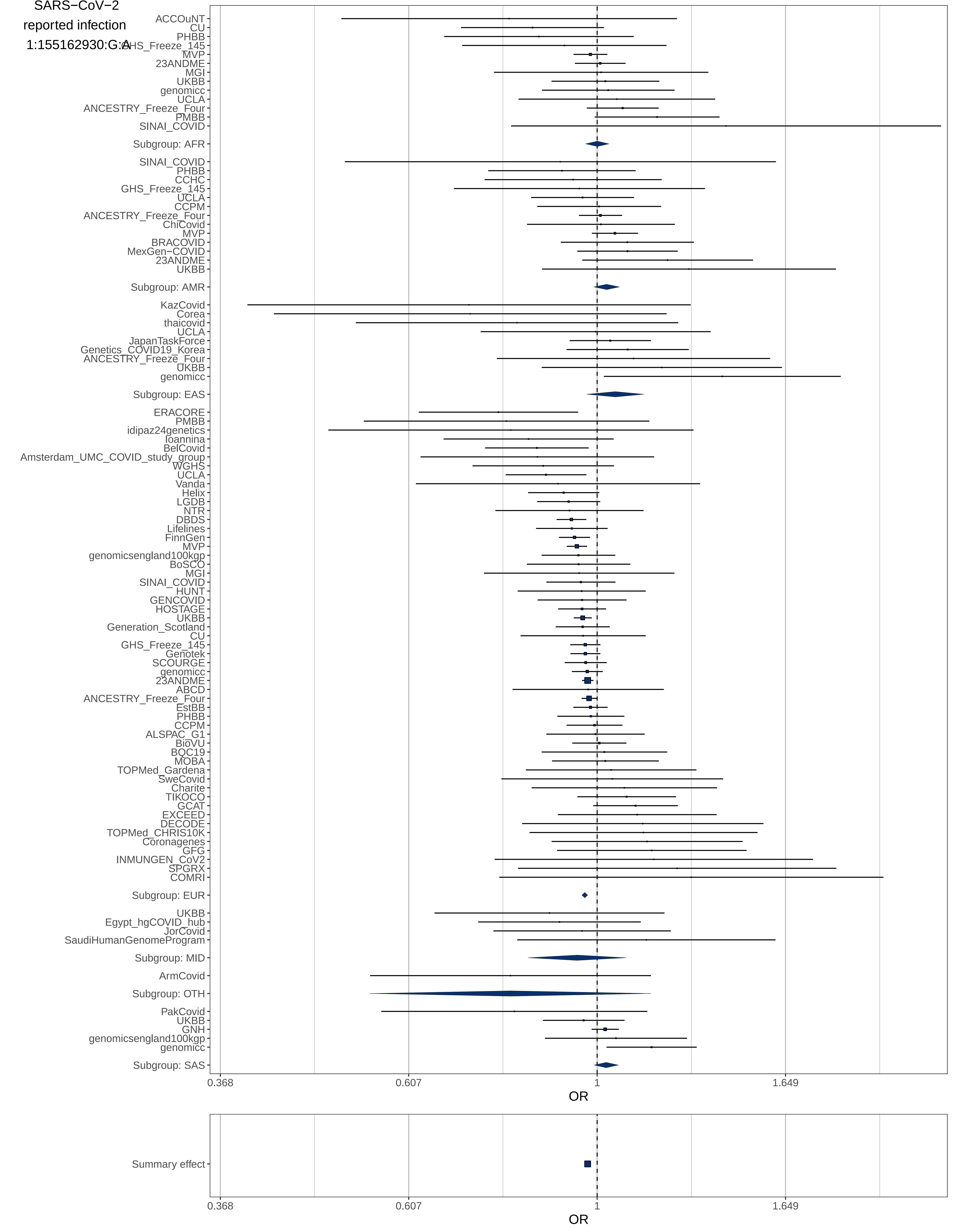

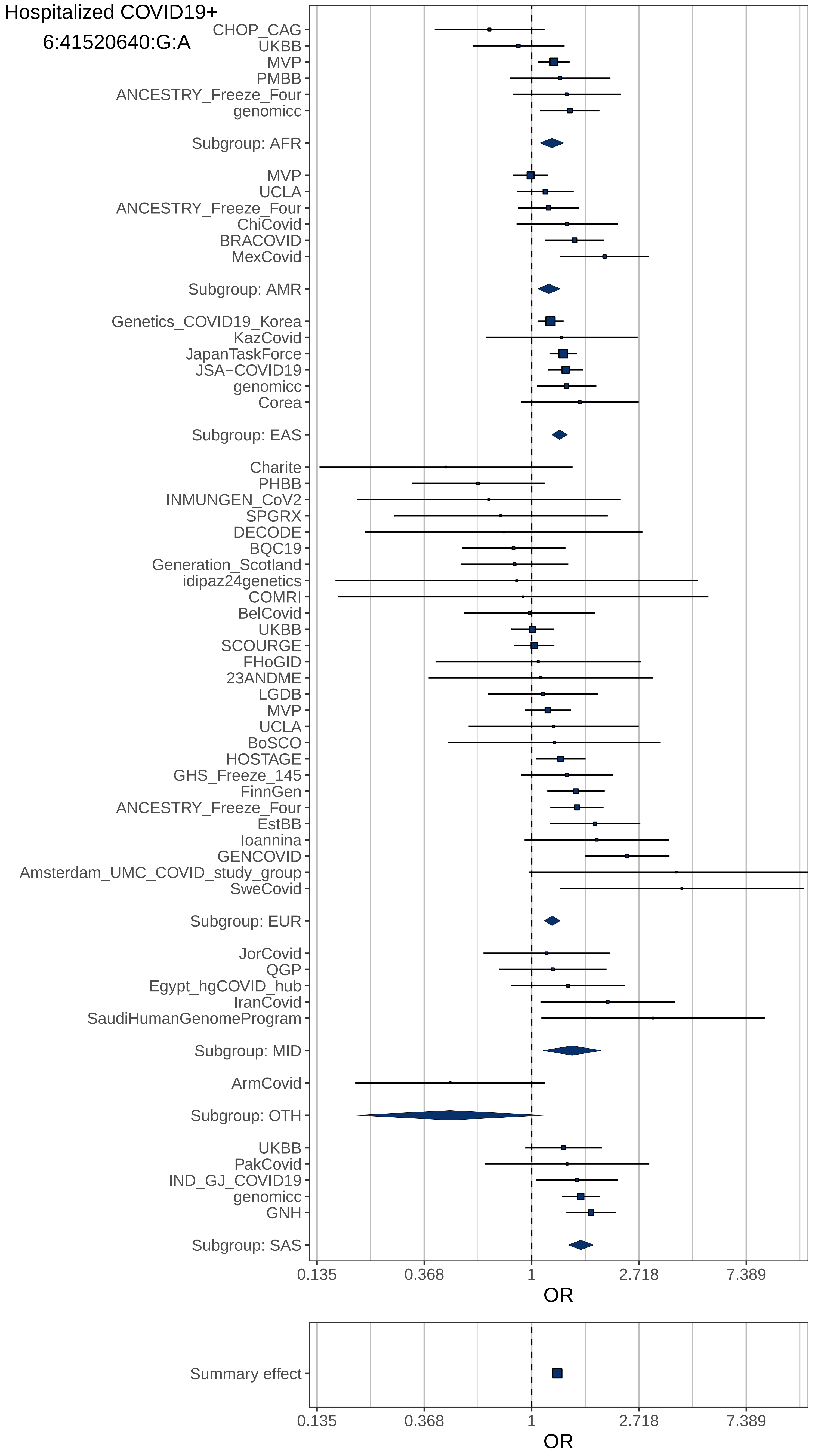
