## Supplementary Fig. 8 for "A second update on mapping the human genetic architecture of COVID-19"

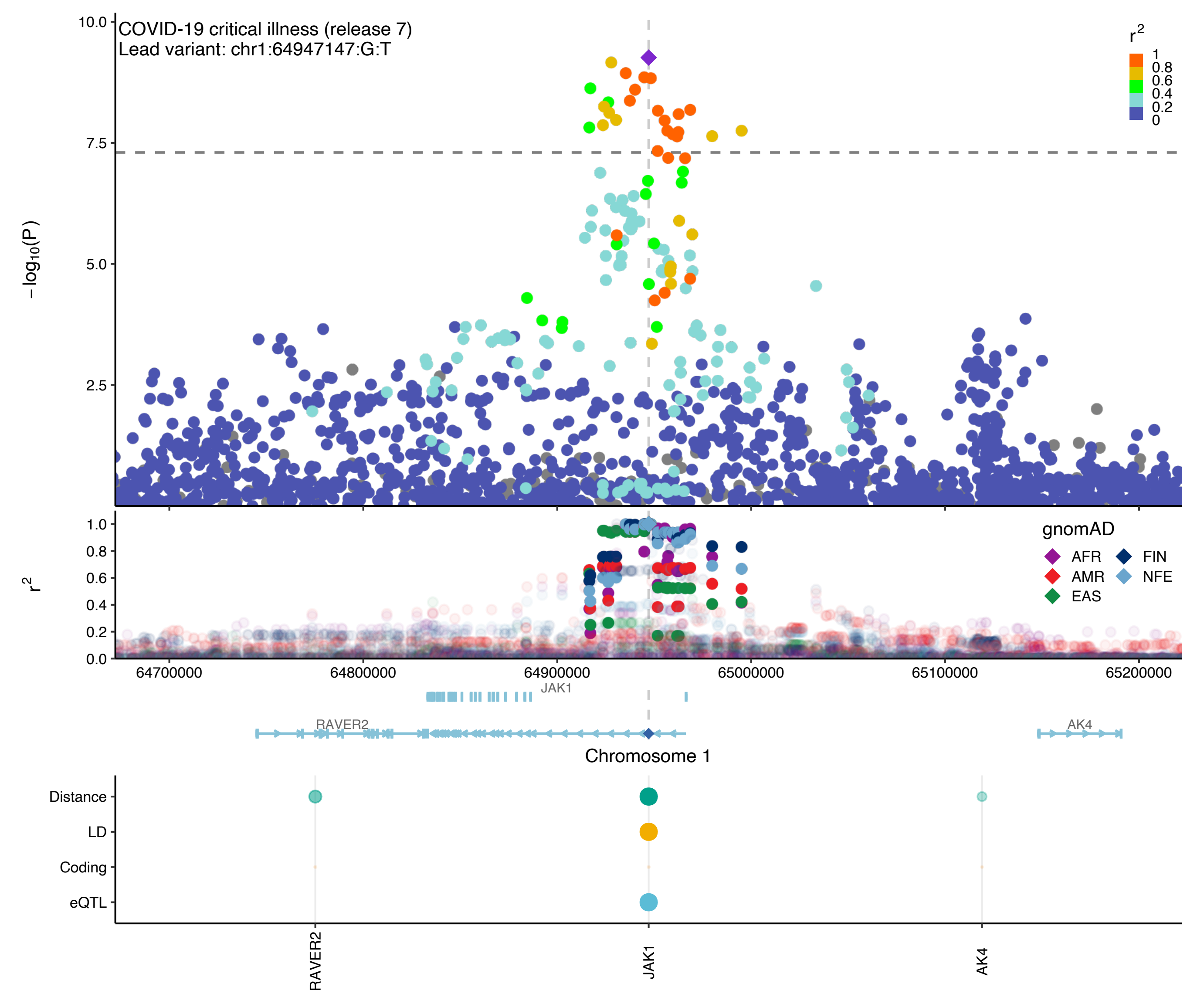

COVID-19 critical illness (release 7)
Lead variant: chr1:64947147:G:T
−log10(P)
10.0
7.5
5.0
2.5
r2
1
0.8
0.6
0.4
0.2
0
1.0
0.8
0.6
0.4
0.2
0.0
gnomAD
AFR
FIN
AMR
NFE
EAS
64700000
64800000
64900000
65000000
65100000
65200000
JAK1
RAVER2
AK4
Chromosome 1
Distance
LD
Coding
eQTL
RAVER2
JAK1
AK4

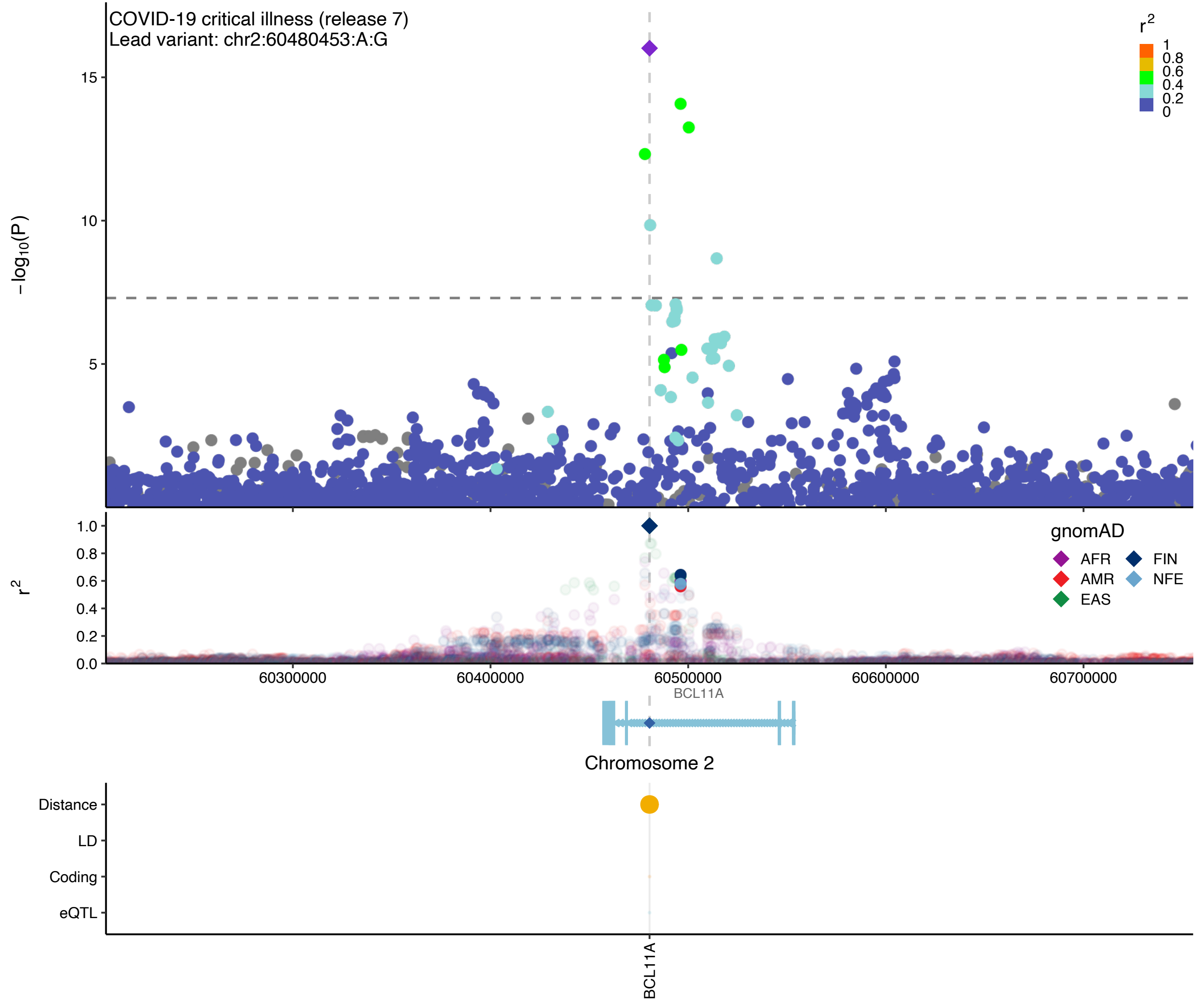

COVID-19 critical illness (release 7)
Lead variant: chr2:60480453:A:G
$-\log_{10}(P)$
15
10
5
$r^2$
1
0.8
0.6
0.4
0.2
0
1.0
0.8
0.6
0.4
0.2
0.0
$r^2$
gnomAD
AFR
FIN
AMR
NFE
EAS
60300000
60400000
60500000
60600000
60700000
BCL11A
Chromosome 2
Distance
LD
Coding
eQTL
BCL11A

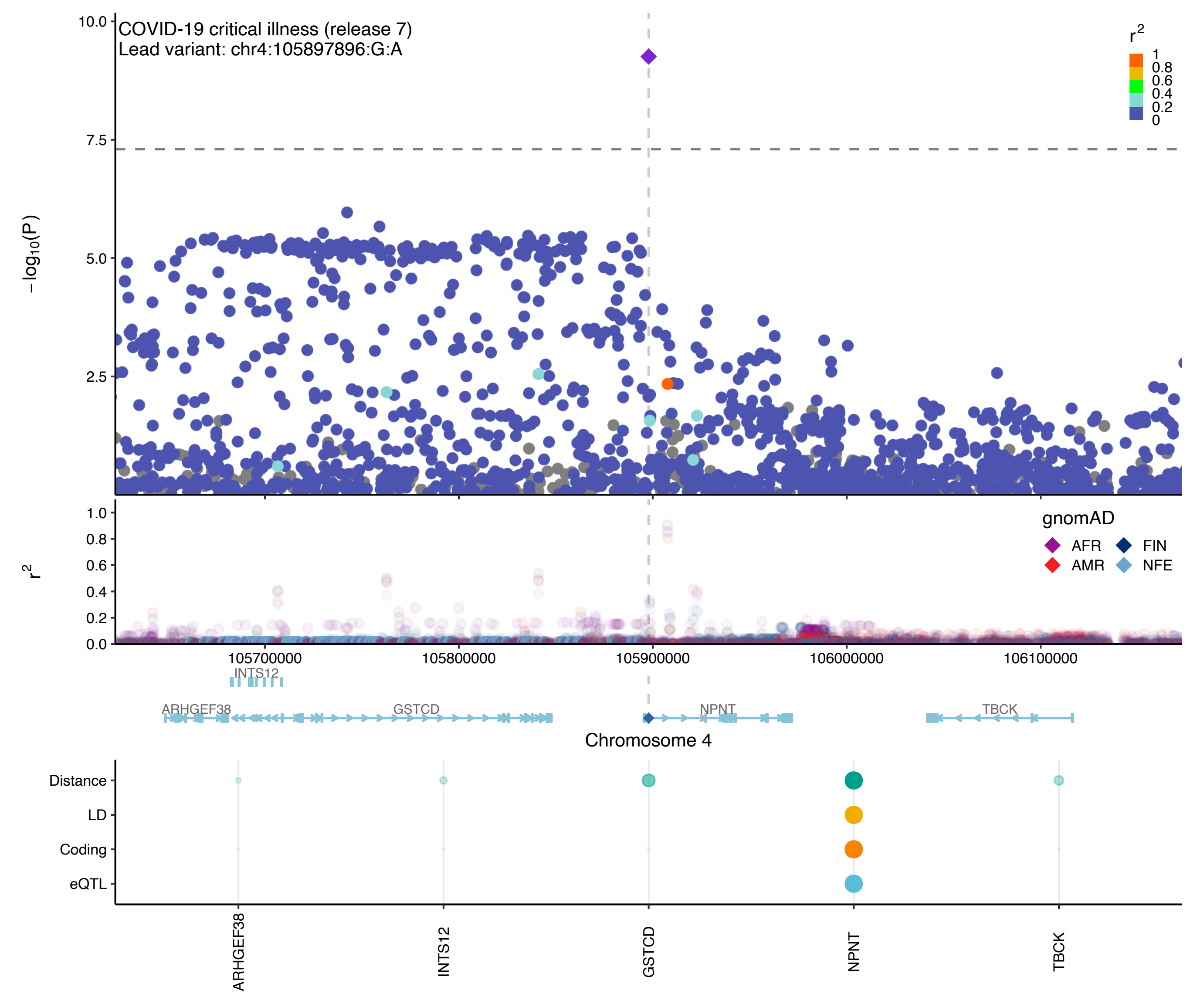

COVID-19 critical illness (release 7)
Lead variant: chr4:105897896:G:A
−log10(P)
10.0
7.5
5.0
2.5
r2
1
0.8
0.6
0.4
0.2
0
r2
1.0
0.8
0.6
0.4
0.2
0.0
gnomAD
AFR
FIN
AMR
NFE
105700000
105800000
105900000
106000000
106100000
INTS12
ARHGEF38
GSTCD
NPNT
TBCK
Chromosome 4
Distance
LD
Coding
eQTL
ARHGEF38
INTS12
GSTCD
NPNT
TBCK

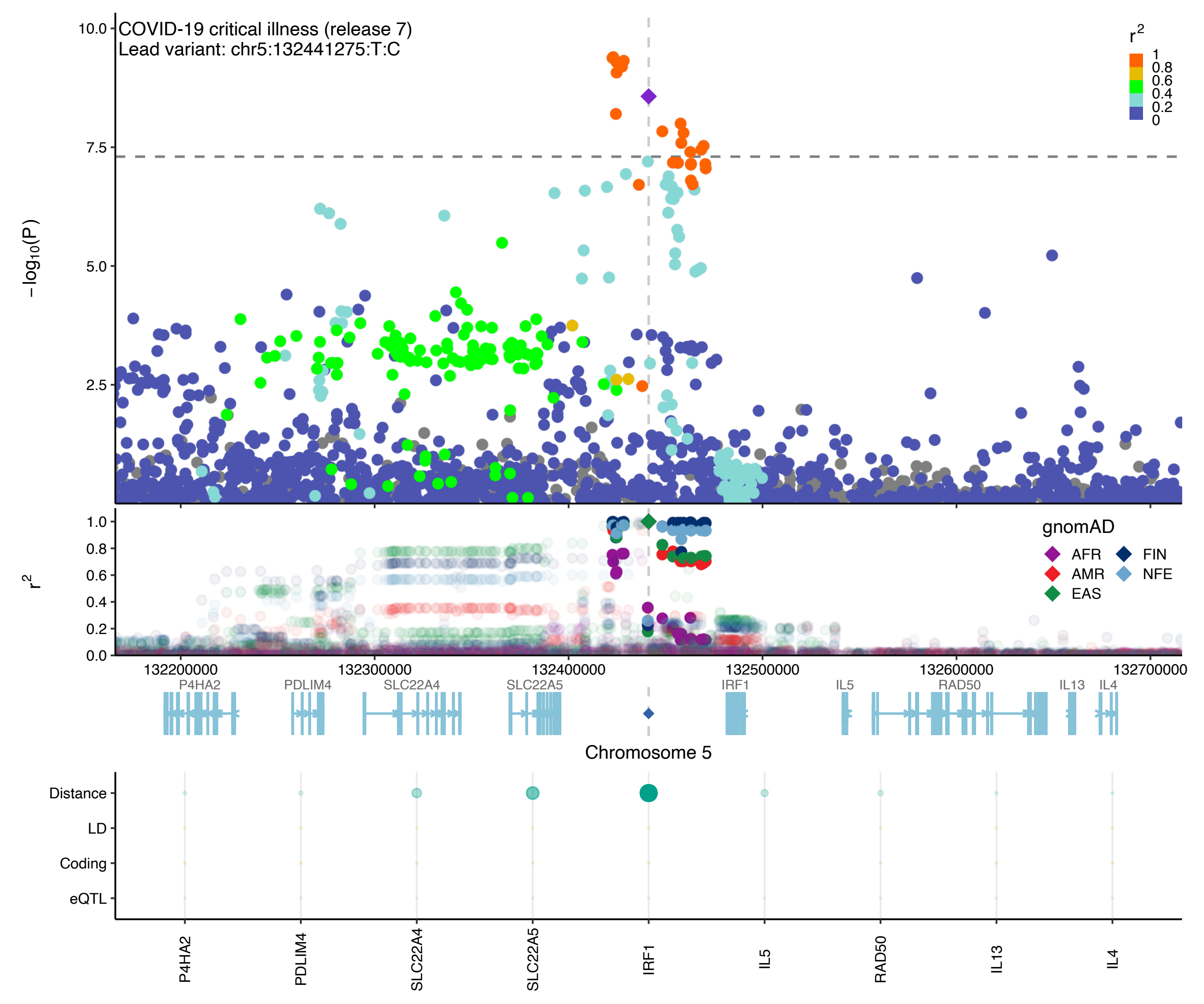

COVID-19 critical illness (release 7)
Lead variant: chr5:132441275:T:C
−log10(P)
10.0
7.5
5.0
2.5
r2
1
0.8
0.6
0.4
0.2
0
gnomAD
AFR
AMR
EAS
FIN
NFE
1.0
0.8
0.6
0.4
0.2
0.0
132200000
132300000
132400000
132500000
132600000
132700000
P4HA2
PDLIM4
SLC22A4
SLC22A5
IRF1
IL5
RAD50
IL13
IL4
Chromosome 5
Distance
LD
Coding
eQTL
P4HA2
PDLIM4
SLC22A4
SLC22A5
IRF1
IL5
RAD50
IL13
IL4

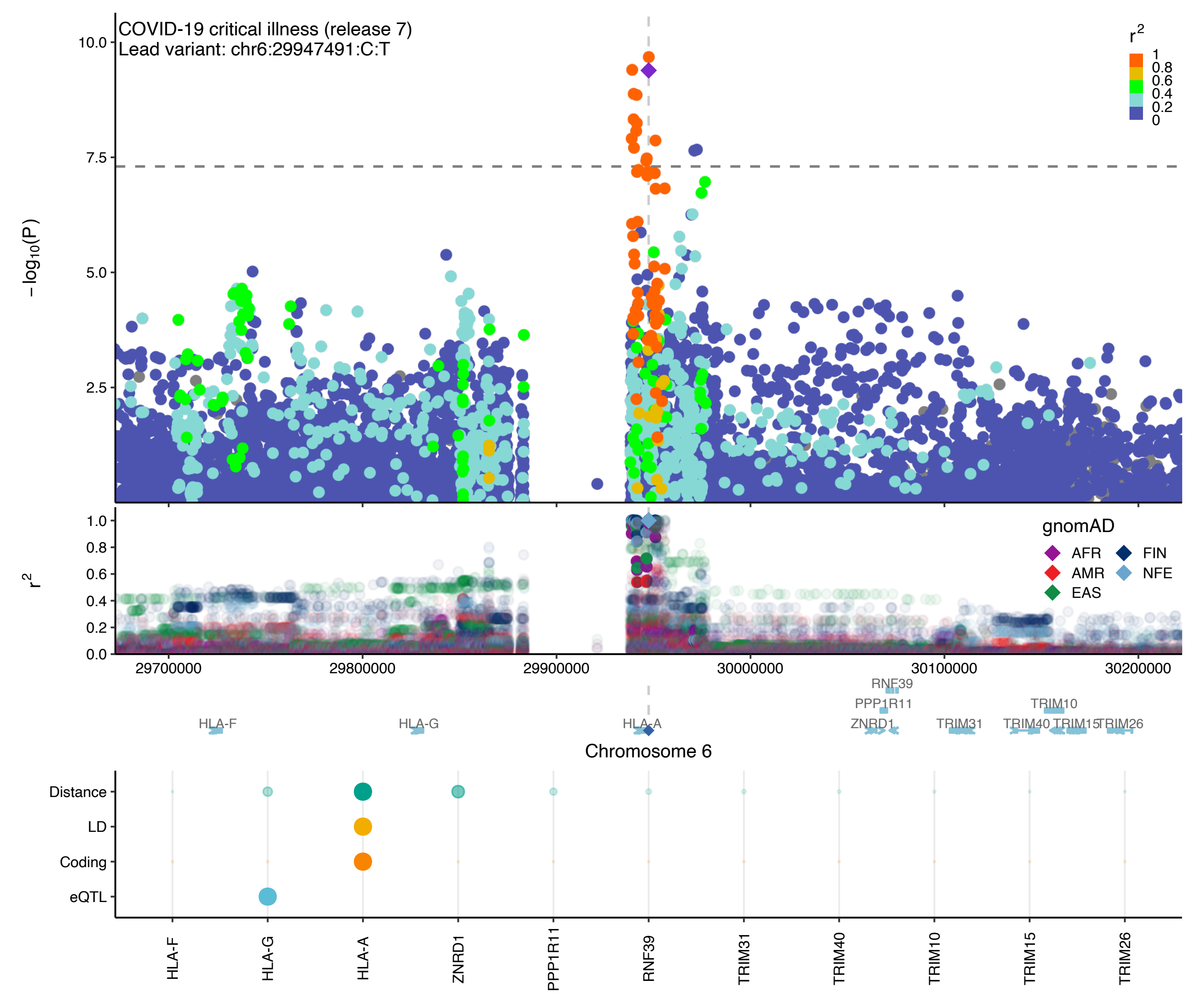
COVID-19 critical illness (release 7)
Lead variant: chr6:29947491:C:T
$-\log_{10}(P)$
10.0
7.5
5.0
2.5
$r^2$
1
0.8
0.6
0.4
0.2
0
1.0
0.8
0.6
0.4
0.2
0.0
gnomAD
AFR
FIN
AMR
NFE
EAS
29700000
29800000
29900000
30000000
30100000
30200000
RNF39
PPP1R11
TRIM10
HLA-F
HLA-G
HLA-A
ZNRD1
TRIM31
TRIM40
TRIM15
TRIM26
Chromosome 6
Distance
LD
Coding
eQTL
HLA-F
HLA-G
HLA-A
ZNRD1
PPP1R11
RNF39
TRIM31
TRIM40
TRIM10
TRIM15
TRIM26

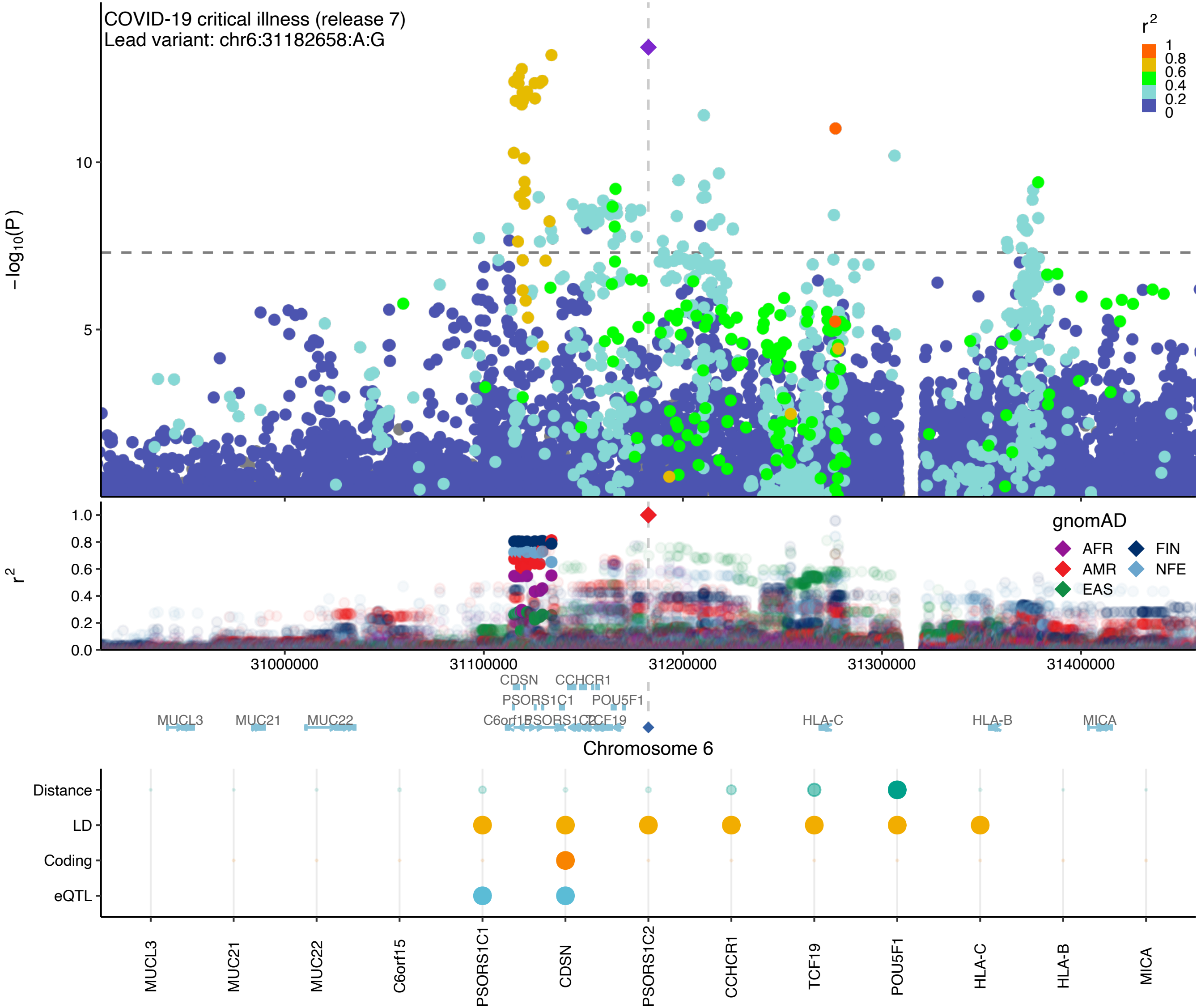

COVID-19 critical illness (release 7)
Lead variant: chr6:31182658:A:G
r2
1
0.8
0.6
0.4
0.2
0
−log10(P)
10
5
r2
1.0
0.8
0.6
0.4
0.2
0.0
gnomAD
AFR
AMR
EAS
FIN
NFE
31000000
31100000
31200000
31300000
31400000
CDSN
CCHCR1
PSORS1C1
POU5F1
MUCL3
MUC21
MUC22
HLA-C
HLA-B
MICA
Chromosome 6
Distance
LD
Coding
eQTL
MUCL3
MUC21
MUC22
C6orf15
PSORS1C1
CDSN
PSORS1C2
CCHCR1
TCF19
POU5F1
HLA-C
HLA-B
MICA

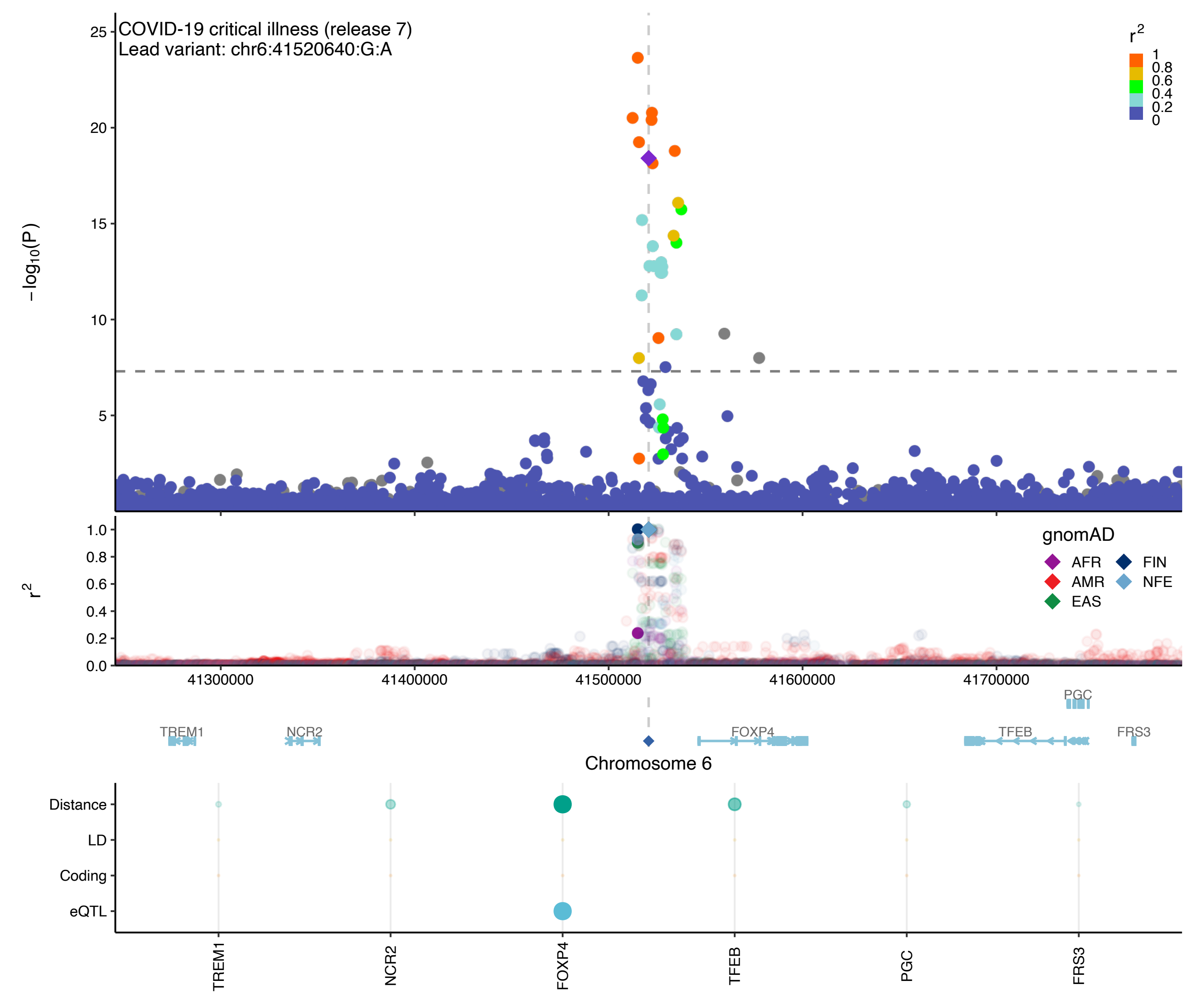

COVID-19 critical illness (release 7)
Lead variant: chr6:41520640:G:A
−log10(P)
r2
1
0.8
0.6
0.4
0.2
0
gnomAD
AFR
AMR
EAS
FIN
NFE
41300000
41400000
41500000
41600000
41700000
TREM1
NCR2
FOXP4
TFEB
PGC
FRS3
Chromosome 6
Distance
LD
Coding
eQTL
TREM1
NCR2
FOXP4
TFEB
PGC
FRS3

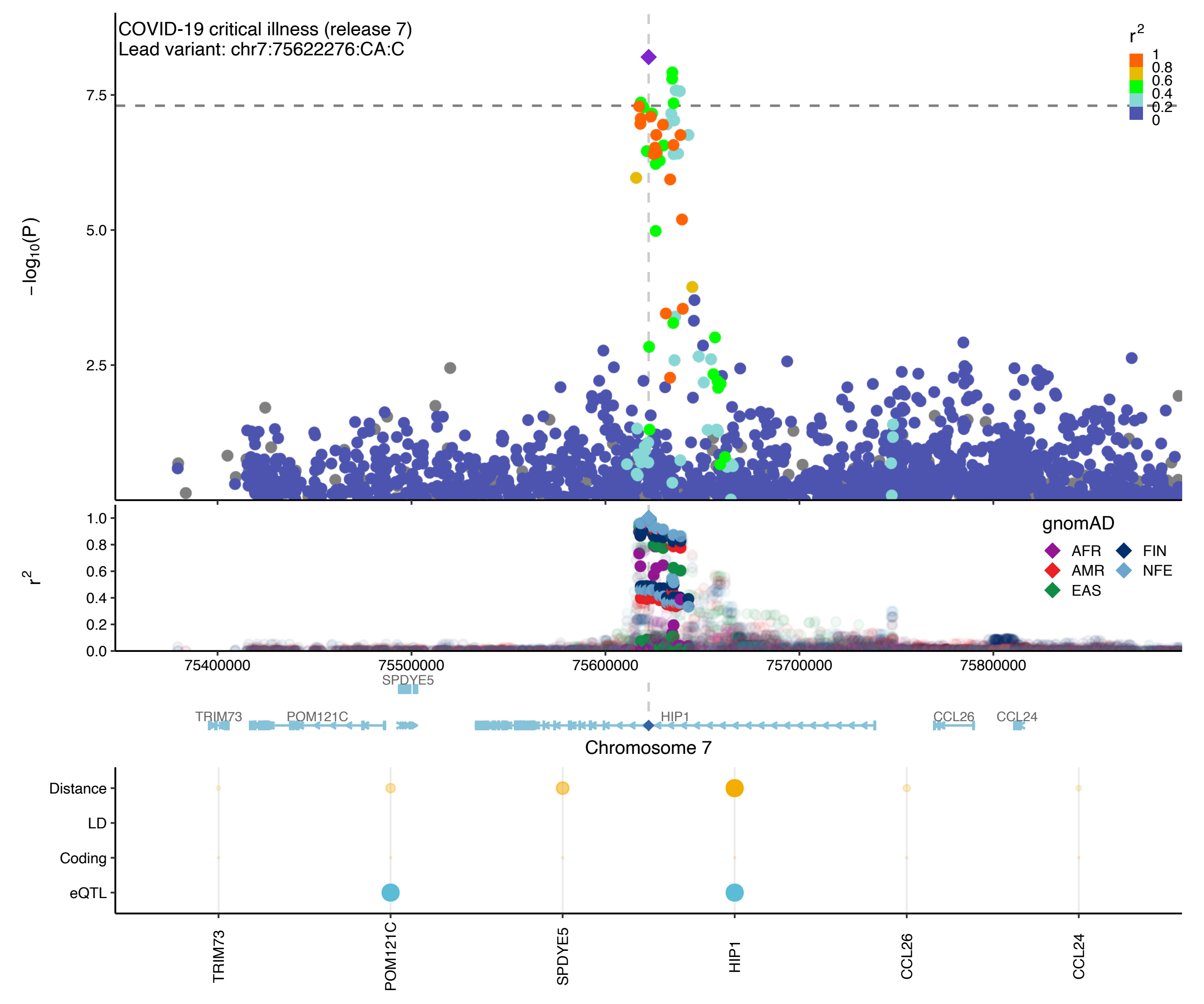
COVID-19 critical illness (release 7)
Lead variant: chr7:75622276:CA:C
r2
1
0.8
0.6
0.4
0.2
0
7.5
5.0
2.5
−log10(P)
1.0
0.8
0.6
0.4
0.2
0.0
r2
gnomAD
AFR
AMR
EAS
FIN
NFE
75400000
75500000
75600000
75700000
75800000
SPDYE5
TRIM73
POM121C
HIP1
CCL26
CCL24
Chromosome 7
Distance
LD
Coding
eQTL
TRIM73
POM121C
SPDYE5
HIP1
CCL26
CCL24

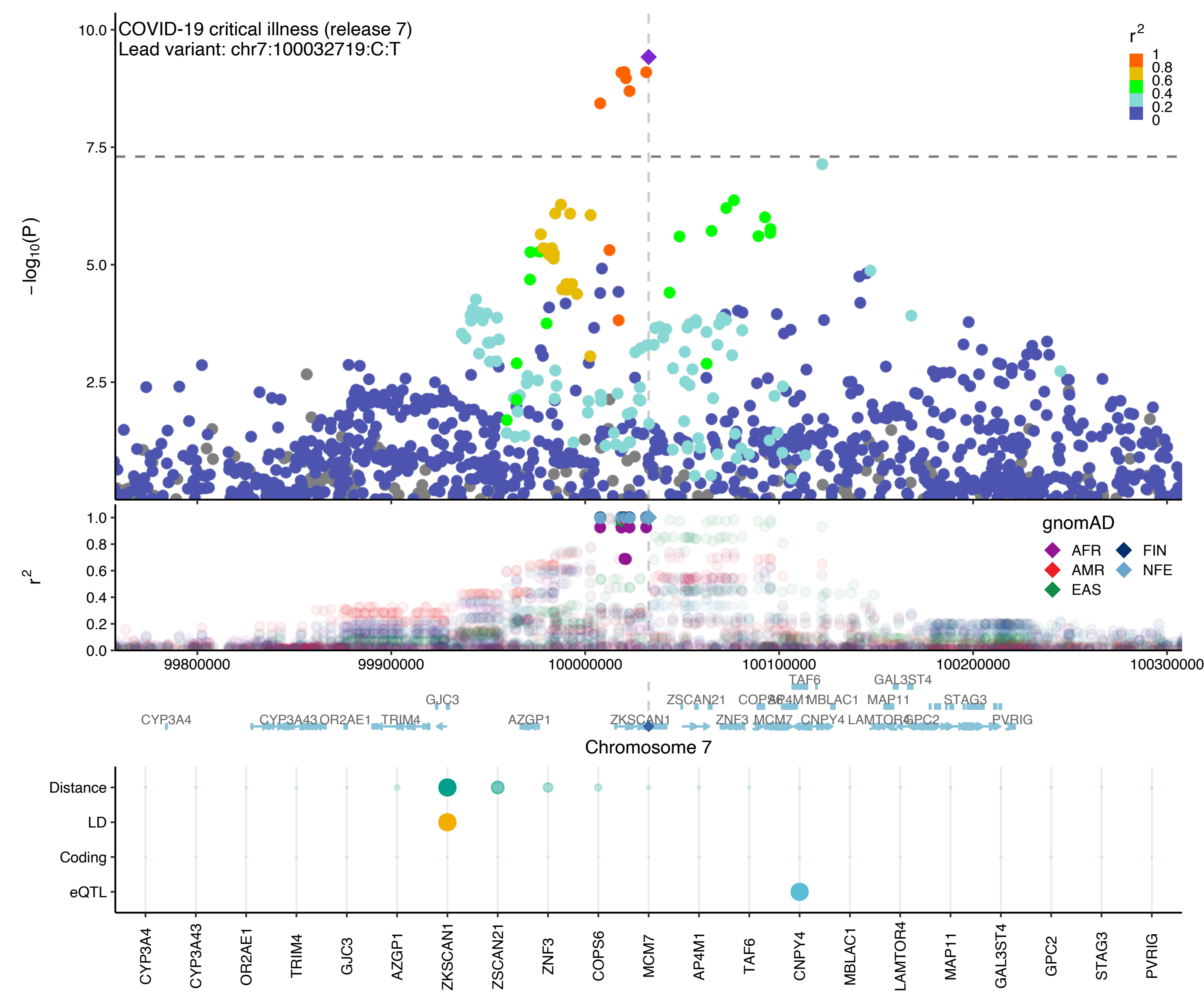

COVID-19 critical illness (release 7)
Lead variant: chr7:100032719:C:T
r2
1
0.8
0.6
0.4
0.2
0
−log10(P)
10.0
7.5
5.0
2.5
r2
1.0
0.8
0.6
0.4
0.2
0.0
gnomAD
AFR
AMR
EAS
FIN
NFE
99800000
99900000
100000000
100100000
100200000
100300000
Chromosome 7
Distance
LD
Coding
eQTL
CYP3A4
CYP3A43
OR2AE1
TRIM4
GJC3
AZGP1
ZKSCAN1
ZSCAN21
ZNF3
COPS6
MCM7
AP4M1
TAF6
CNPY4
MBLAC1
LAMTOR4
MAP11
GAL3ST4
GPC2
STAG3
PVRIG

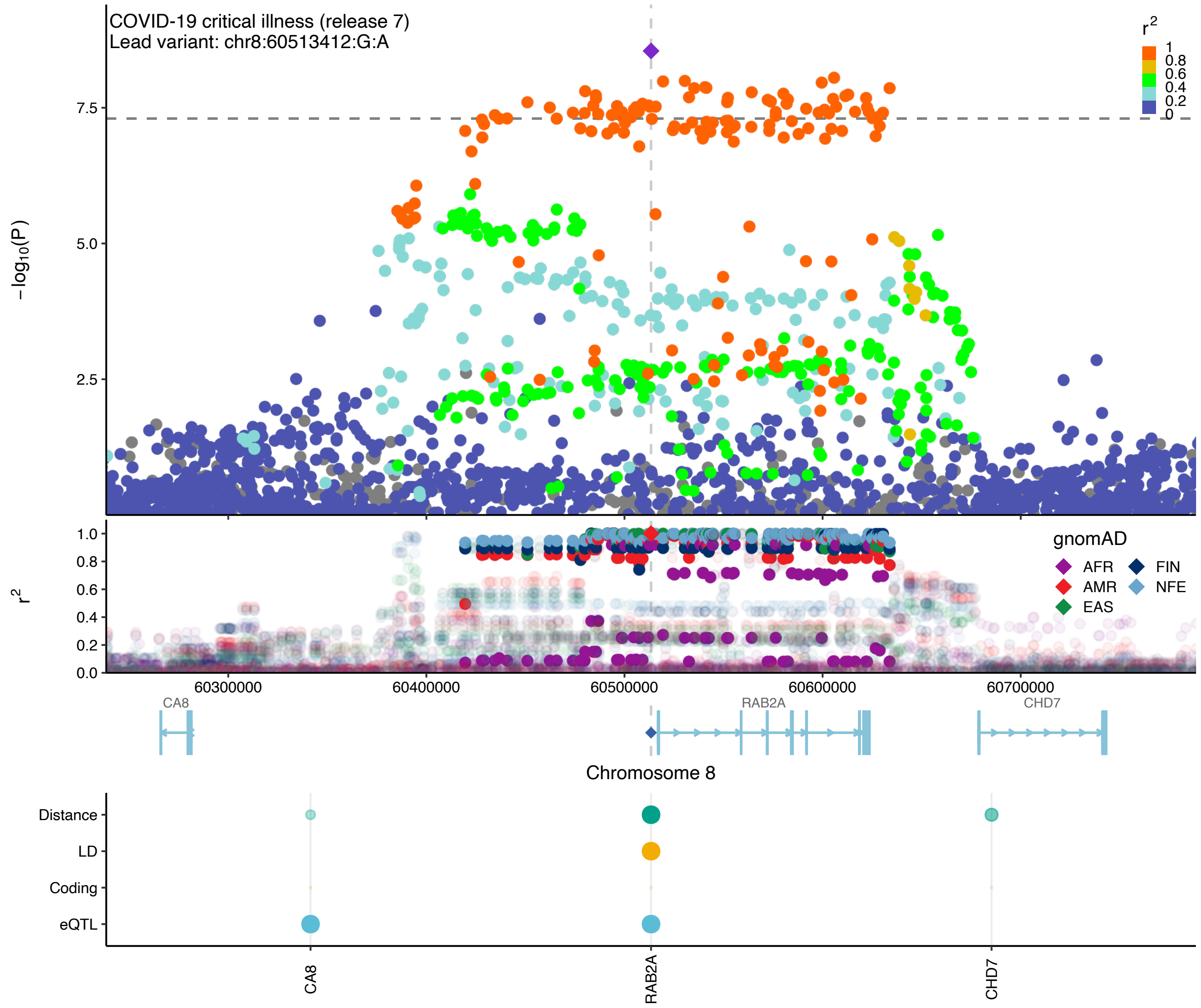

COVID-19 critical illness (release 7)
Lead variant: chr8:60513412:G:A
r²
1
0.8
0.6
0.4
0.2
0
−log10(P)
7.5
5.0
2.5
r²
1.0
0.8
0.6
0.4
0.2
0.0
gnomAD
AFR
AMR
EAS
FIN
NFE
60300000
60400000
60500000
60600000
60700000
CA8
RAB2A
CHD7
Chromosome 8
Distance
LD
Coding
eQTL
CA8
RAB2A
CHD7

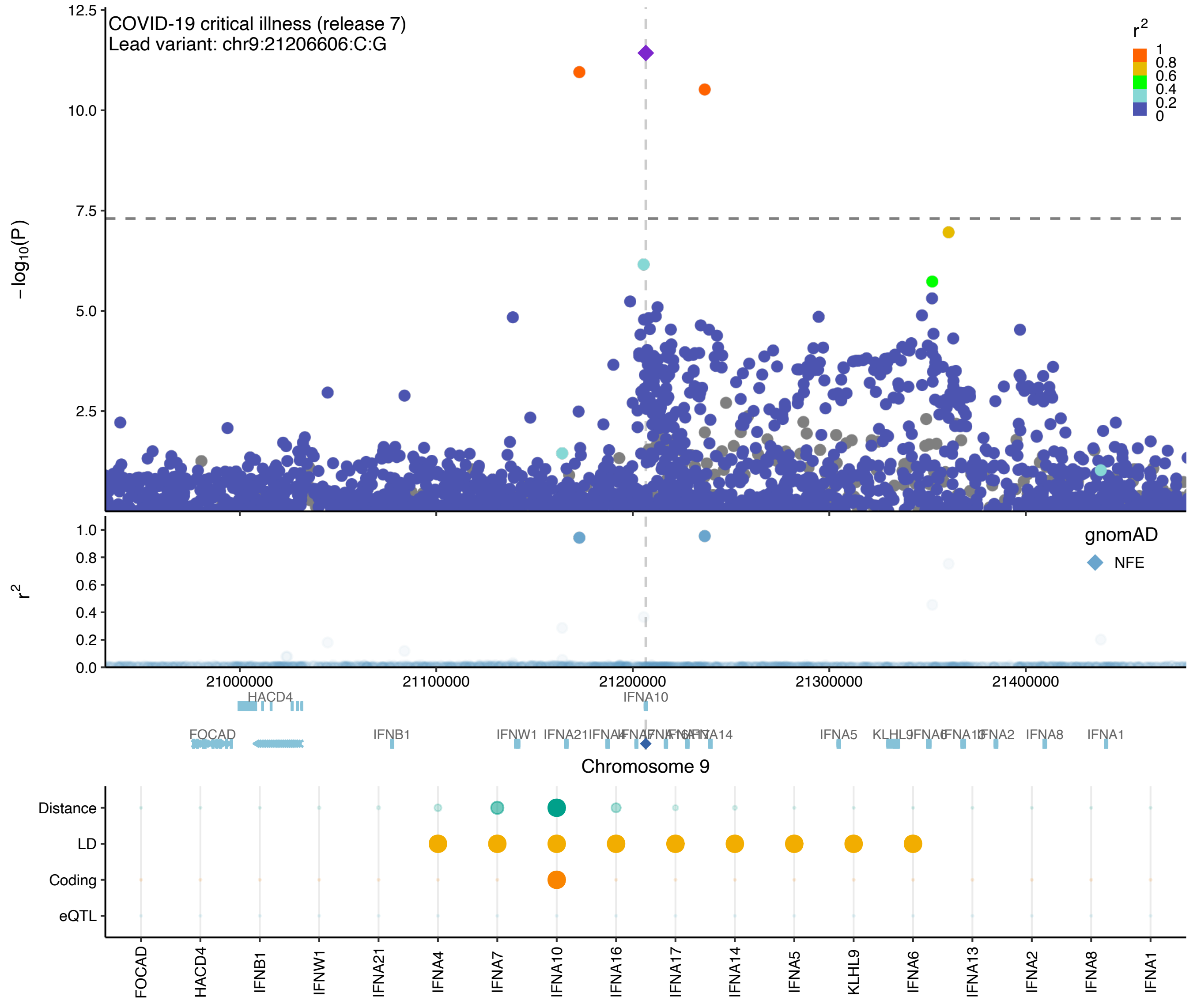
COVID-19 critical illness (release 7)
Lead variant: chr9:21206606:C:G
$-\log_{10}(P)$
$r^2$
gnomAD
NFE
Chromosome 9
HACD4
IFNA10
FOCAD
IFNB1
IFNW1
IFNA21
IFNA5
IFNA8
IFNA1
Distance
LD
Coding
eQTL
FOCAD
HACD4
IFNB1
IFNW1
IFNA21
IFNA4
IFNA7
IFNA10
IFNA16
IFNA17
IFNA14
IFNA5
KLHL9
IFNA6
IFNA13
IFNA2
IFNA8
IFNA1

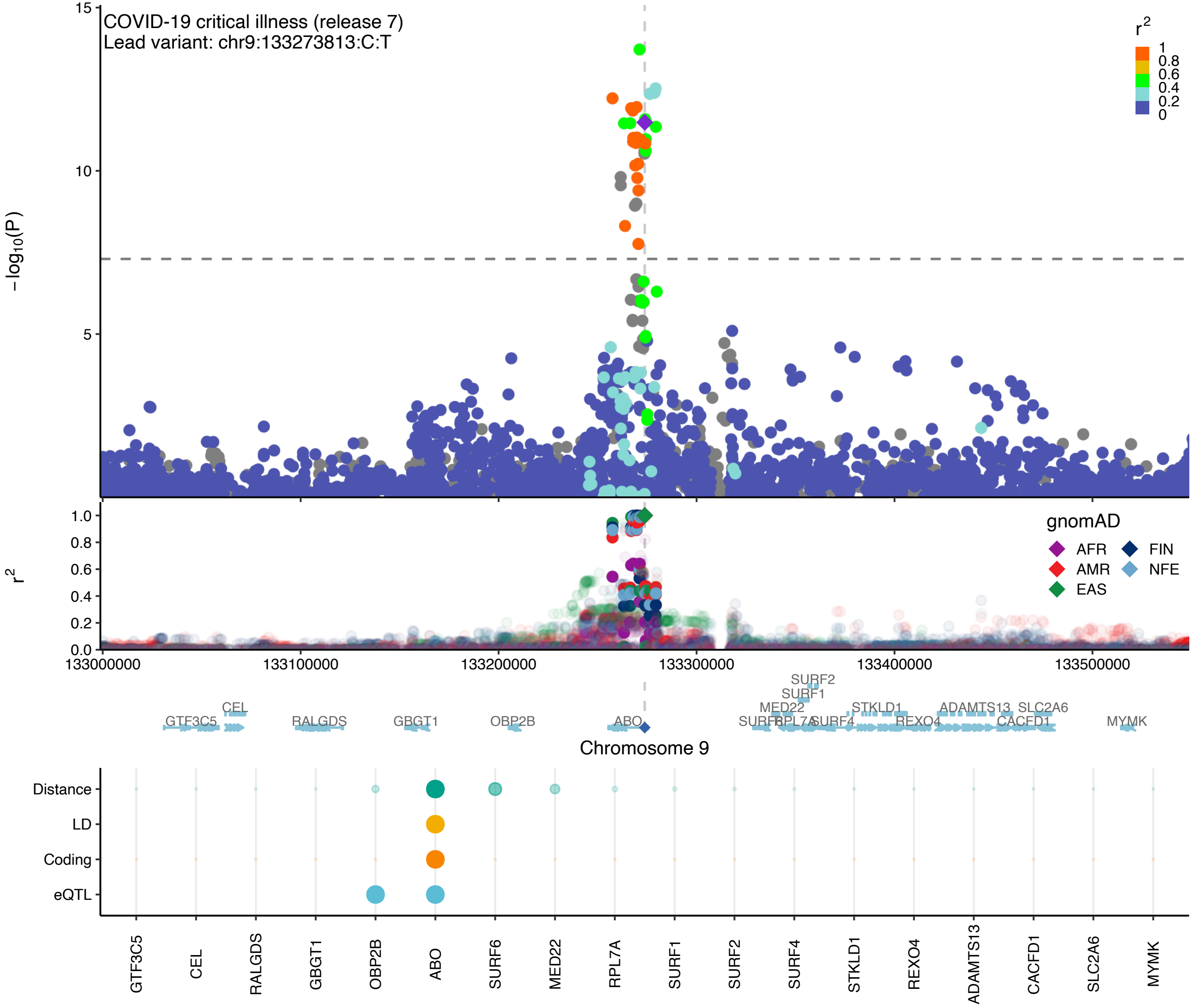
COVID-19 critical illness (release 7)
Lead variant: chr9:133273813:C:T
r2
1
0.8
0.6
0.4
0.2
0
−log10(P)
15
10
5
gnomAD
AFR
AMR
EAS
FIN
NFE
r2
1.0
0.8
0.6
0.4
0.2
0.0
133000000
133100000
133200000
133300000
133400000
133500000
SURF2
SURF1
MED22
STKLD1
ADAMTS13
SLC2A6
CEL
GTF3C5
RALGDS
GBGT1
OBP2B
ABO
SURF6
RPL7A
SURF4
REXO4
CACFD1
MYMK
Chromosome 9
Distance
LD
Coding
eQTL
GTF3C5
CEL
RALGDS
GBGT1
OBP2B
ABO
SURF6
MED22
RPL7A
SURF1
SURF2
SURF4
STKLD1
REXO4
ADAMTS13
CACFD1
SLC2A6
MYMK

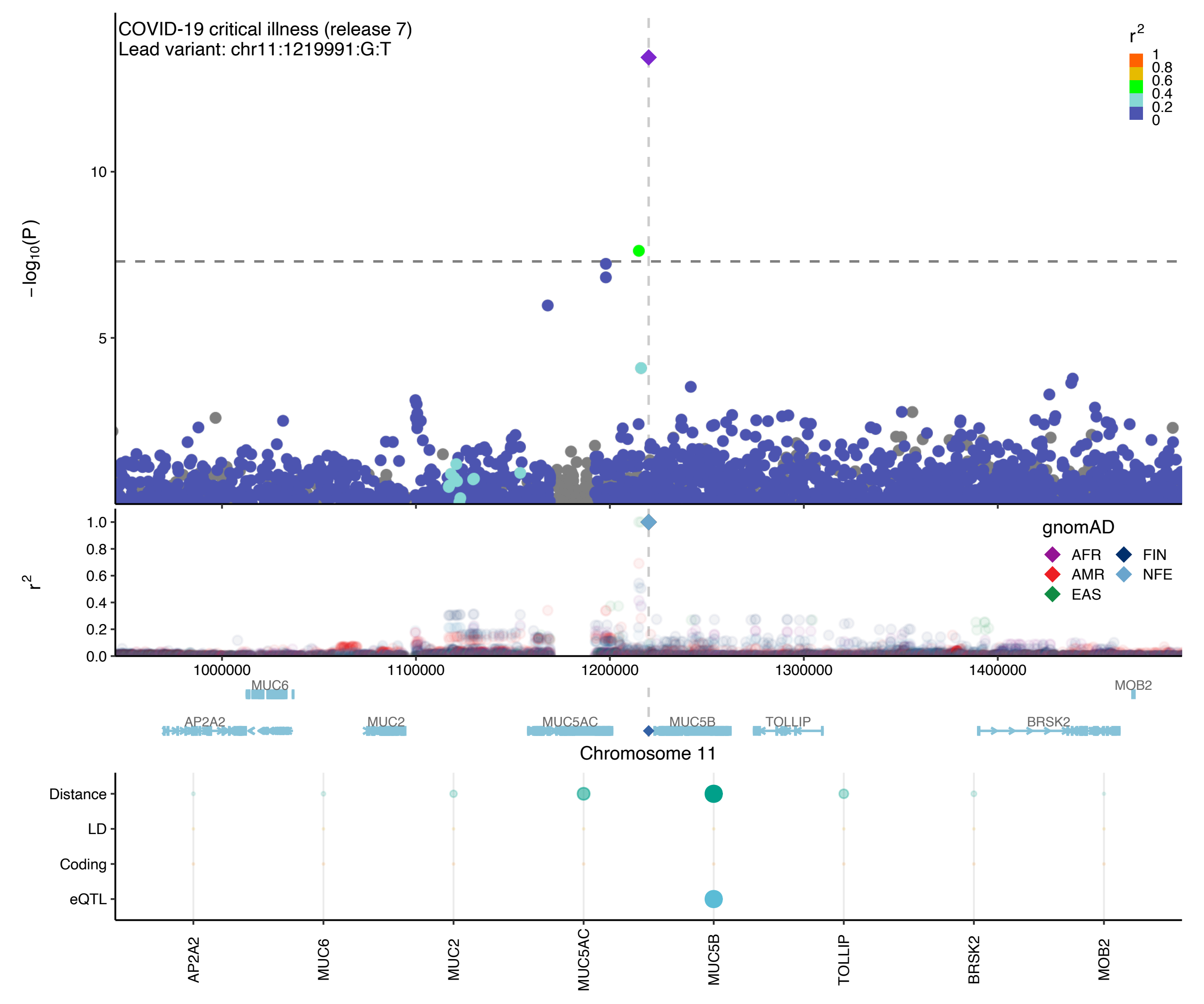

COVID-19 critical illness (release 7)
Lead variant: chr11:1219991:G:T
r2
1
0.8
0.6
0.4
0.2
0
−log10(P)
10
5
r2
1.0
0.8
0.6
0.4
0.2
0.0
gnomAD
AFR
FIN
AMR
NFE
EAS
1000000
1100000
1200000
1300000
1400000
MUC6
MOB2
AP2A2
MUC2
MUC5AC
MUC5B
TOLLIP
BRSK2
Chromosome 11
Distance
LD
Coding
eQTL
AP2A2
MUC6
MUC2
MUC5AC
MUC5B
TOLLIP
BRSK2
MOB2

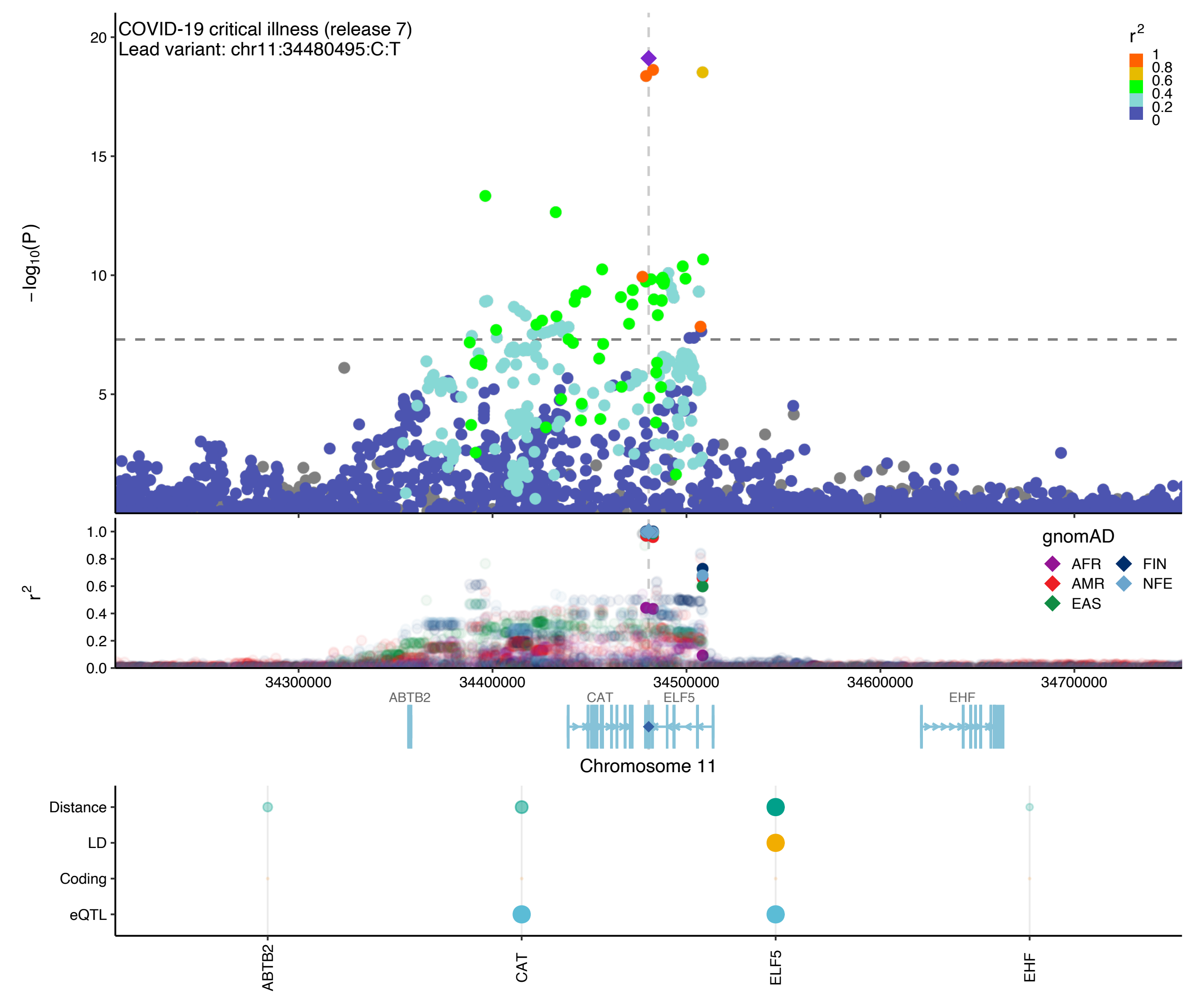
COVID-19 critical illness (release 7)
Lead variant: chr11:34480495:C:T
−log10(P)
20
15
10
5
r2
1
0.8
0.6
0.4
0.2
0
1.0
0.8
0.6
0.4
0.2
0.0
gnomAD
AFR
AMR
EAS
FIN
NFE
34300000
34400000
34500000
34600000
34700000
ABTB2
CAT
ELF5
EHF
Chromosome 11
Distance
LD
Coding
eQTL
ABTB2
CAT
ELF5
EHF

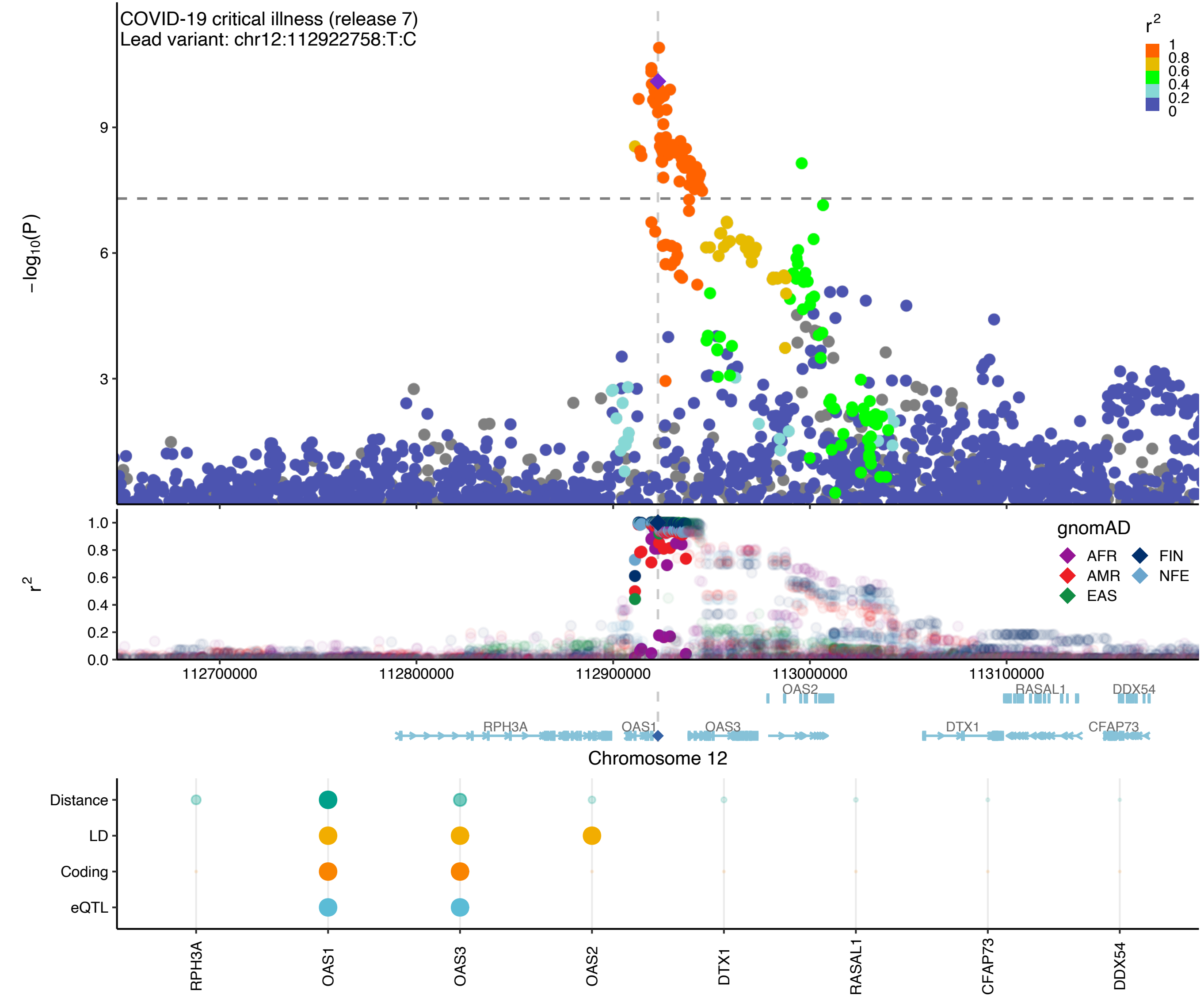
COVID-19 critical illness (release 7)
Lead variant: chr12:112922758:T:C
r²
1
0.8
0.6
0.4
0.2
0
−log10(P)
9
6
3
r²
1.0
0.8
0.6
0.4
0.2
0.0
gnomAD
AFR
FIN
AMR
NFE
EAS
112700000
112800000
112900000
113000000
113100000
OAS2
RASAL1
DDX54
RPH3A
OAS1
OAS3
DTX1
CFAP73
Chromosome 12
Distance
LD
Coding
eQTL
RPH3A
OAS1
OAS3
OAS2
DTX1
RASAL1
CFAP73
DDX54

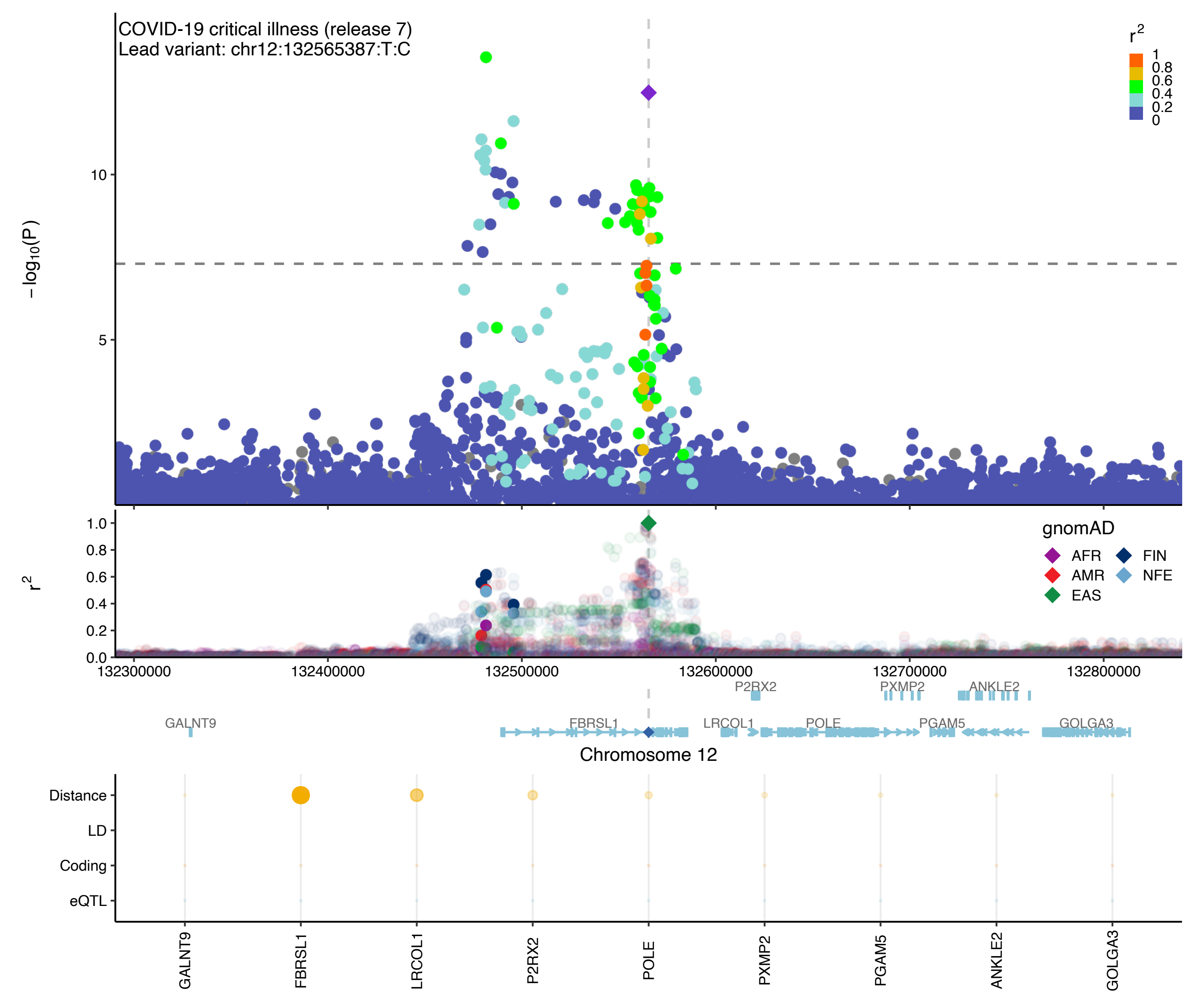

COVID-19 critical illness (release 7)
Lead variant: chr12:132565387:T:C
r2
1
0.8
0.6
0.4
0.2
0
−log10(P)
10
5
r2
1.0
0.8
0.6
0.4
0.2
0.0
gnomAD
AFR
FIN
AMR
NFE
EAS
132300000
132400000
132500000
132600000
132700000
132800000
P2RX2
PXMP2
ANKLE2
GALNT9
FBRSL1
LRCOL1
POLE
PGAM5
GOLGA3
Chromosome 12
Distance
LD
Coding
eQTL
GALNT9
FBRSL1
LRCOL1
P2RX2
POLE
PXMP2
PGAM5
ANKLE2
GOLGA3

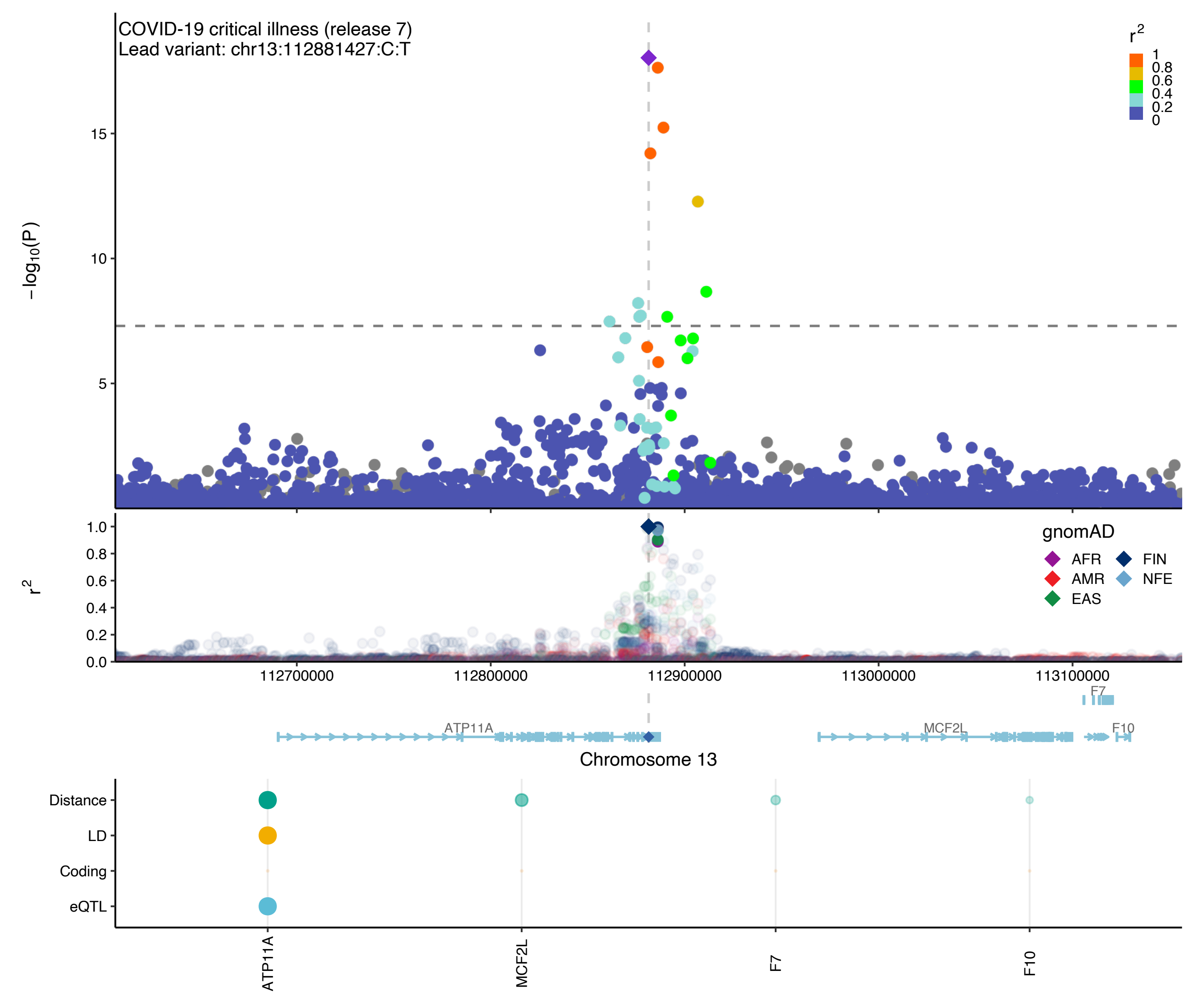

COVID-19 critical illness (release 7)
Lead variant: chr13:112881427:C:T
−log10(P)
15
10
5
r2
1
0.8
0.6
0.4
0.2
0
gnomAD
AFR
AMR
EAS
FIN
NFE
r2
1.0
0.8
0.6
0.4
0.2
0.0
112700000
112800000
112900000
113000000
113100000
F7
ATP11A
MCF2L
F10
Chromosome 13
Distance
LD
Coding
eQTL
ATP11A
MCF2L
F7
F10

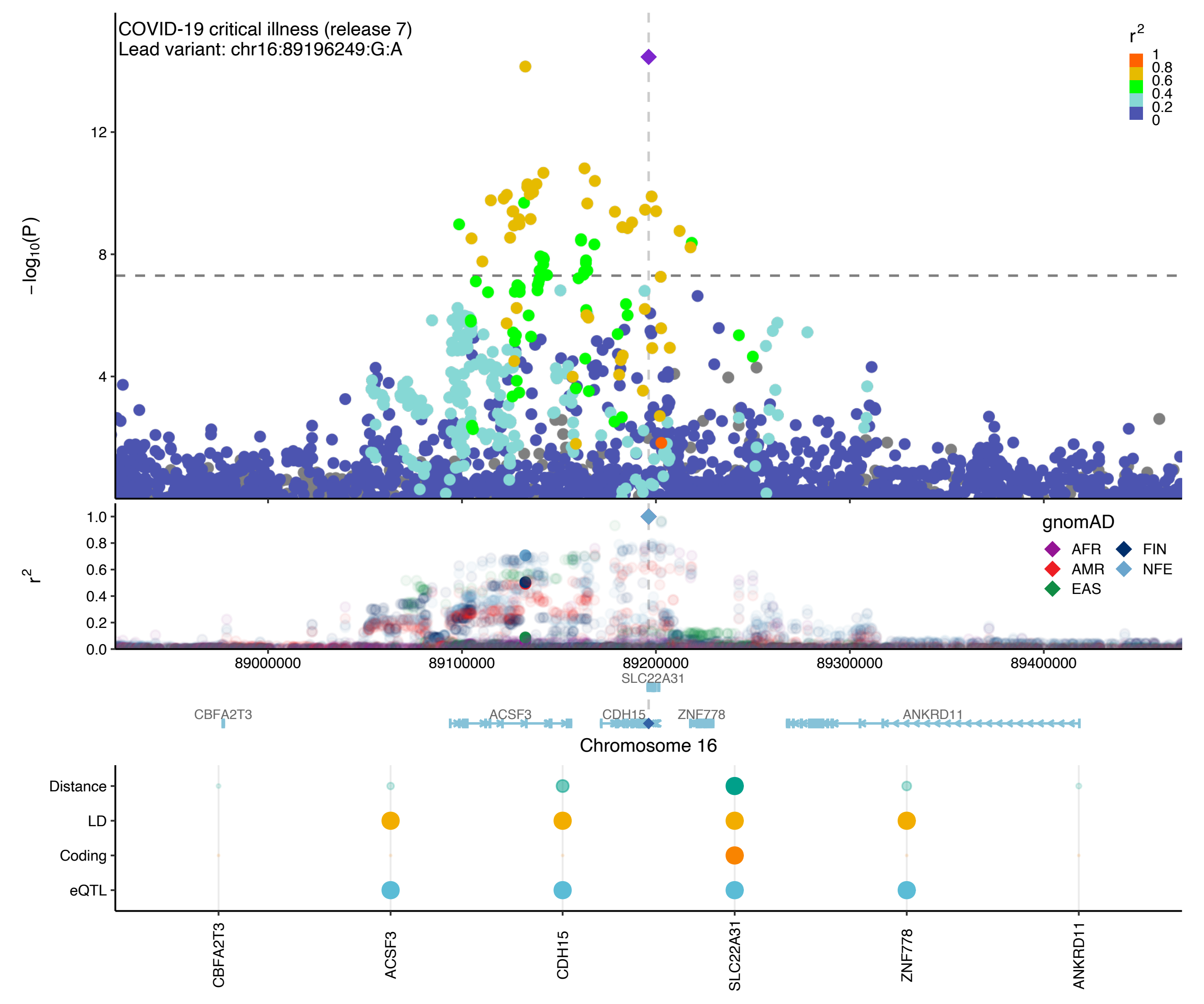

COVID-19 critical illness (release 7)
Lead variant: chr16:89196249:G:A
r2
1
0.8
0.6
0.4
0.2
0
−log10(P)
12
8
4
r2
1.0
0.8
0.6
0.4
0.2
0.0
gnomAD
AFR
AMR
EAS
FIN
NFE
89000000
89100000
89200000
89300000
89400000
SLC22A31
CBFA2T3
ACSF3
CDH15
ZNF778
ANKRD11
Chromosome 16
Distance
LD
Coding
eQTL
CBFA2T3
ACSF3
CDH15
SLC22A31
ZNF778
ANKRD11

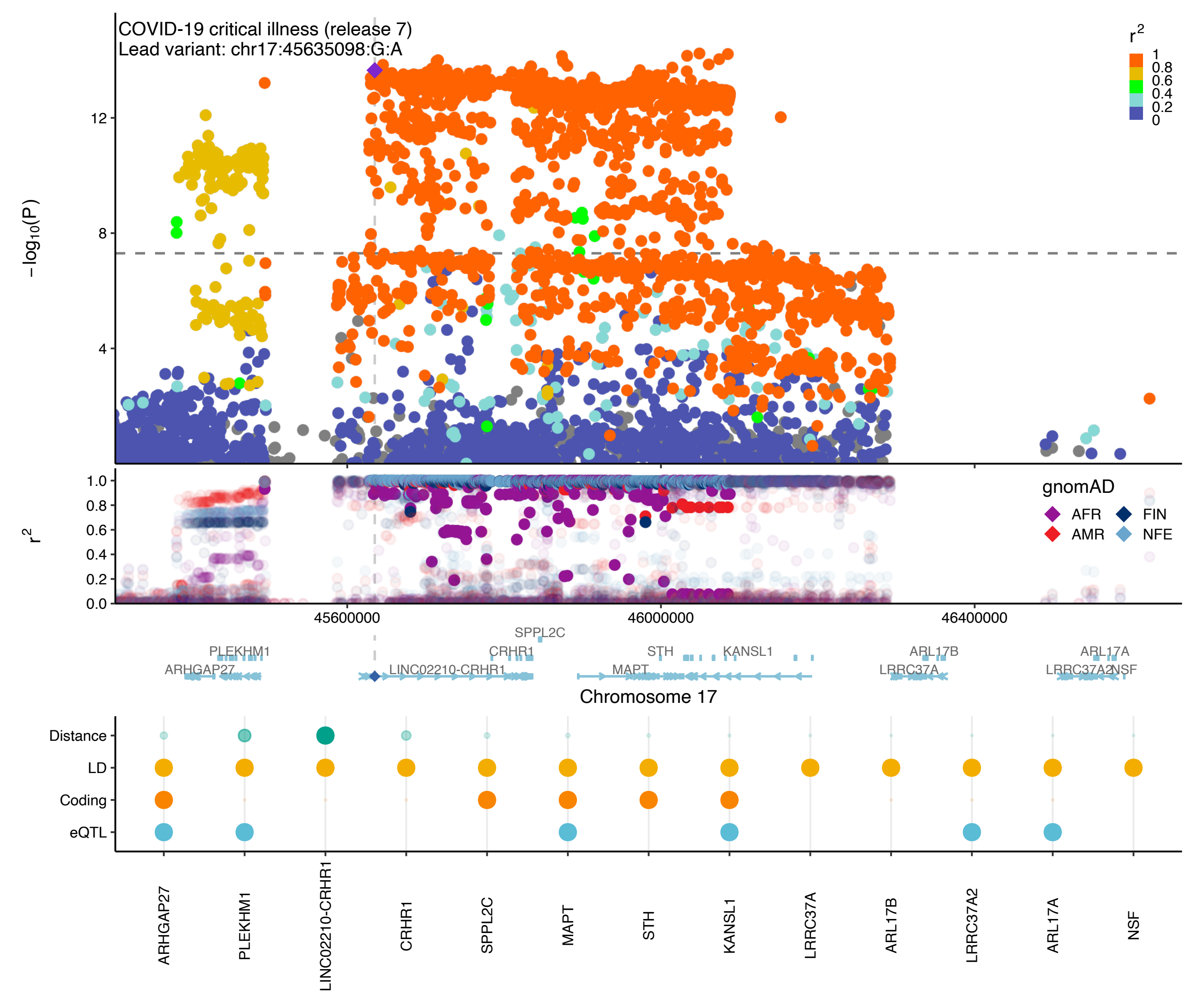

COVID-19 critical illness (release 7)
Lead variant: chr17:45635098:G:A
−log10(P)
12
8
4
r2
1
0.8
0.6
0.4
0.2
0
1.0
0.8
0.6
0.4
0.2
0.0
gnomAD
AFR
FIN
AMR
NFE
45600000
46000000
46400000
SPPL2C
PLEKHM1
CRHR1
STH
KANSL1
ARL17B
ARL17A
ARHGAP27
LINC02210-CRHR1
MAPT
LRRC37A
LRRC37A2
NSF
Chromosome 17
Distance
LD
Coding
eQTL
ARHGAP27
PLEKHM1
LINC02210-CRHR1
CRHR1
SPPL2C
MAPT
STH
KANSL1
LRRC37A
ARL17B
LRRC37A2
ARL17A
NSF

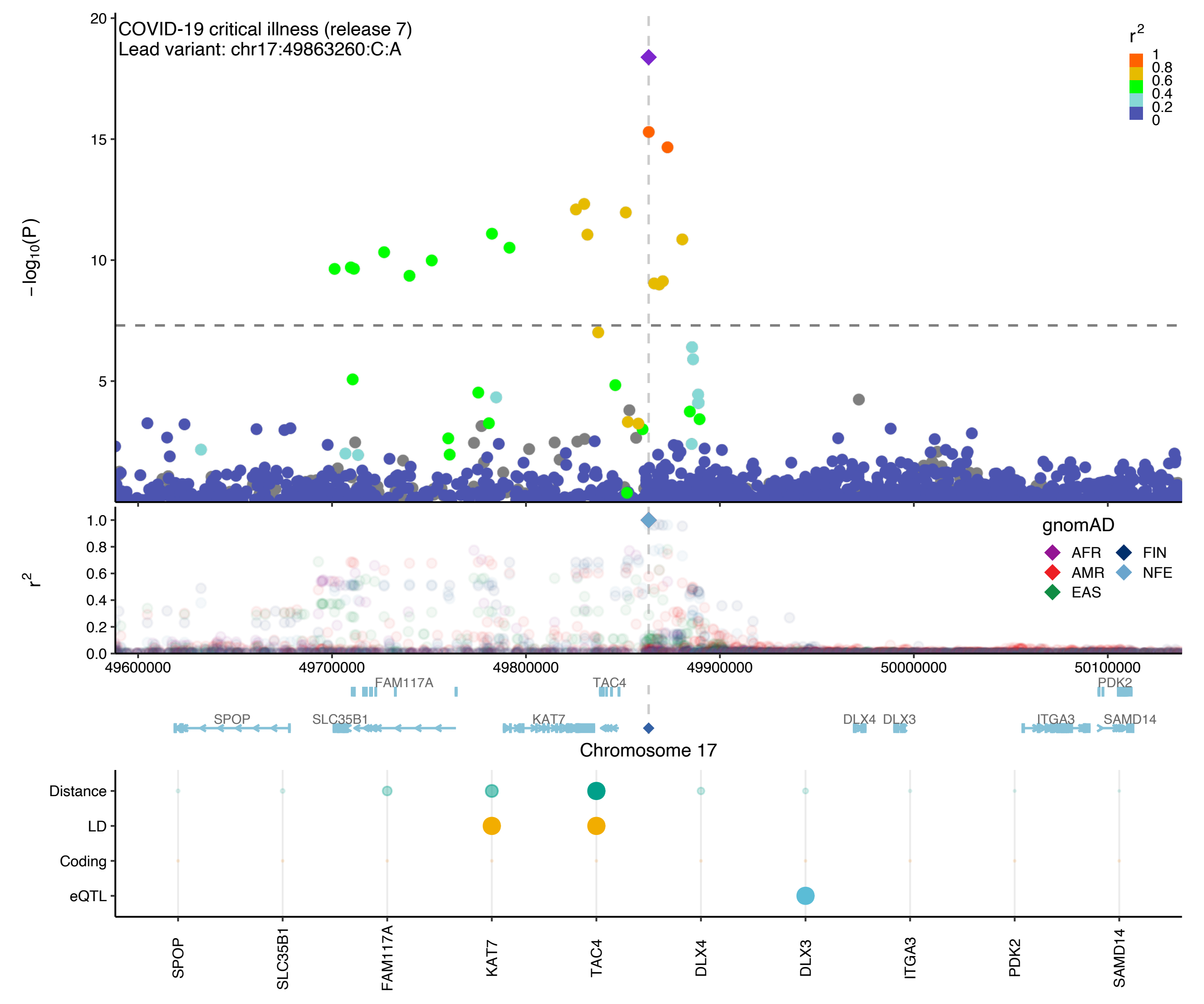

COVID-19 critical illness (release 7)
Lead variant: chr17:49863260:C:A
−log10(P)
r2
1
0.8
0.6
0.4
0.2
0
gnomAD
AFR
AMR
EAS
FIN
NFE
49600000
49700000
49800000
49900000
50000000
50100000
FAM117A
TAC4
PDK2
SPOP
SLC35B1
KAT7
DLX4
DLX3
ITGA3
SAMD14
Chromosome 17
Distance
LD
Coding
eQTL
SPOP
SLC35B1
FAM117A
KAT7
TAC4
DLX4
DLX3
ITGA3
PDK2
SAMD14

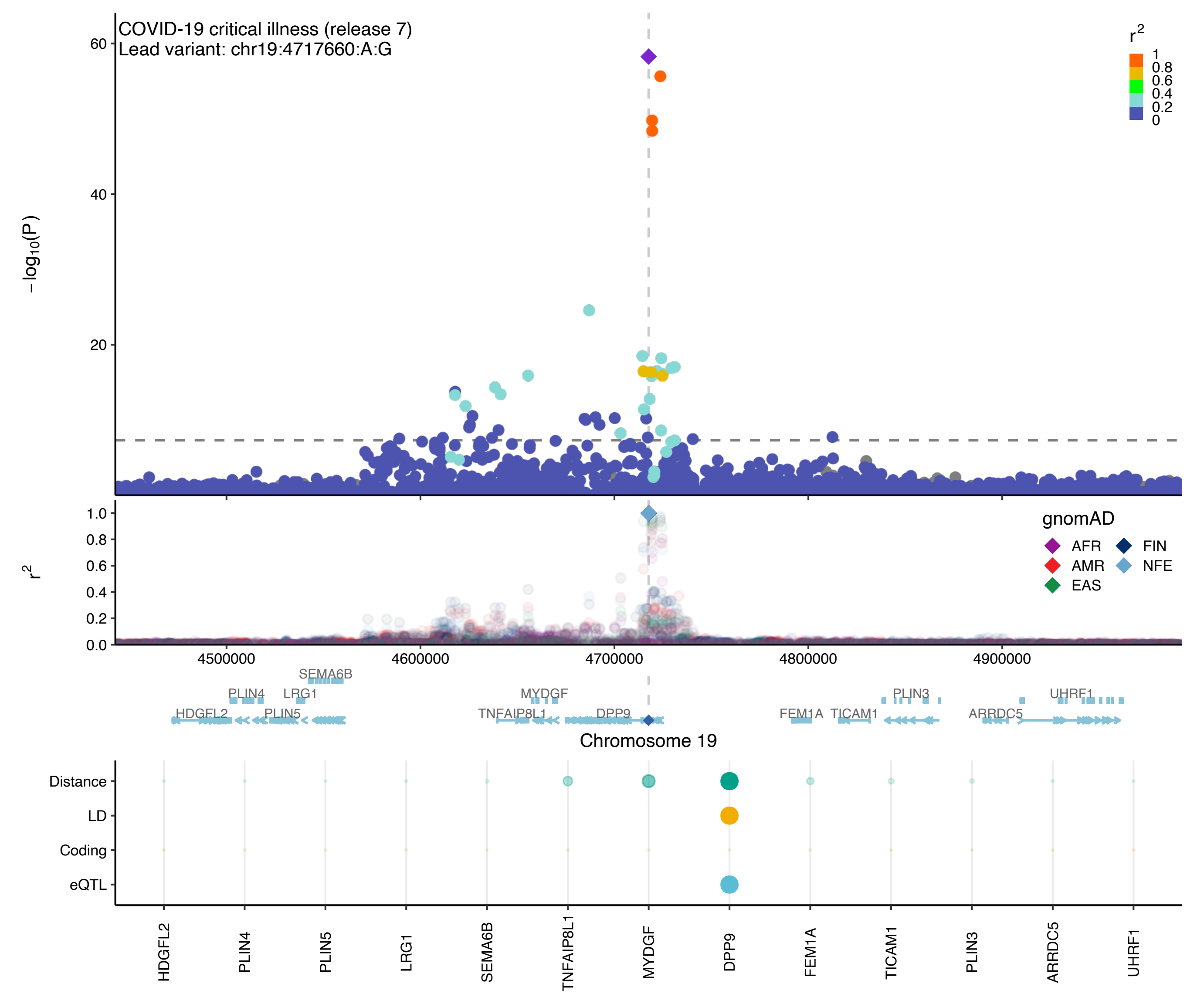

COVID-19 critical illness (release 7)
Lead variant: chr19:4717660:A:G
−log10(P)
r2
1
0.8
0.6
0.4
0.2
0
gnomAD
AFR
AMR
EAS
FIN
NFE
4500000
4600000
4700000
4800000
4900000
SEMA6B
PLIN4
LRG1
MYDGF
PLIN3
UHRF1
HDGFL2
PLIN5
TNFAIP8L1
DPP9
FEM1A
TICAM1
ARRDC5
Chromosome 19
Distance
LD
Coding
eQTL
HDGFL2
PLIN4
PLIN5
LRG1
SEMA6B
TNFAIP8L1
MYDGF
DPP9
FEM1A
TICAM1
PLIN3
ARRDC5
UHRF1

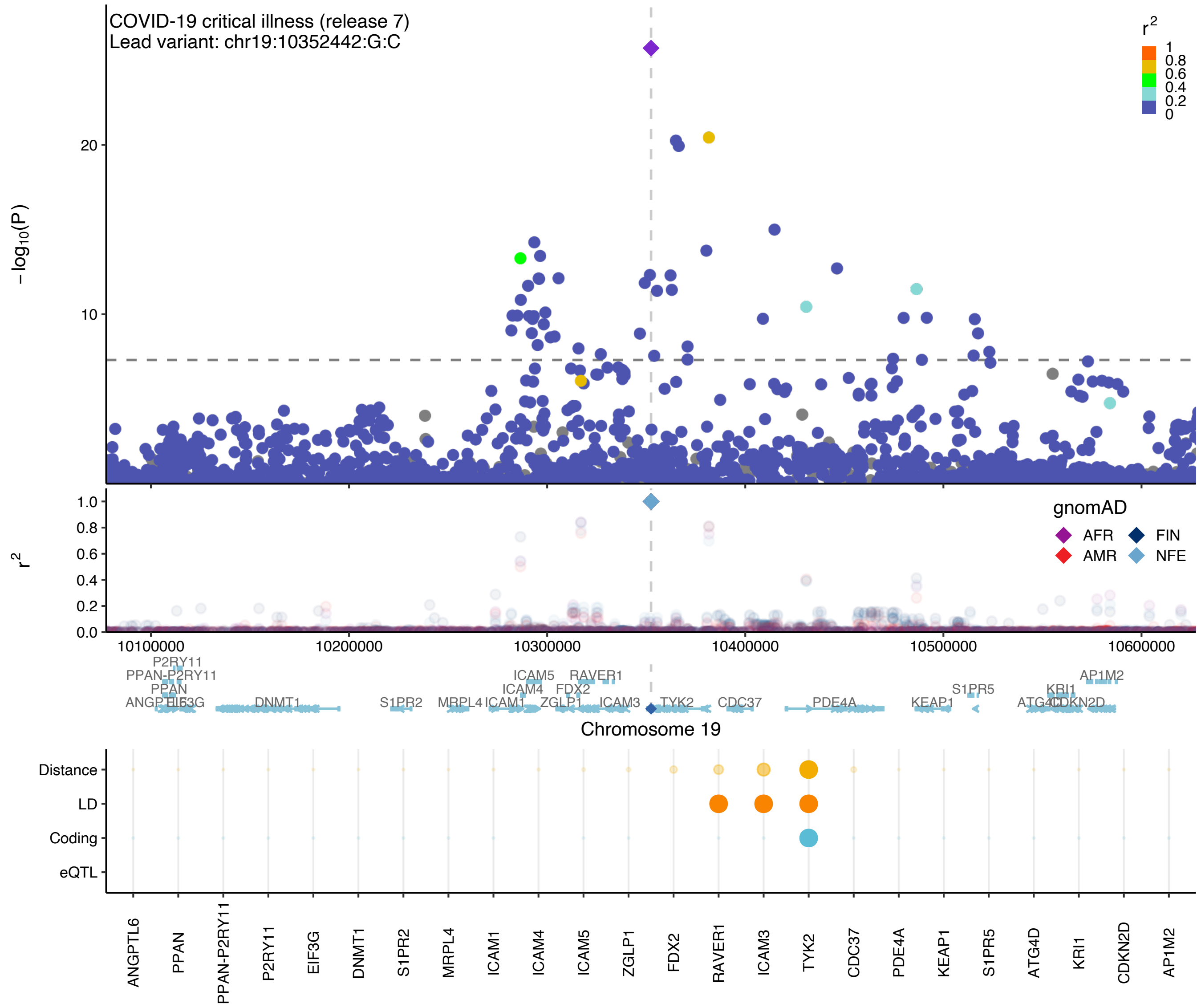

COVID-19 critical illness (release 7)
Lead variant: chr19:10352442:G:C
r2
1
0.8
0.6
0.4
0.2
0
−log10(P)
20
10
r2
1.0
0.8
0.6
0.4
0.2
0.0
gnomAD
AFR
FIN
AMR
NFE
10100000
10200000
10300000
10400000
10500000
10600000
P2RY11
PPAN-P2RY11
PPAN
ANGPTL6
EIF3G
DNMT1
S1PR2
MRPL4
ICAM1
ICAM4
ICAM5
ZGLP1
FDX2
RAVER1
ICAM3
TYK2
CDC37
PDE4A
KEAP1
S1PR5
ATG4D
KRI1
CDKN2D
AP1M2
Chromosome 19
Distance
LD
Coding
eQTL
ANGPTL6
PPAN
PPAN-P2RY11
P2RY11
EIF3G
DNMT1
S1PR2
MRPL4
ICAM1
ICAM4
ICAM5
ZGLP1
FDX2
RAVER1
ICAM3
TYK2
CDC37
PDE4A
KEAP1
S1PR5
ATG4D
KRI1
CDKN2D
AP1M2

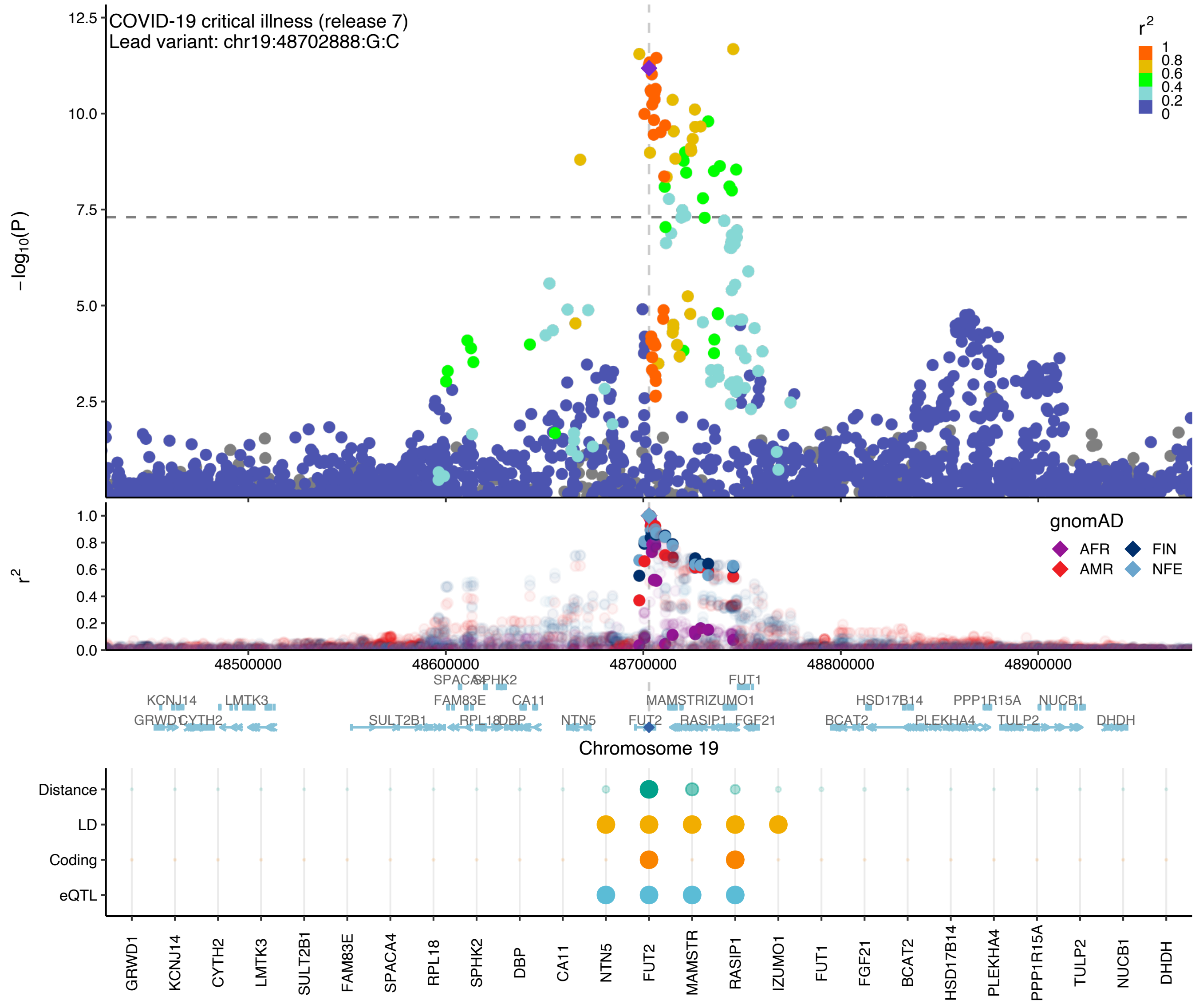

COVID-19 critical illness (release 7)
Lead variant: chr19:48702888:G:C
−log10(P)
r2
gnomAD
AFR
FIN
AMR
NFE
Chromosome 19
48500000
48600000
48700000
48800000
48900000
Distance
LD
Coding
eQTL
GRWD1
KCNJ14
CYTH2
LMTK3
SULT2B1
FAM83E
SPACA4
RPL18
SPHK2
DBP
CA11
NTN5
FUT2
MAMSTR
RASIP1
IZUMO1
FUT1
FGF21
BCAT2
HSD17B14
PLEKHA4
PPP1R15A
TULP2
NUCB1
DHDH

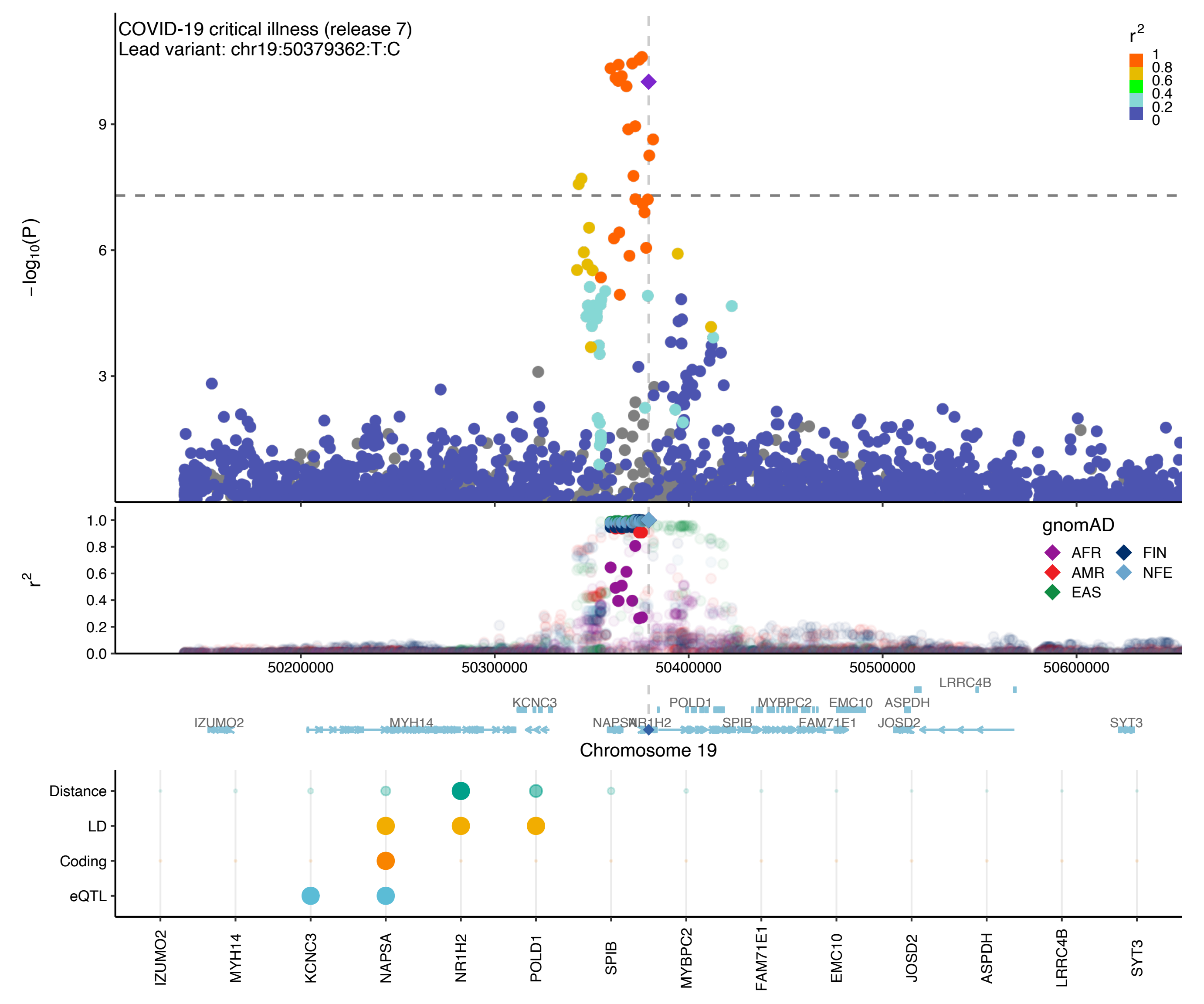

COVID-19 critical illness (release 7)
Lead variant: chr19:50379362:T:C
r2
1
0.8
0.6
0.4
0.2
0
−log10(P)
9
6
3
r2
1.0
0.8
0.6
0.4
0.2
0.0
gnomAD
AFR
AMR
EAS
FIN
NFE
50200000
50300000
50400000
50500000
50600000
LRRC4B
KCNC3
POLD1
MYBPC2
EMC10
ASPDH
IZUMO2
MYH14
NAPSA
NR1H2
SPIB
FAM71E1
JOSD2
SYT3
Chromosome 19
Distance
LD
Coding
eQTL
IZUMO2
MYH14
KCNC3
NAPSA
NR1H2
POLD1
SPIB
MYBPC2
FAM71E1
EMC10
JOSD2
ASPDH
LRRC4B
SYT3

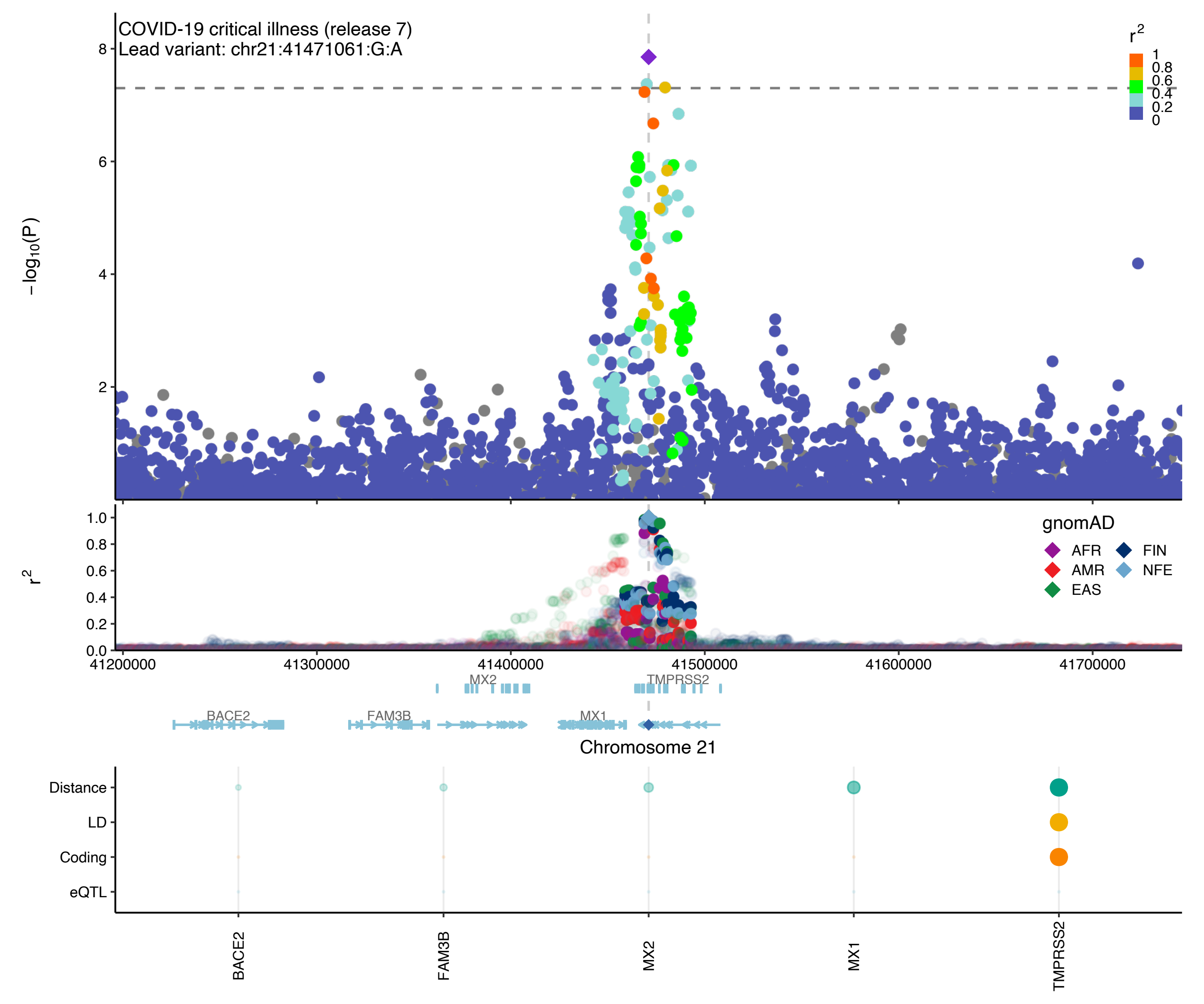

COVID-19 critical illness (release 7)
Lead variant: chr21:41471061:G:A
−log10(P)
r2
1
0.8
0.6
0.4
0.2
0
gnomAD
AFR
AMR
EAS
FIN
NFE
41200000
41300000
41400000
41500000
41600000
41700000
MX2
TMPRSS2
BACE2
FAM3B
MX1
Chromosome 21
Distance
LD
Coding
eQTL
BACE2
FAM3B
MX2
MX1
TMPRSS2

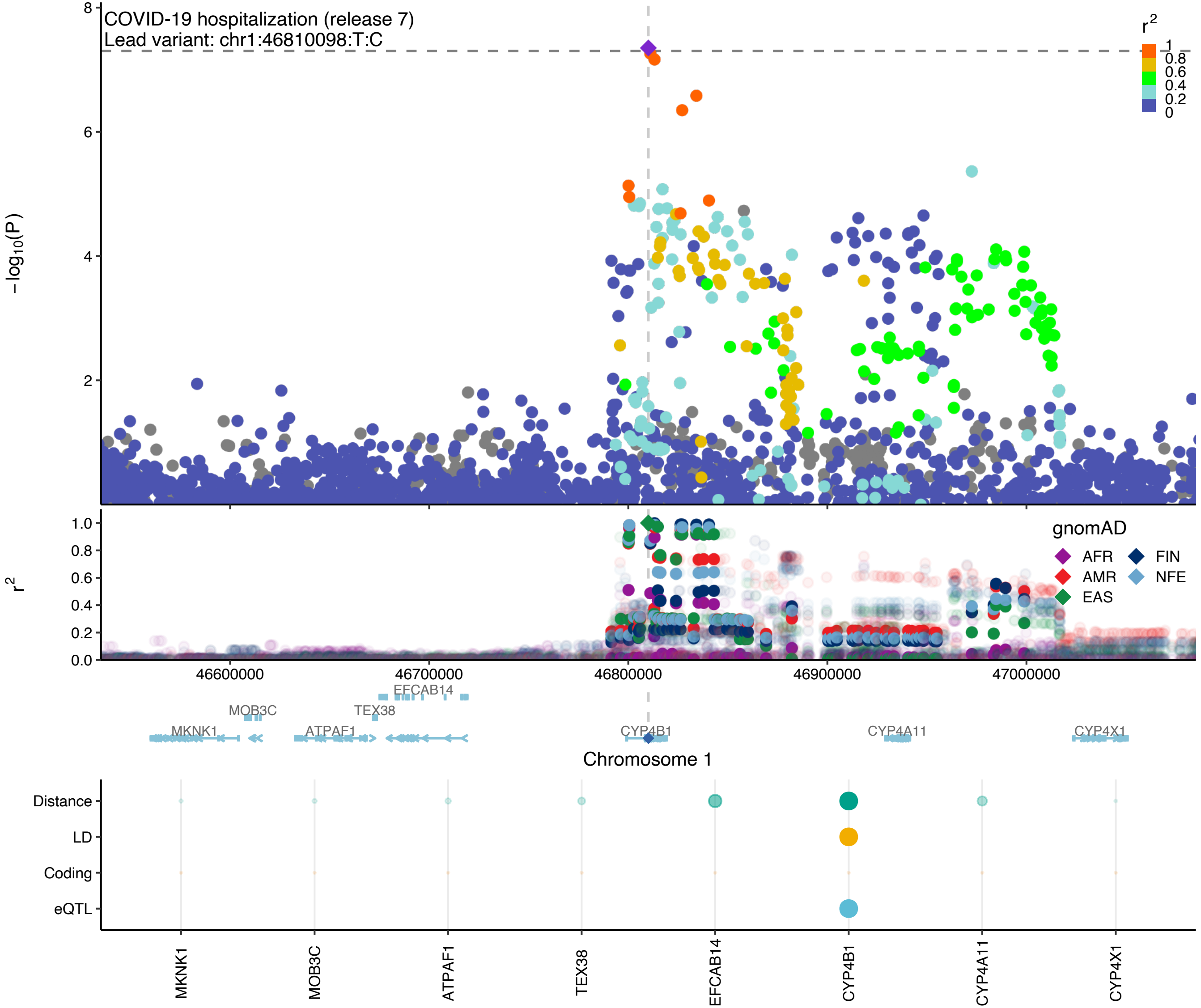

COVID-19 hospitalization (release 7)
Lead variant: chr1:46810098:T:C
−log10(P)
r2
1
0.8
0.6
0.4
0.2
0
gnomAD
AFR
AMR
EAS
FIN
NFE
46600000
46700000
46800000
46900000
47000000
EFCAB14
MOB3C
TEX38
MKNK1
ATPAF1
CYP4B1
CYP4A11
CYP4X1
Chromosome 1
Distance
LD
Coding
eQTL
MKNK1
MOB3C
ATPAF1
TEX38
EFCAB14
CYP4B1
CYP4A11
CYP4X1

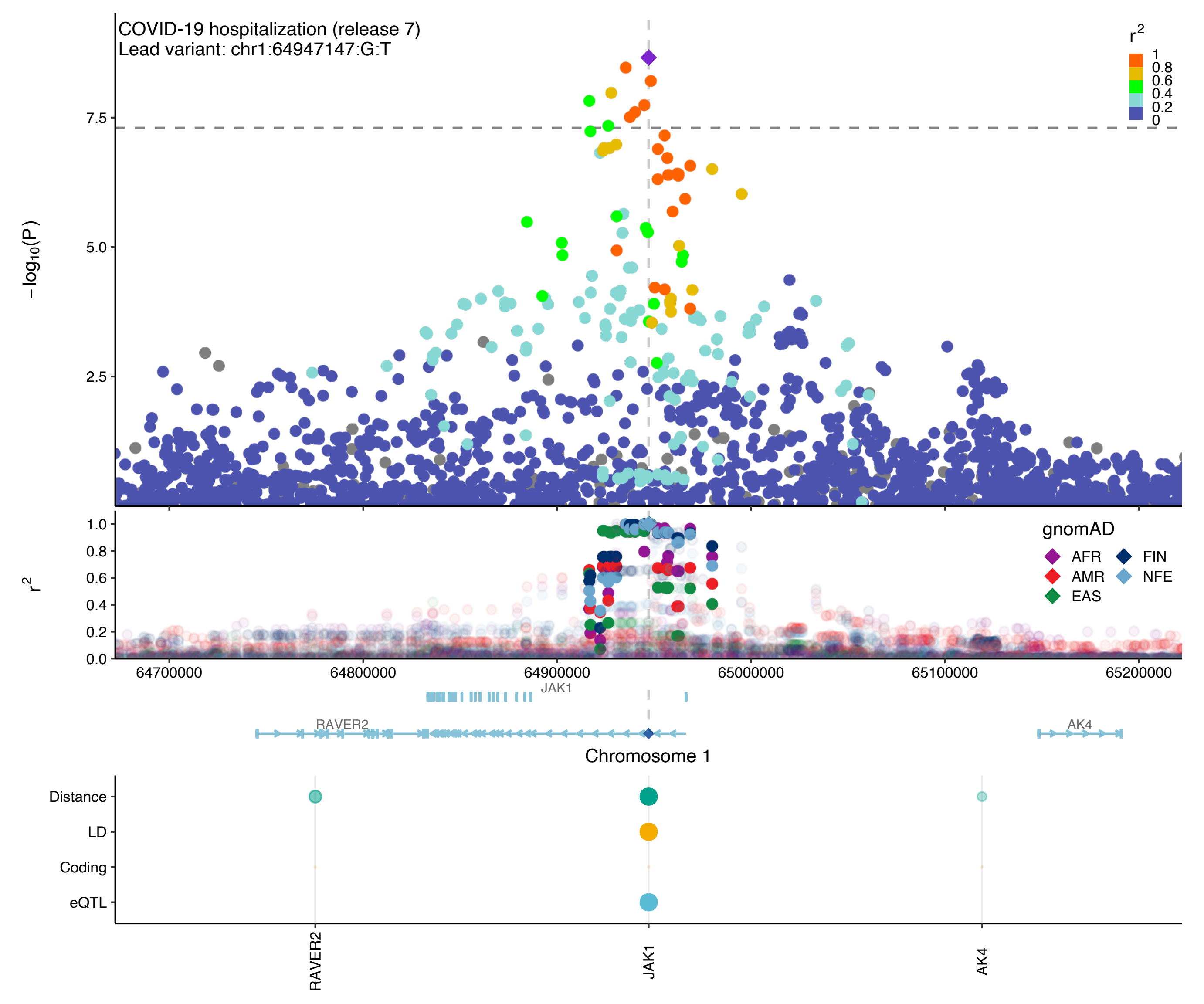

COVID-19 hospitalization (release 7)
Lead variant: chr1:64947147:G:T
r2
1
0.8
0.6
0.4
0.2
0
7.5
5.0
2.5
– log10(P)
1.0
0.8
0.6
0.4
0.2
0.0
r2
gnomAD
AFR
FIN
AMR
NFE
EAS
64700000
64800000
64900000
65000000
65100000
65200000
JAK1
RAVER2
AK4
Chromosome 1
Distance
LD
Coding
eQTL
RAVER2
JAK1
AK4

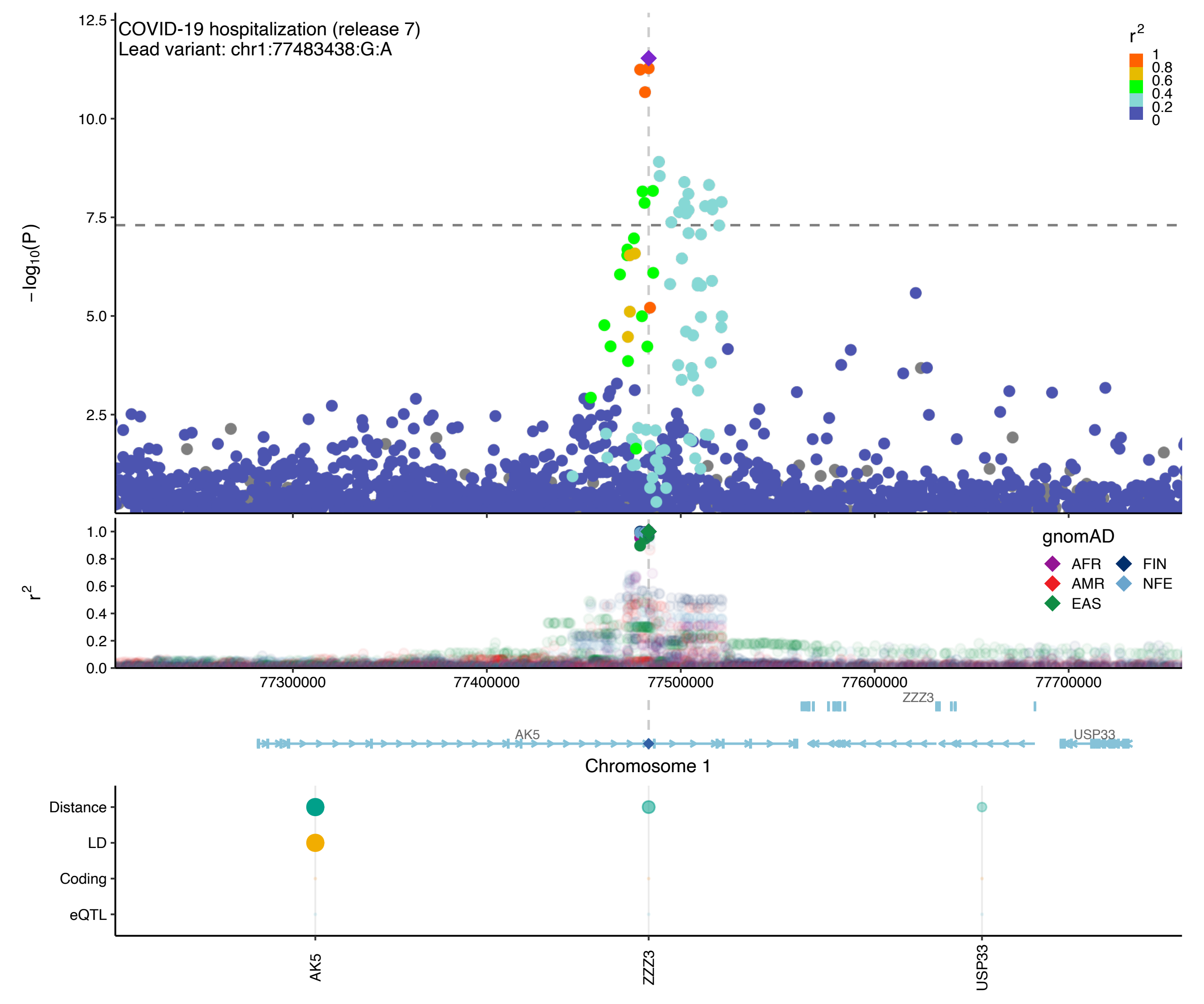

COVID-19 hospitalization (release 7)
Lead variant: chr1:77483438:G:A
−log10(P)
12.5
10.0
7.5
5.0
2.5
r2
1
0.8
0.6
0.4
0.2
0
1.0
0.8
0.6
0.4
0.2
0.0
gnomAD
AFR
AMR
EAS
FIN
NFE
77300000
77400000
77500000
77600000
77700000
ZZZ3
AK5
USP33
Chromosome 1
Distance
LD
Coding
eQTL
AK5
ZZZ3
USP33

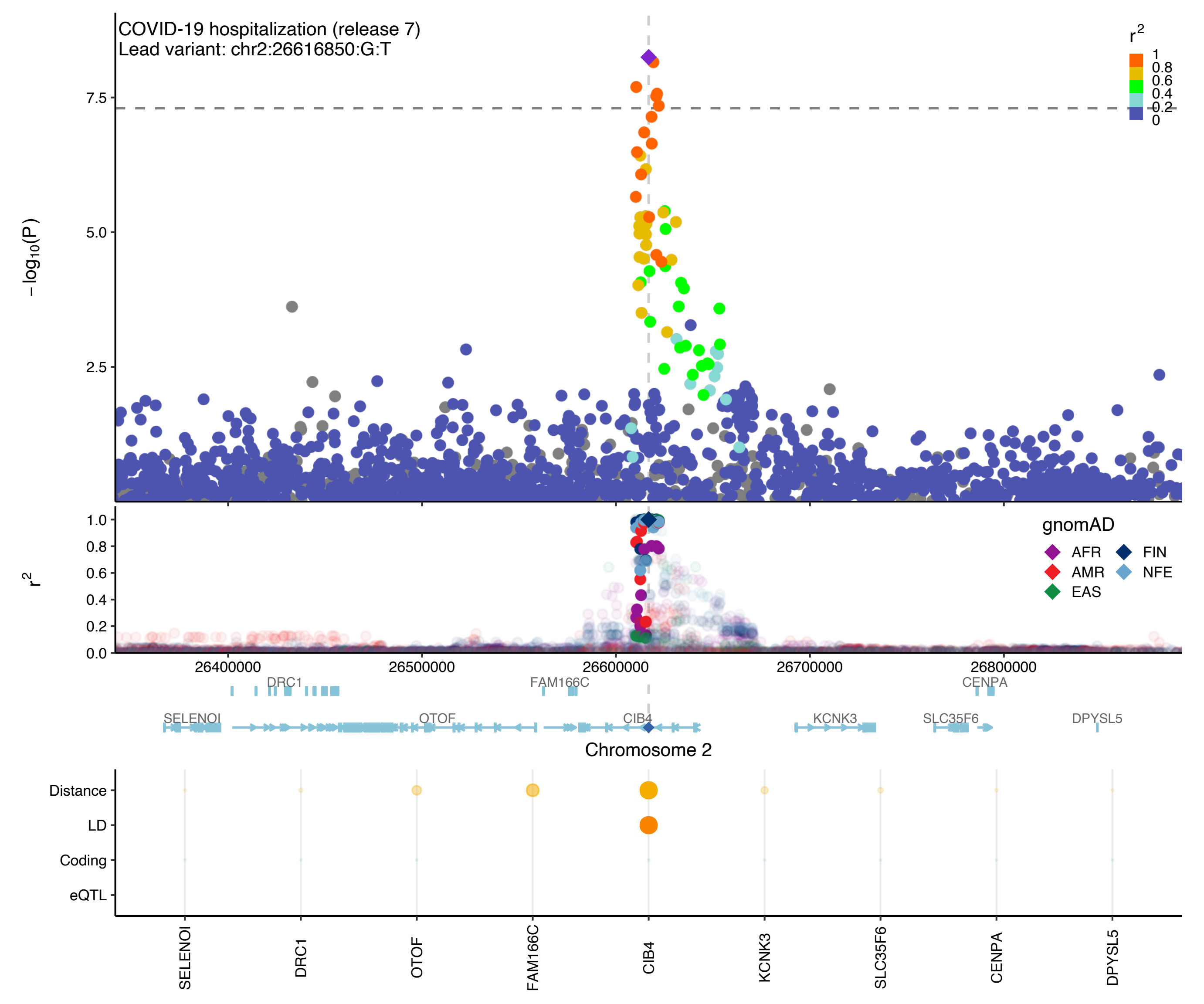
COVID-19 hospitalization (release 7)
Lead variant: chr2:26616850:G:T
r2
1
0.8
0.6
0.4
0.2
0
−log10(P)
7.5
5.0
2.5
r2
1.0
0.8
0.6
0.4
0.2
0.0
gnomAD
AFR
FIN
AMR
NFE
EAS
26400000
26500000
26600000
26700000
26800000
DRC1
FAM166C
CENPA
SELENOI
OTOF
CIB4
KCNK3
SLC35F6
DPYSL5
Chromosome 2
Distance
LD
Coding
eQTL
SELENOI
DRC1
OTOF
FAM166C
CIB4
KCNK3
SLC35F6
CENPA
DPYSL5

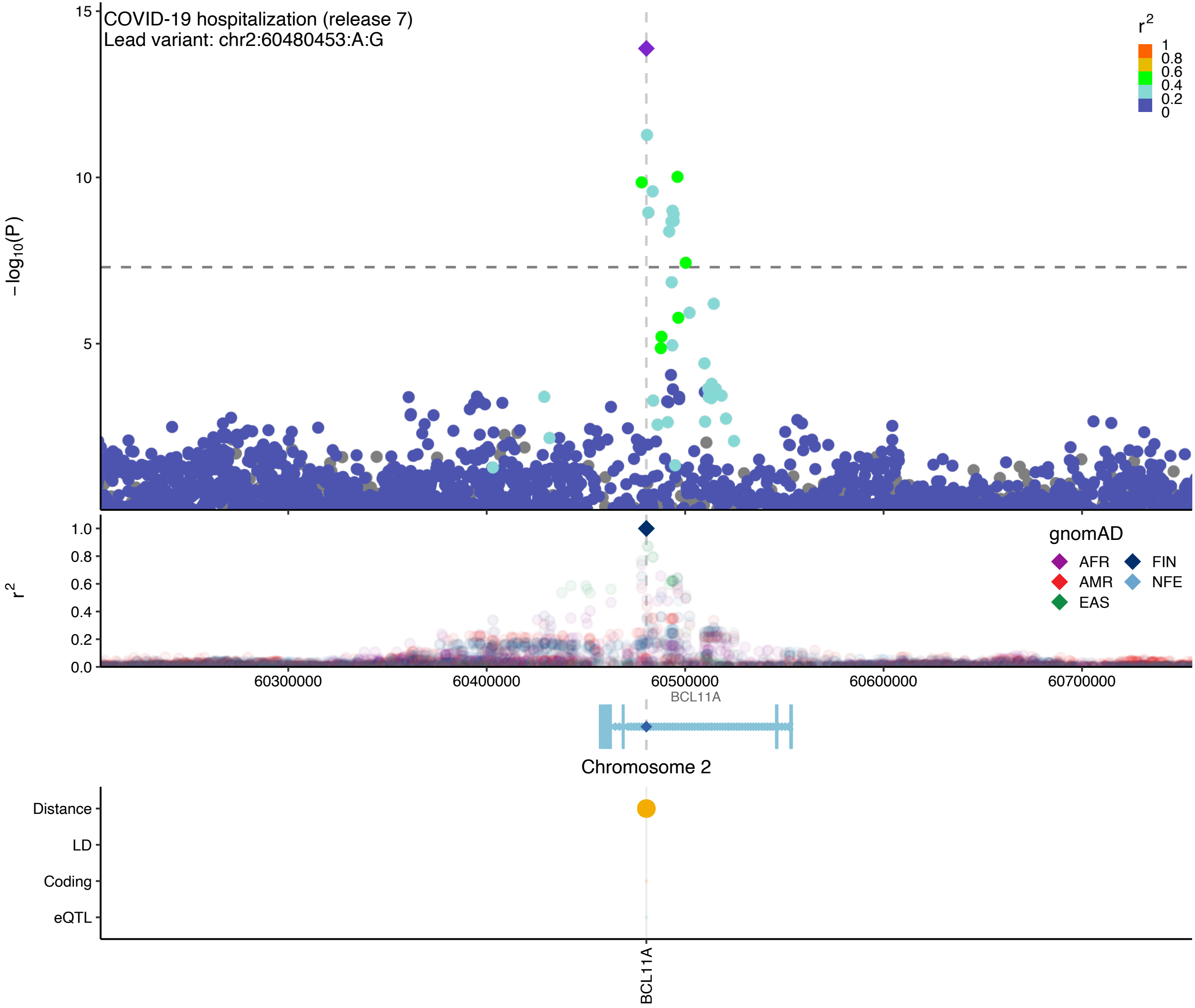
COVID-19 hospitalization (release 7)
Lead variant: chr2:60480453:A:G
−log10(P)
15
10
5
r2
1
0.8
0.6
0.4
0.2
0
1.0
0.8
0.6
0.4
0.2
0.0
gnomAD
AFR
AMR
EAS
FIN
NFE
60300000
60400000
60500000
60600000
60700000
BCL11A
Chromosome 2
Distance
LD
Coding
eQTL
BCL11A

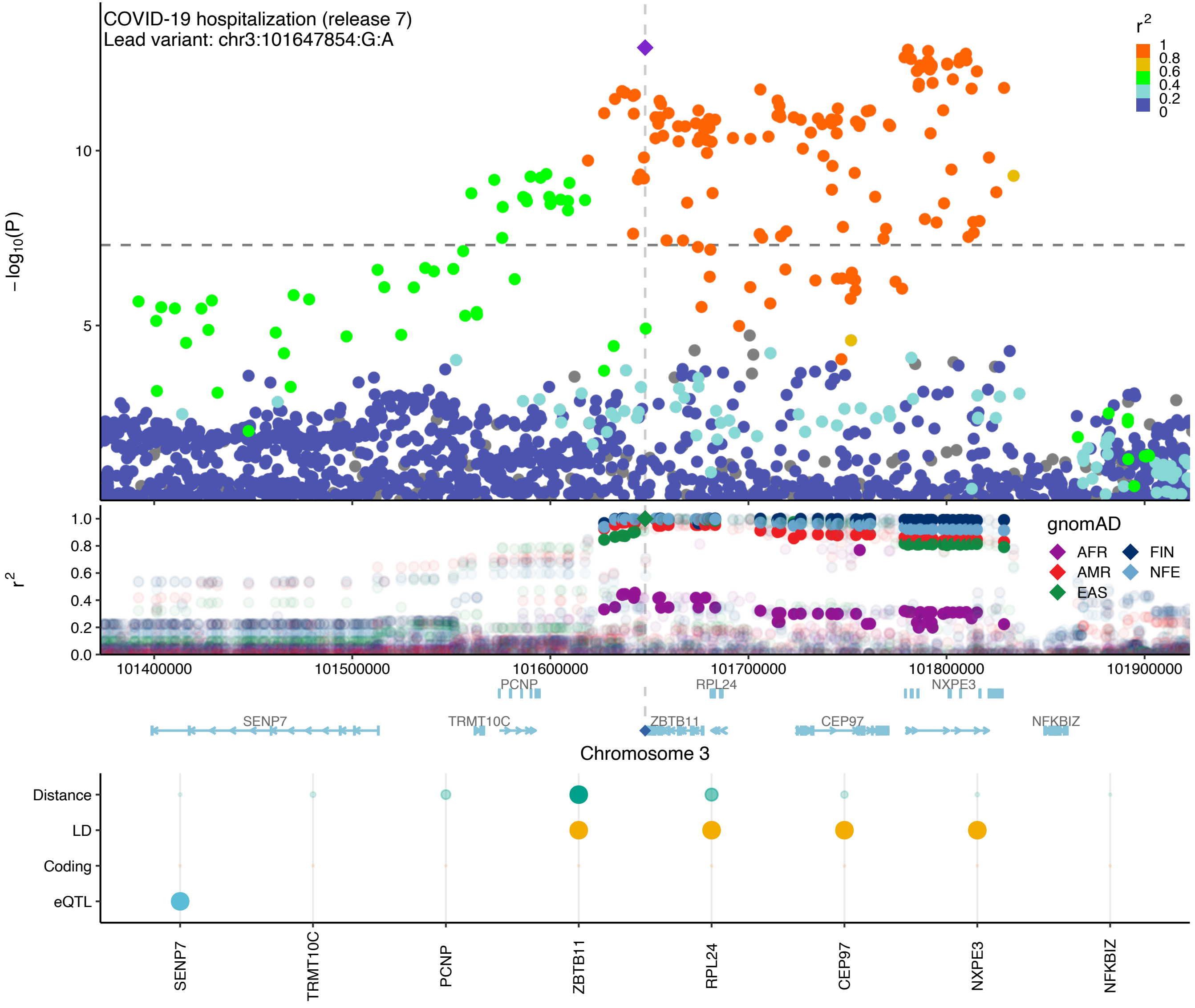
COVID-19 hospitalization (release 7)
Lead variant: chr3:101647854:G:A
$-\log_{10}(P)$
$r^2$
1
0.8
0.6
0.4
0.2
0
gnomAD
AFR
AMR
EAS
FIN
NFE
101400000
101500000
101600000
101700000
101800000
101900000
PCNP
RPL24
NXPE3
SENP7
TRMT10C
ZBTB11
CEP97
NFKBIZ
Chromosome 3
Distance
LD
Coding
eQTL
SENP7
TRMT10C
PCNP
ZBTB11
RPL24
CEP97
NXPE3
NFKBIZ

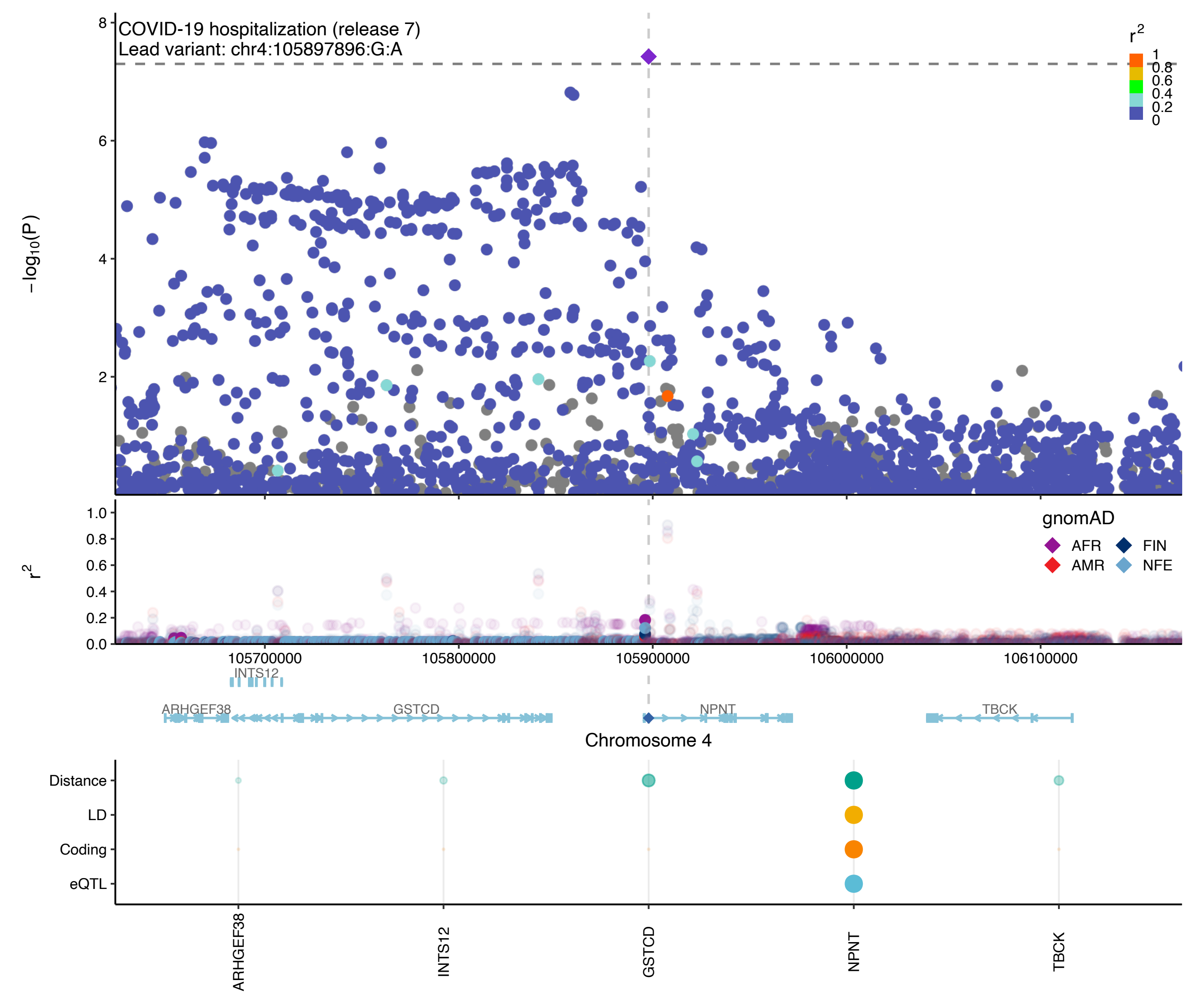

COVID-19 hospitalization (release 7)
Lead variant: chr4:105897896:G:A
−log10(P)
r2
gnomAD
AFR
FIN
AMR
NFE
105700000
105800000
105900000
106000000
106100000
INTS12
ARHGEF38
GSTCD
NPNT
TBCK
Chromosome 4
Distance
LD
Coding
eQTL
ARHGEF38
INTS12
GSTCD
NPNT
TBCK

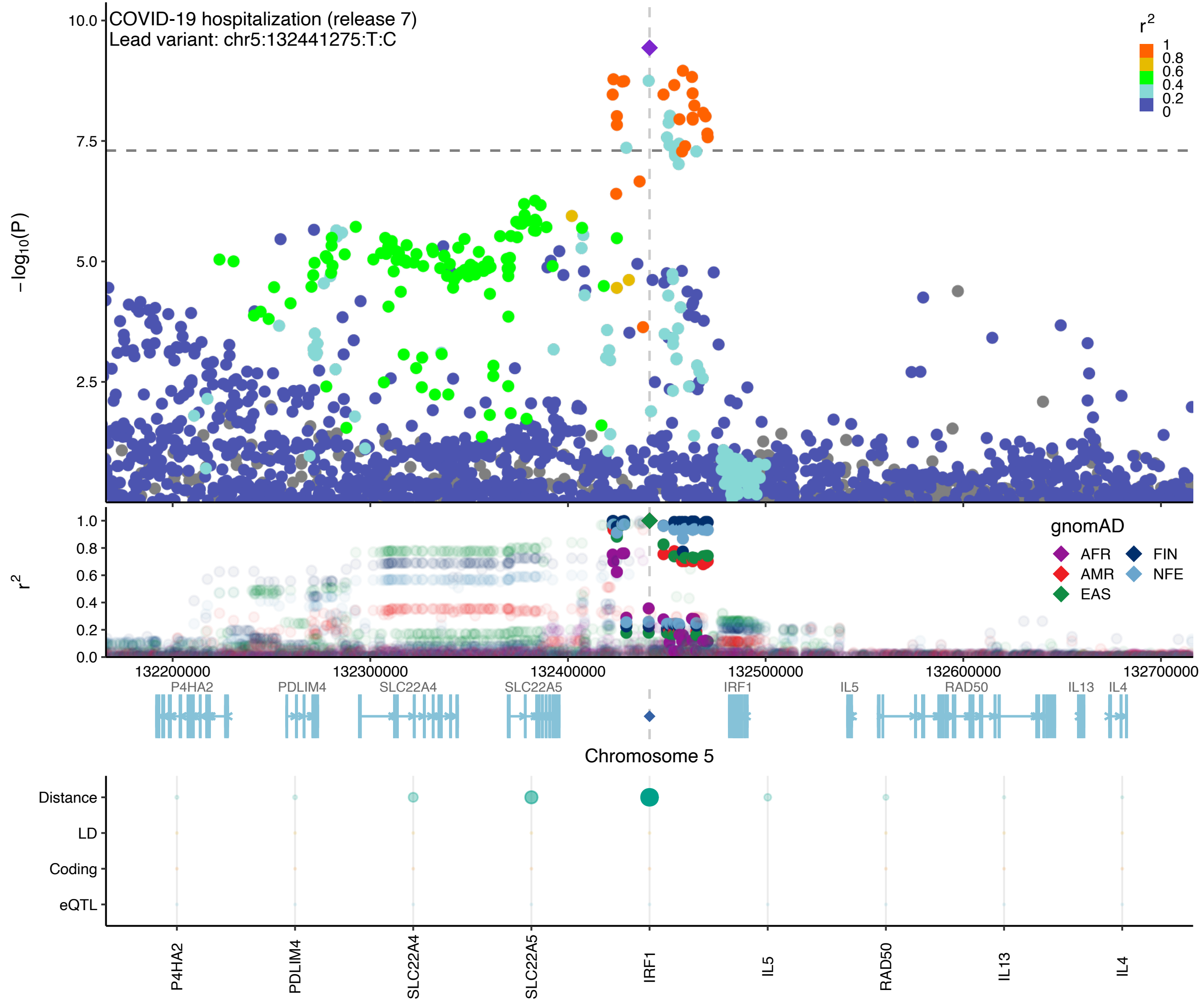

COVID-19 hospitalization (release 7)
Lead variant: chr5:132441275:T:C
−log10(P)
10.0
7.5
5.0
2.5
r2
1
0.8
0.6
0.4
0.2
0
1.0
0.8
0.6
0.4
0.2
0.0
gnomAD
AFR
FIN
AMR
NFE
EAS
132200000
132300000
132400000
132500000
132600000
132700000
P4HA2
PDLIM4
SLC22A4
SLC22A5
IRF1
IL5
RAD50
IL13
IL4
Chromosome 5
Distance
LD
Coding
eQTL
P4HA2
PDLIM4
SLC22A4
SLC22A5
IRF1
IL5
RAD50
IL13
IL4

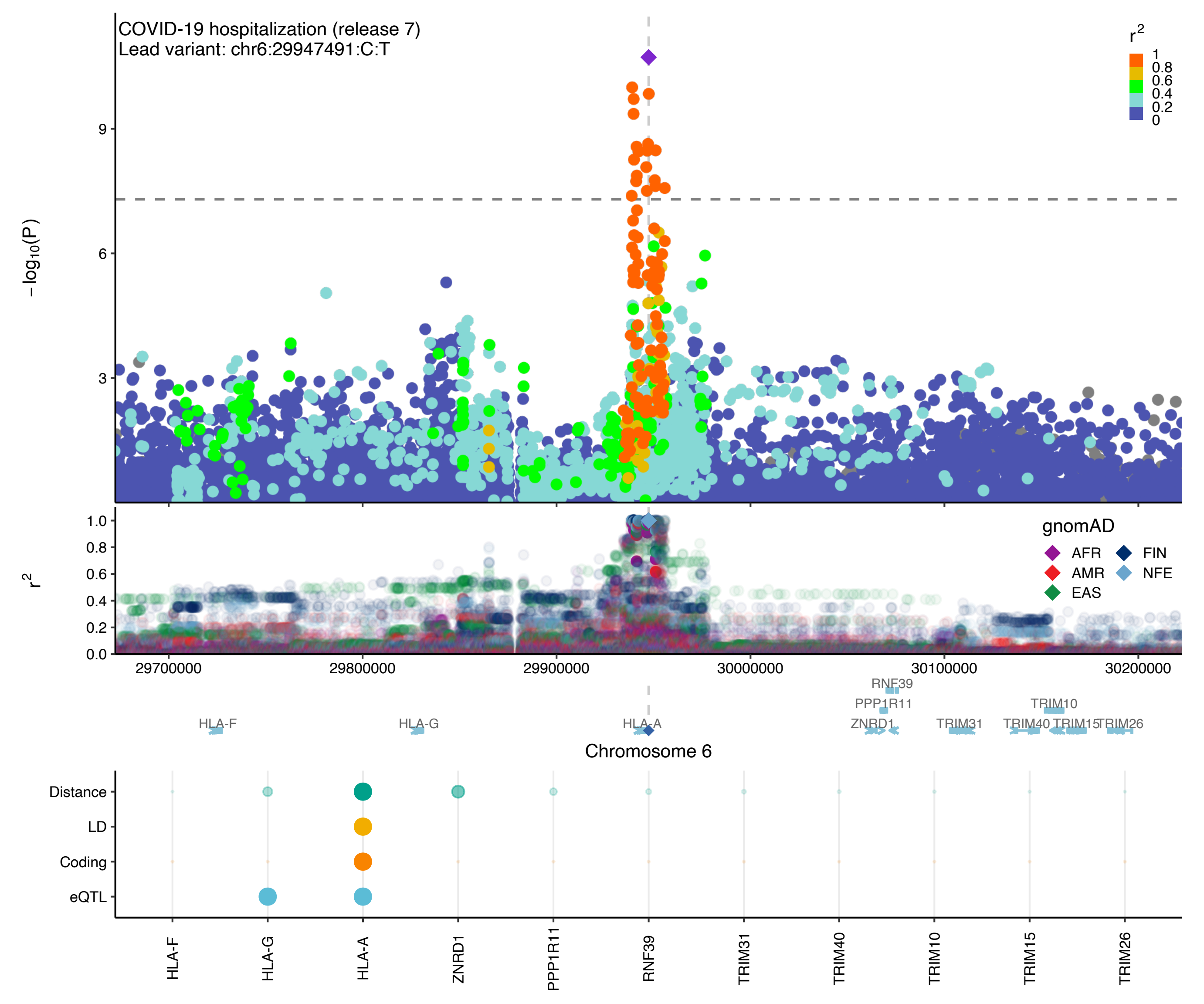

COVID-19 hospitalization (release 7)
Lead variant: chr6:29947491:C:T
r2
1
0.8
0.6
0.4
0.2
0
−log10(P)
9
6
3
r2
1.0
0.8
0.6
0.4
0.2
0.0
gnomAD
AFR
FIN
AMR
NFE
EAS
29700000
29800000
29900000
30000000
30100000
30200000
RNF39
PPP1R11
TRIM10
HLA-F
HLA-G
HLA-A
ZNRD1
TRIM31
TRIM40
TRIM15
TRIM26
Chromosome 6
Distance
LD
Coding
eQTL
HLA-F
HLA-G
HLA-A
ZNRD1
PPP1R11
RNF39
TRIM31
TRIM40
TRIM10
TRIM15
TRIM26

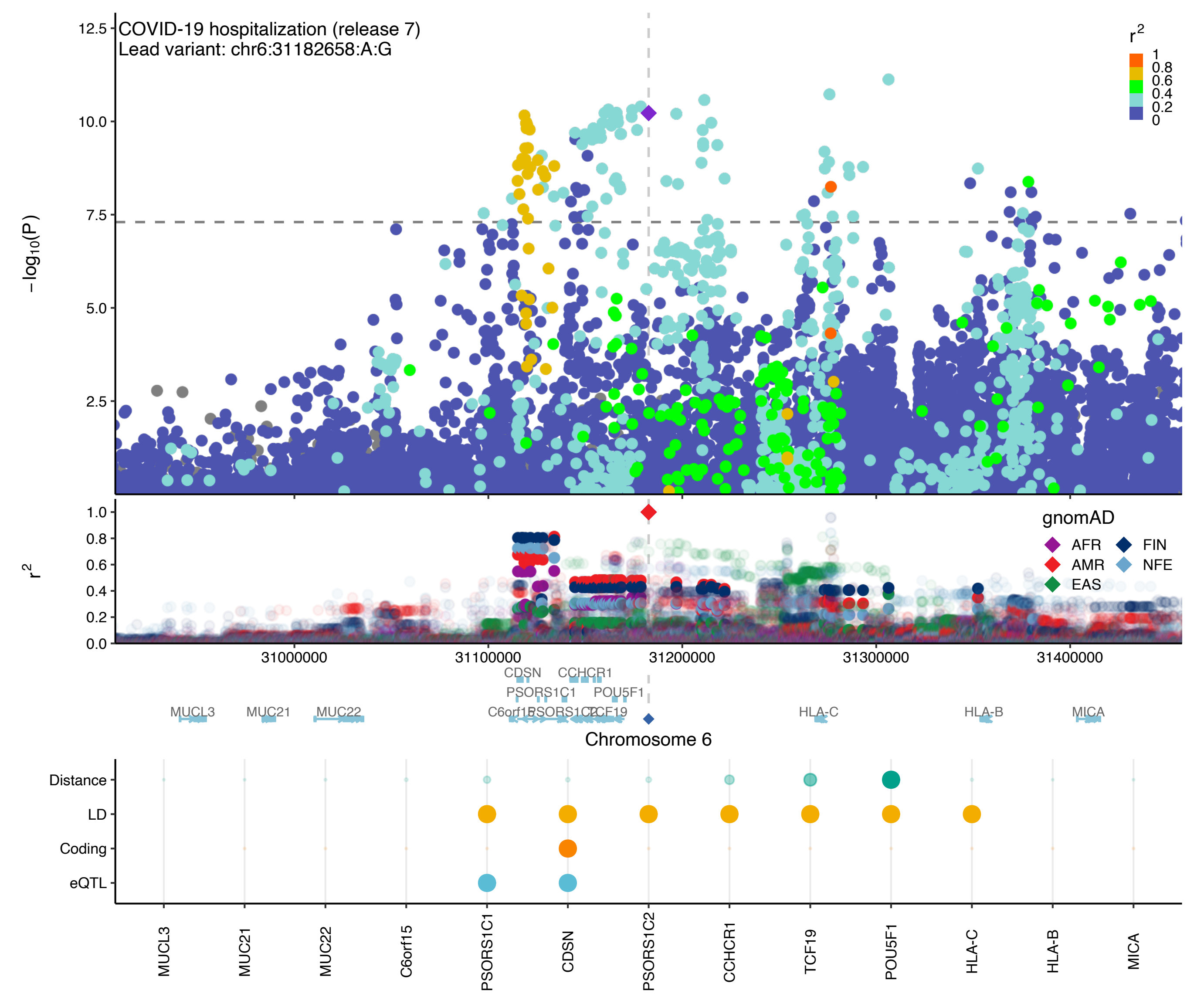

COVID-19 hospitalization (release 7)
Lead variant: chr6:31182658:A:G
$-\log_{10}(P)$
$r^2$
1
0.8
0.6
0.4
0.2
0
gnomAD
AFR
AMR
EAS
FIN
NFE
31000000
31100000
31200000
31300000
31400000
CDSN
CCHCR1
PSORS1C1
POU5F1
MUCL3
MUC21
MUC22
C6orf15
PSORS1C2
TCF19
HLA-C
HLA-B
MICA
Chromosome 6
Distance
LD
Coding
eQTL
MUCL3
MUC21
MUC22
C6orf15
PSORS1C1
CDSN
PSORS1C2
CCHCR1
TCF19
POU5F1
HLA-C
HLA-B
MICA

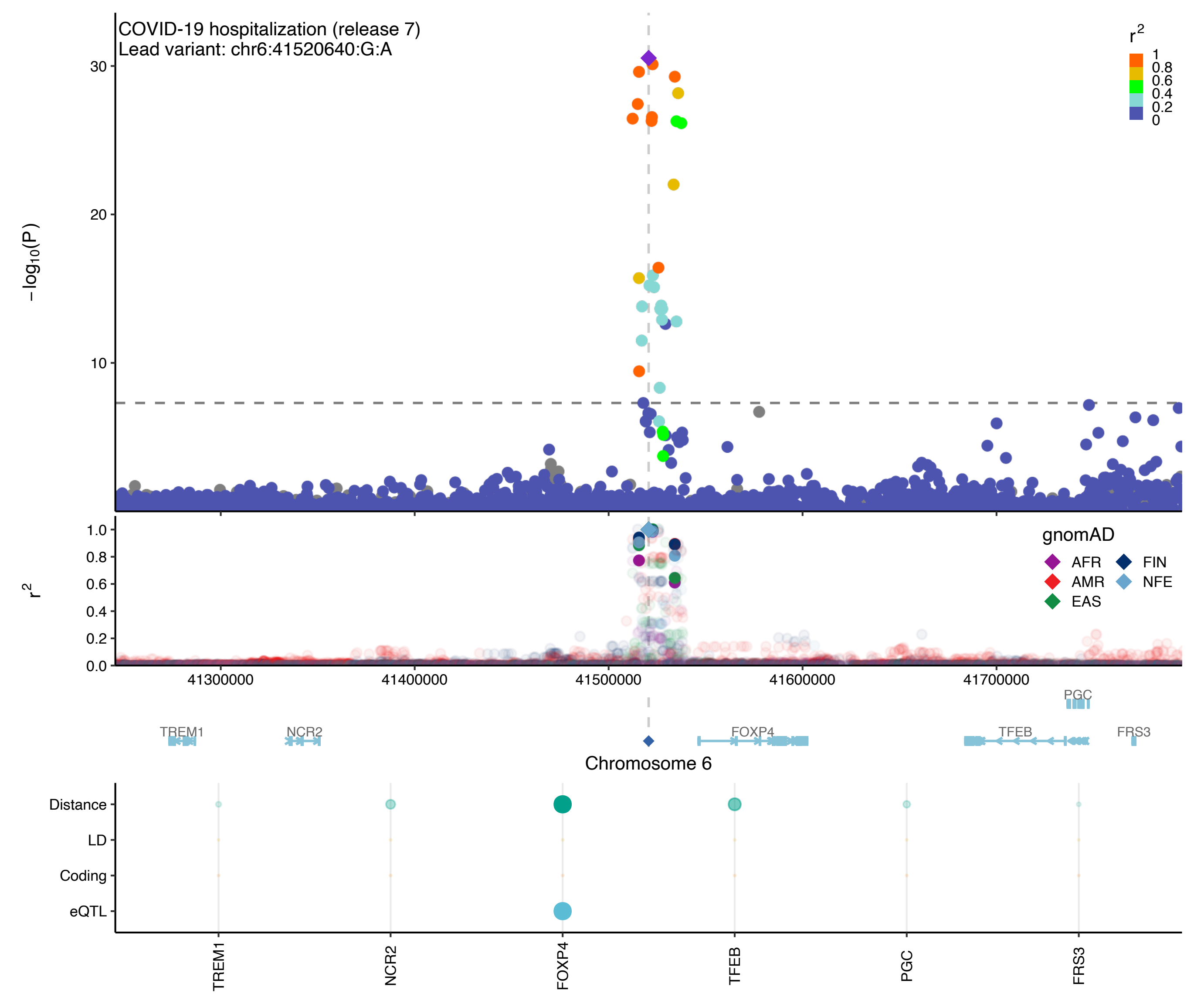
COVID-19 hospitalization (release 7)
Lead variant: chr6:41520640:G:A
r2
1
0.8
0.6
0.4
0.2
0
30
20
10
−log10(P)
1.0
0.8
0.6
0.4
0.2
0.0
r2
gnomAD
AFR
FIN
AMR
NFE
EAS
41300000
41400000
41500000
41600000
41700000
PGC
TREM1
NCR2
FOXP4
TFEB
FRS3
Chromosome 6
Distance
LD
Coding
eQTL
TREM1
NCR2
FOXP4
TFEB
PGC
FRS3

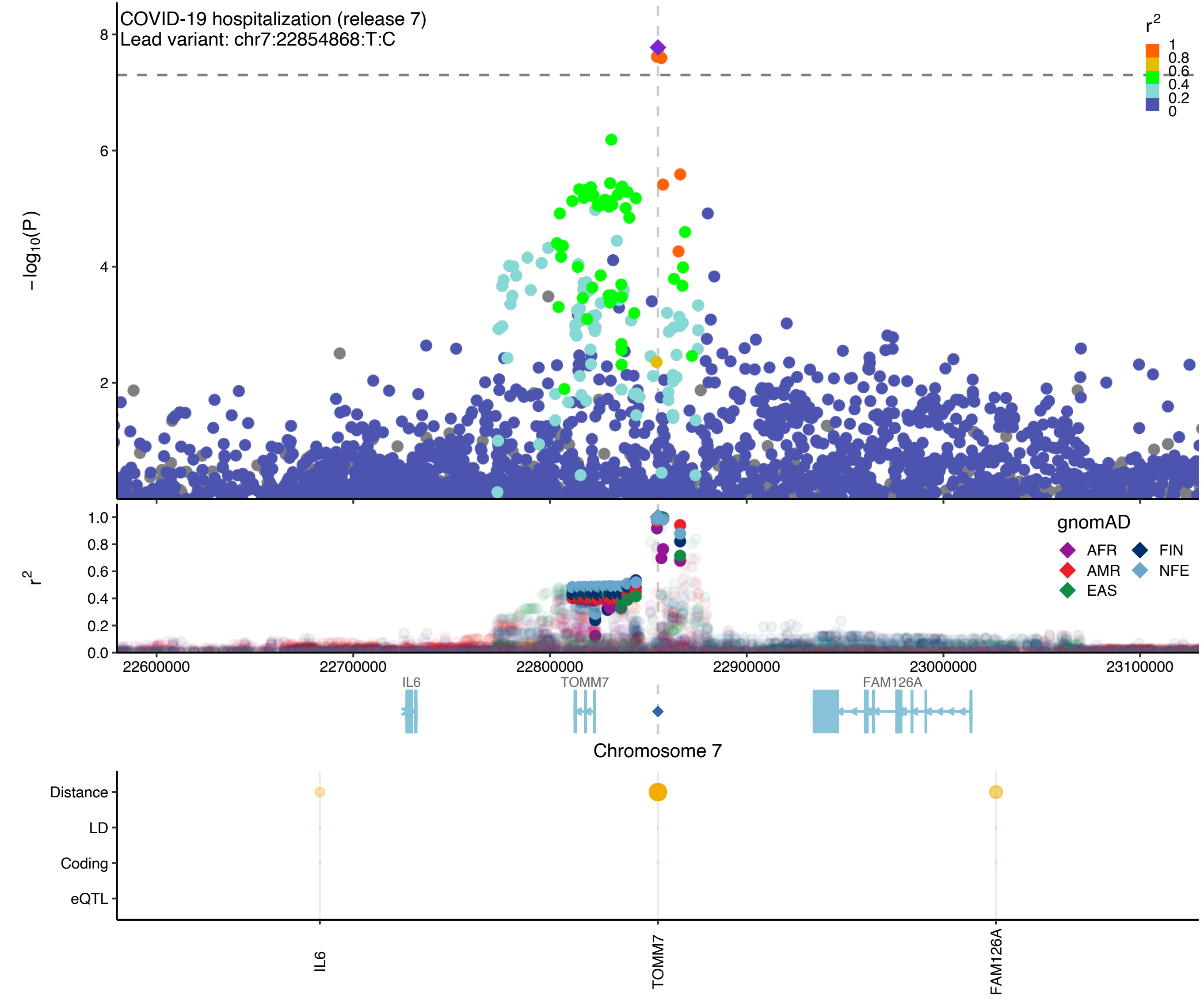

COVID-19 hospitalization (release 7)
Lead variant: chr7:22854868:T:C
−log10(P)
8
6
4
2
r2
1
0.8
0.6
0.4
0.2
0
r2
1.0
0.8
0.6
0.4
0.2
0.0
gnomAD
AFR
FIN
AMR
NFE
EAS
22600000
22700000
22800000
22900000
23000000
23100000
IL6
TOMM7
FAM126A
Chromosome 7
Distance
LD
Coding
eQTL
IL6
TOMM7
FAM126A

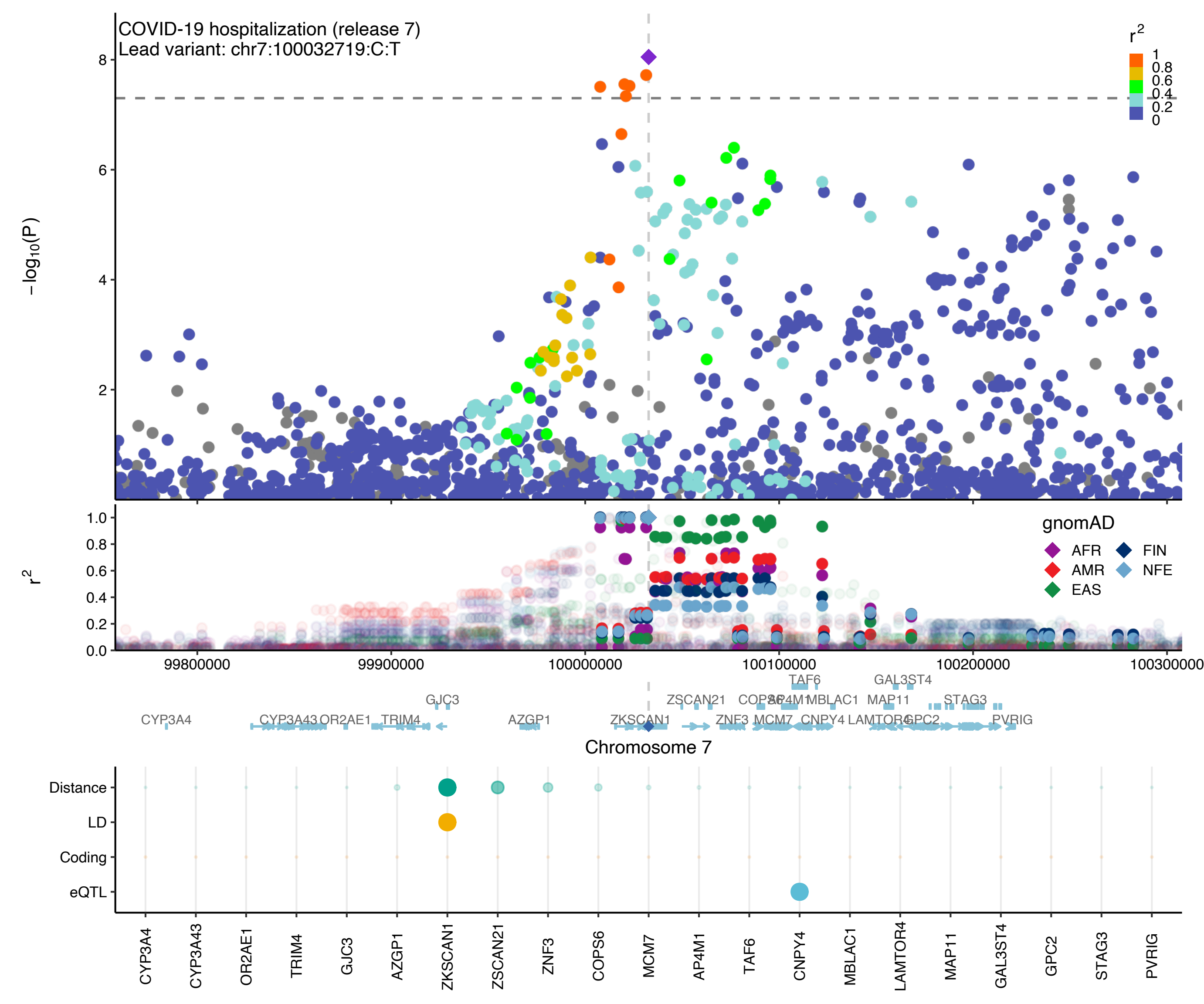

COVID-19 hospitalization (release 7)
Lead variant: chr7:100032719:C:T
r2
1
0.8
0.6
0.4
0.2
0
−log10(P)
8
6
4
2
r2
1.0
0.8
0.6
0.4
0.2
0.0
gnomAD
AFR
AMR
EAS
FIN
NFE
99800000
99900000
100000000
100100000
100200000
100300000
TAF6
GAL3ST4
GJC3
ZSCAN21
COPS6
AP4M1
MBLAC1
MAP11
STAG3
CYP3A4
CYP3A43
OR2AE1
TRIM4
AZGP1
ZKSCAN1
ZNF3
MCM7
CNPY4
LAMTOR4
GPC2
PVRIG
Chromosome 7
Distance
LD
Coding
eQTL
CYP3A4
CYP3A43
OR2AE1
TRIM4
GJC3
AZGP1
ZKSCAN1
ZSCAN21
ZNF3
COPS6
MCM7
AP4M1
TAF6
CNPY4
MBLAC1
LAMTOR4
MAP11
GAL3ST4
GPC2
STAG3
PVRIG

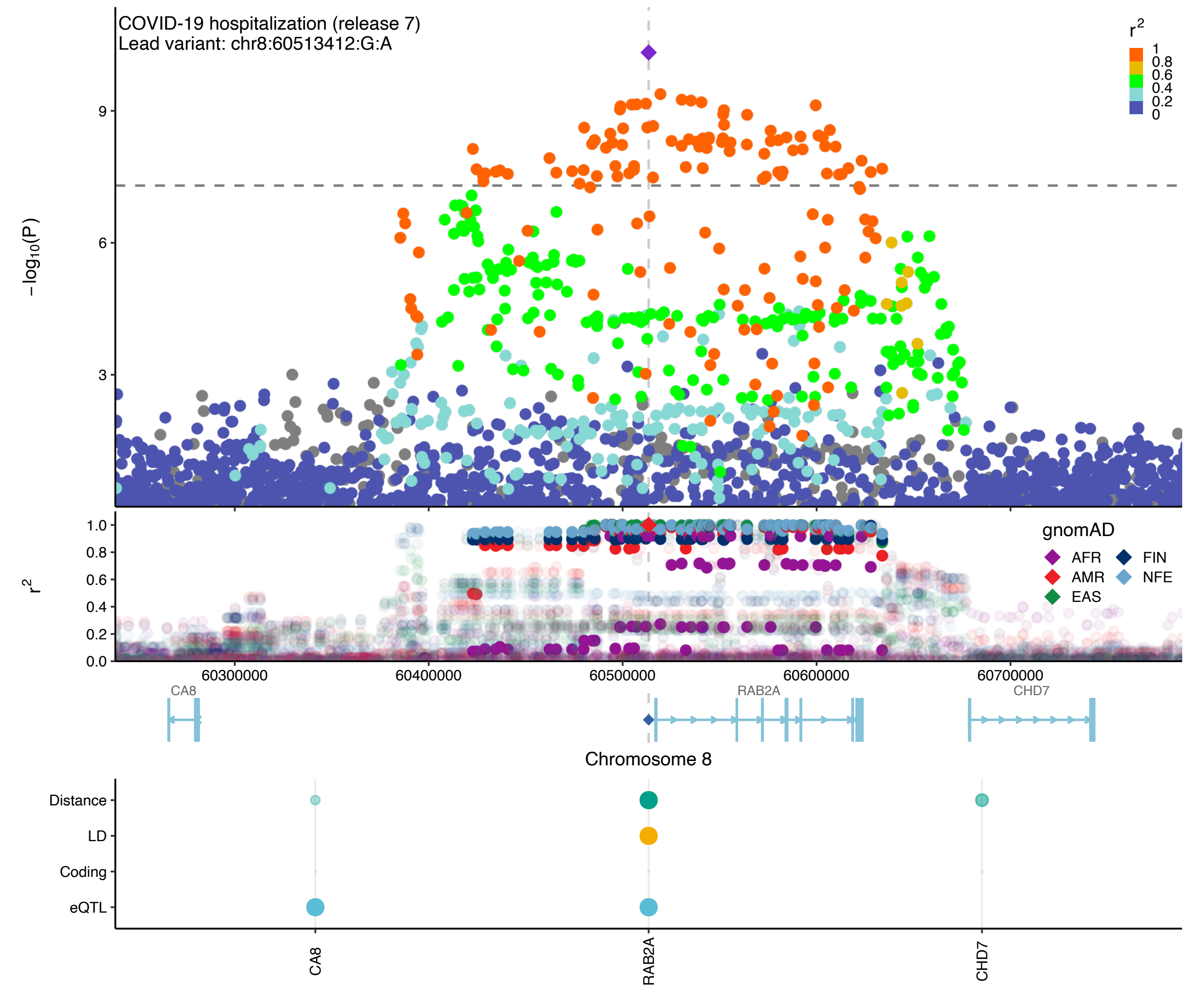
COVID-19 hospitalization (release 7)
Lead variant: chr8:60513412:G:A
r2
1
0.8
0.6
0.4
0.2
0
−log10(P)
9
6
3
r2
1.0
0.8
0.6
0.4
0.2
0.0
gnomAD
AFR
FIN
AMR
NFE
EAS
60300000
60400000
60500000
60600000
60700000
CA8
RAB2A
CHD7
Chromosome 8
Distance
LD
Coding
eQTL
CA8
RAB2A
CHD7

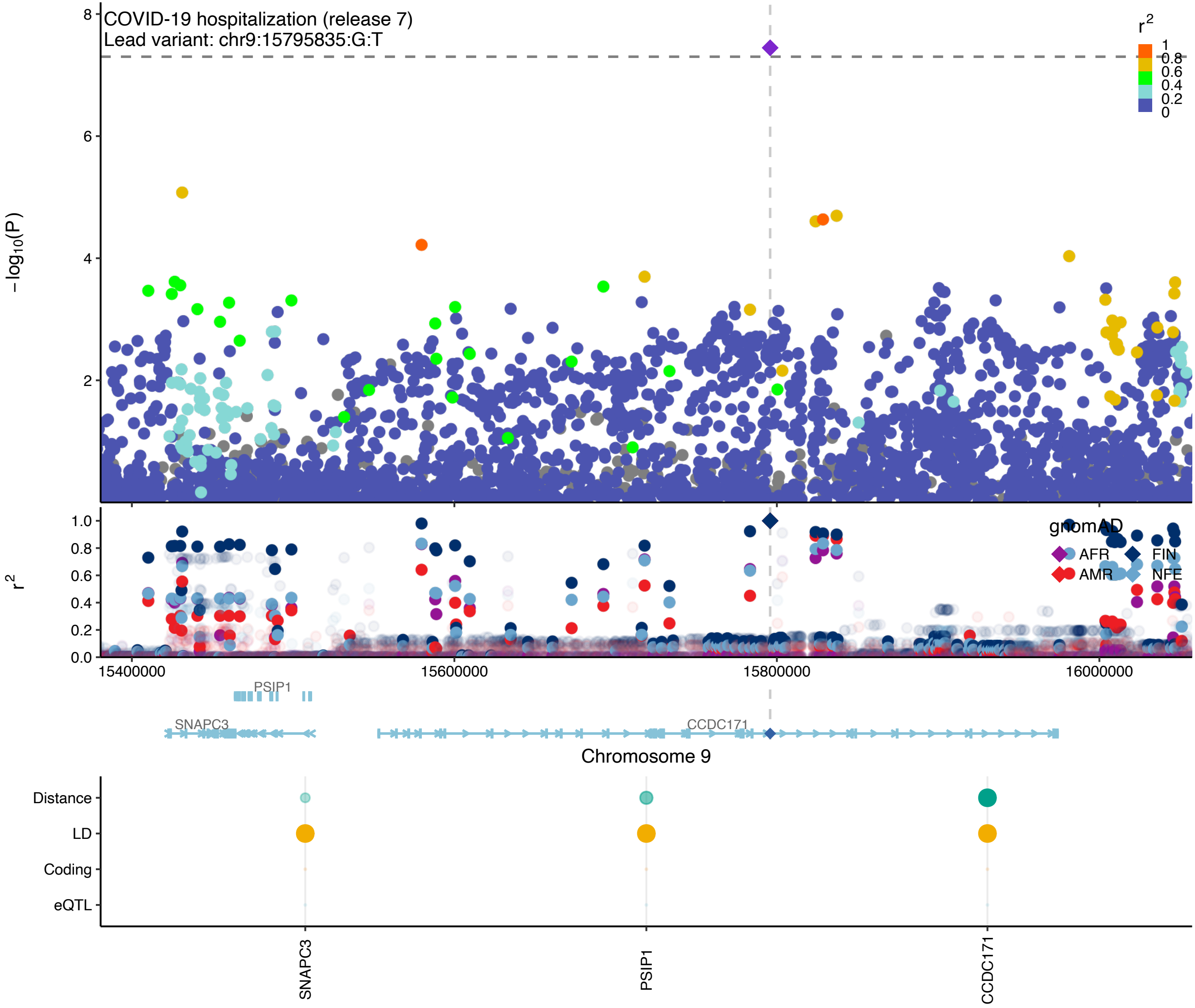

COVID-19 hospitalization (release 7)
Lead variant: chr9:15795835:G:T
−log10(P)
8
6
4
2
r2
1
0.8
0.6
0.4
0.2
0
r2
1.0
0.8
0.6
0.4
0.2
0.0
gnomAD
AFR
FIN
AMR
NFE
15400000
15600000
15800000
16000000
PSIP1
SNAPC3
CCDC171
Chromosome 9
Distance
LD
Coding
eQTL
SNAPC3
PSIP1
CCDC171

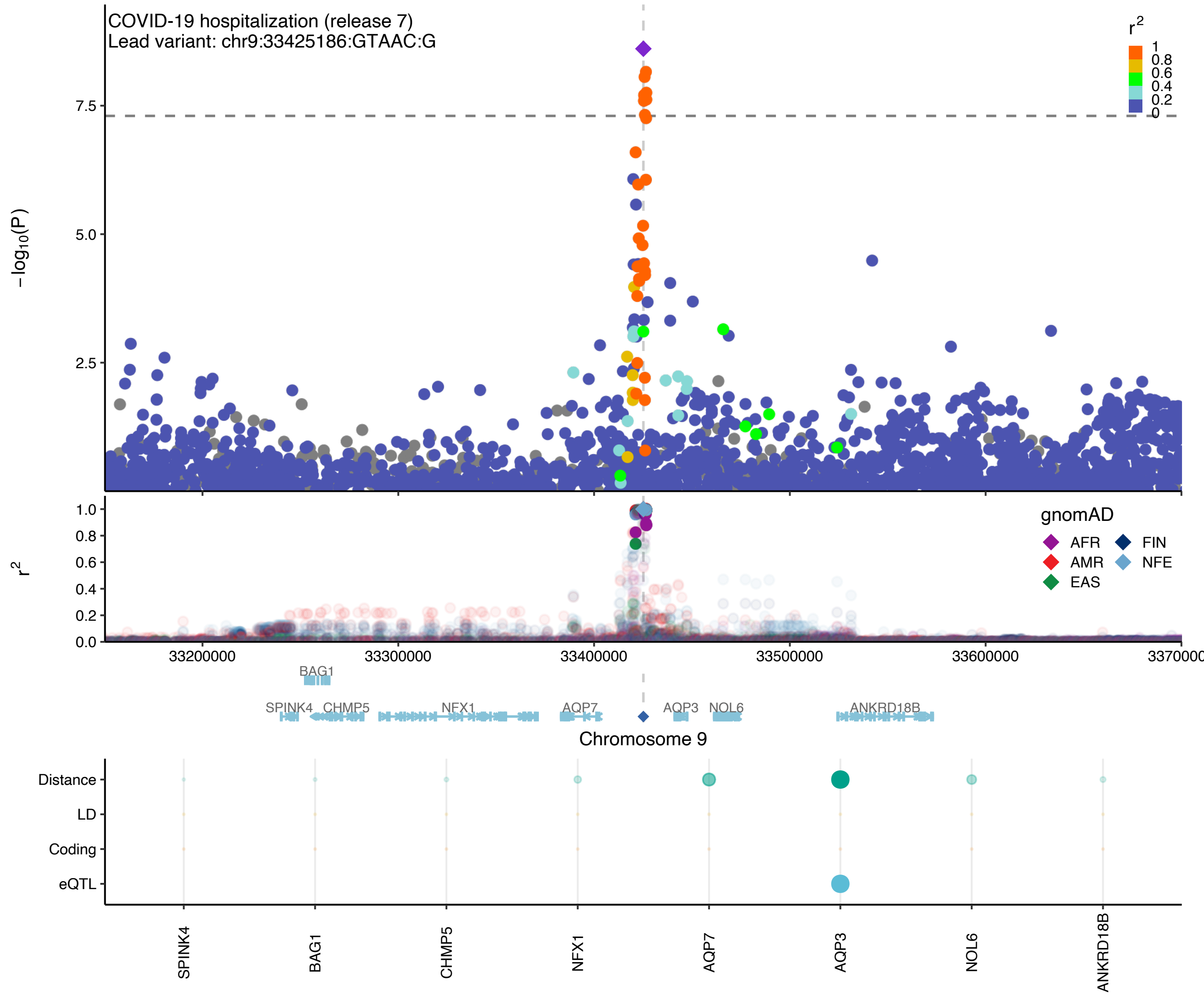

COVID-19 hospitalization (release 7)
Lead variant: chr9:33425186:GTAAC:G
r2
1
0.8
0.6
0.4
0.2
0
−log10(P)
7.5
5.0
2.5
r2
1.0
0.8
0.6
0.4
0.2
0.0
gnomAD
AFR
FIN
AMR
NFE
EAS
33200000
33300000
33400000
33500000
33600000
BAG1
SPINK4
CHMP5
NFX1
AQP7
AQP3
NOL6
ANKRD18B
Chromosome 9
Distance
LD
Coding
eQTL
SPINK4
BAG1
CHMP5
NFX1
AQP7
AQP3
NOL6
ANKRD18B

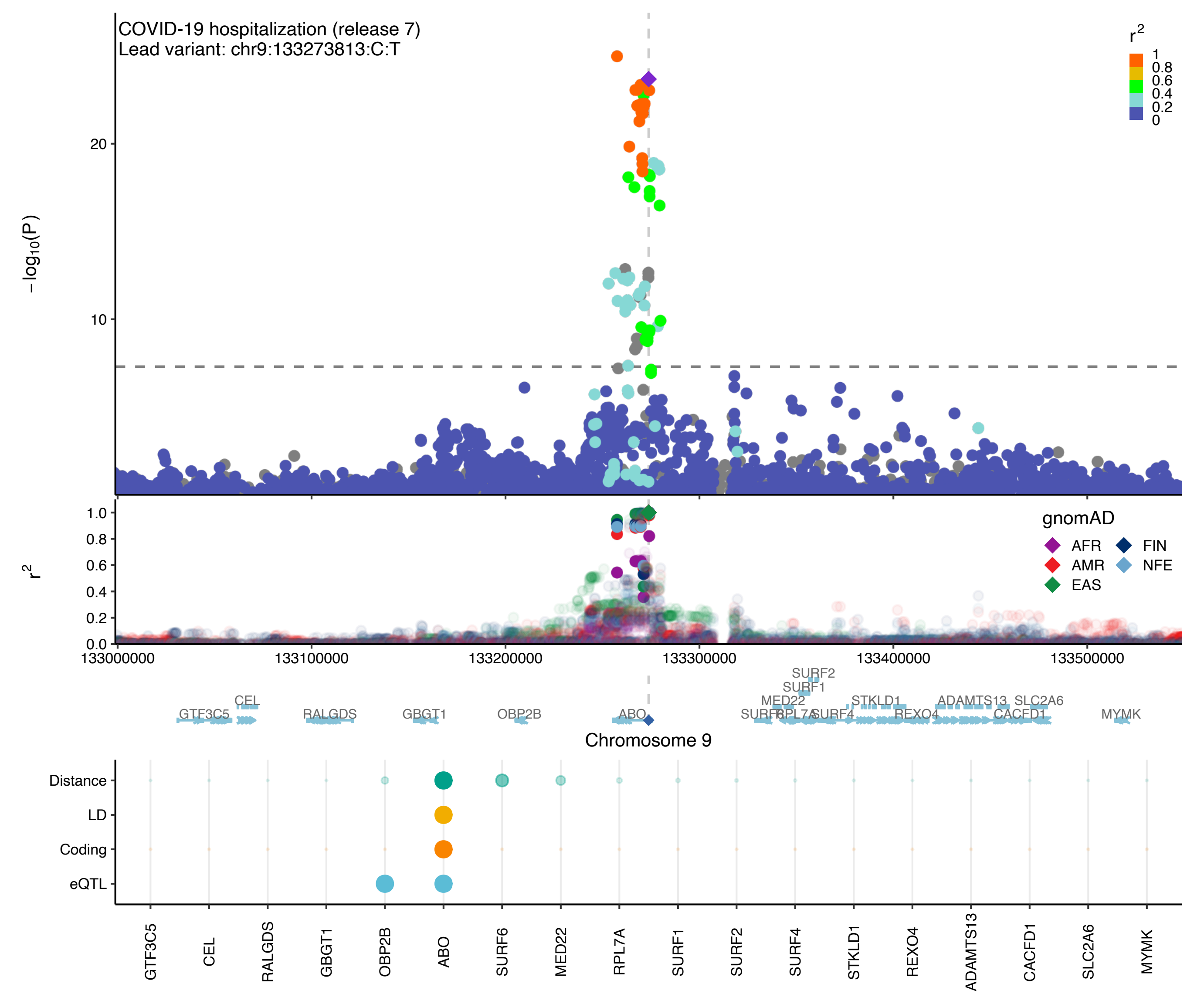

COVID-19 hospitalization (release 7)
Lead variant: chr9:133273813:C:T
−log10(P)
20
10
r2
1
0.8
0.6
0.4
0.2
0
1.0
0.8
0.6
0.4
0.2
0.0
gnomAD
AFR
FIN
AMR
NFE
EAS
133000000
133100000
133200000
133300000
133400000
133500000
SURF2
SURF1
MED22
STKLD1
ADAMTS13
SLC2A6
CEL
GTF3C5
RALGDS
GBGT1
OBP2B
ABO
SURF6
RPL7A
SURF4
REXO4
CACFD1
MYMK
Chromosome 9
Distance
LD
Coding
eQTL
GTF3C5
CEL
RALGDS
GBGT1
OBP2B
ABO
SURF6
MED22
RPL7A
SURF1
SURF2
SURF4
STKLD1
REXO4
ADAMTS13
CACFD1
SLC2A6
MYMK

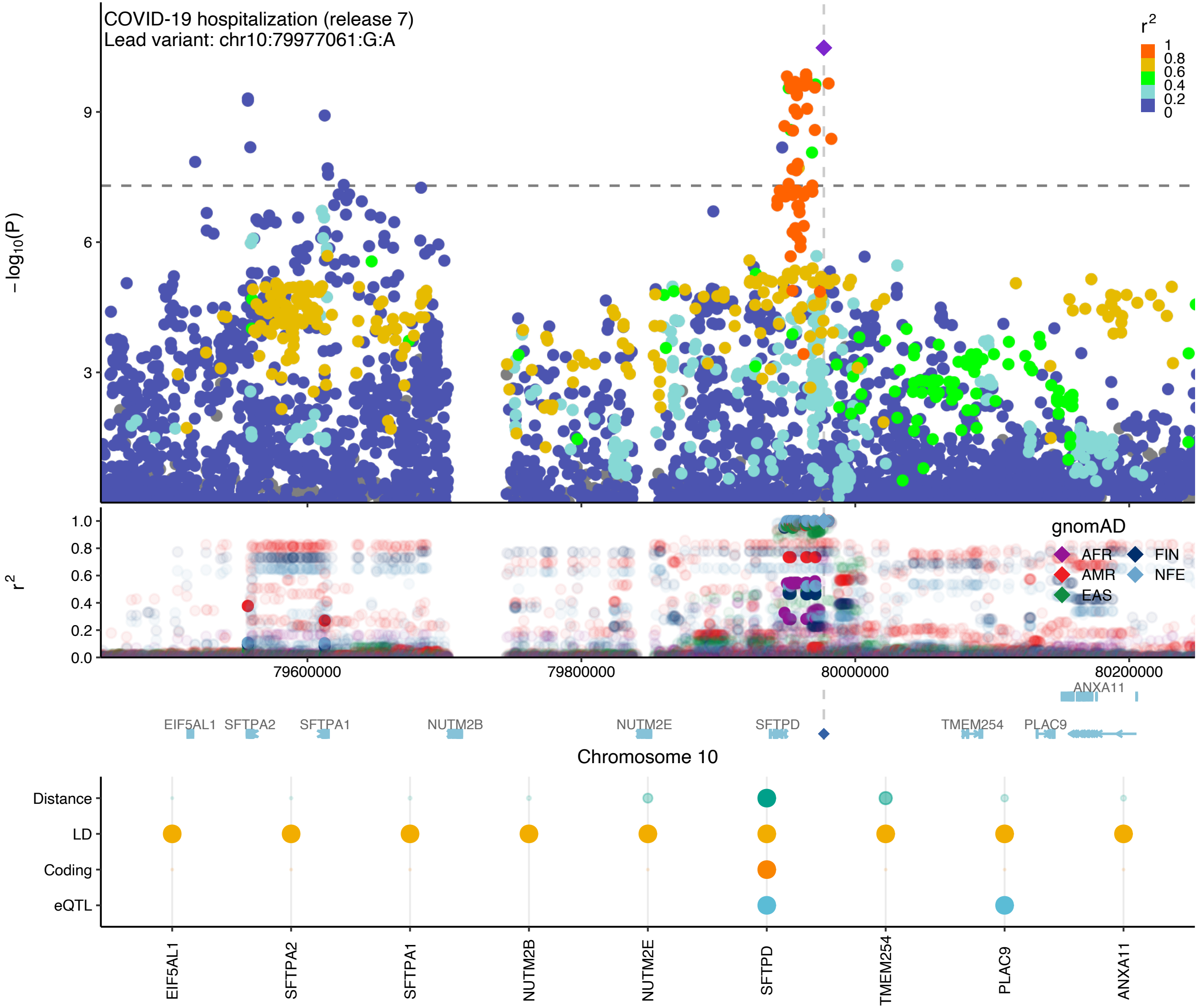

COVID-19 hospitalization (release 7)
Lead variant: chr10:79977061:G:A
r2
1
0.8
0.6
0.4
0.2
0
−log10(P)
9
6
3
r2
1.0
0.8
0.6
0.4
0.2
0.0
gnomAD
AFR
FIN
AMR
NFE
EAS
79600000
79800000
80000000
80200000
ANXA11
EIF5AL1
SFTPA2
SFTPA1
NUTM2B
NUTM2E
SFTPD
TMEM254
PLAC9
Chromosome 10
Distance
LD
Coding
eQTL
EIF5AL1
SFTPA2
SFTPA1
NUTM2B
NUTM2E
SFTPD
TMEM254
PLAC9
ANXA11

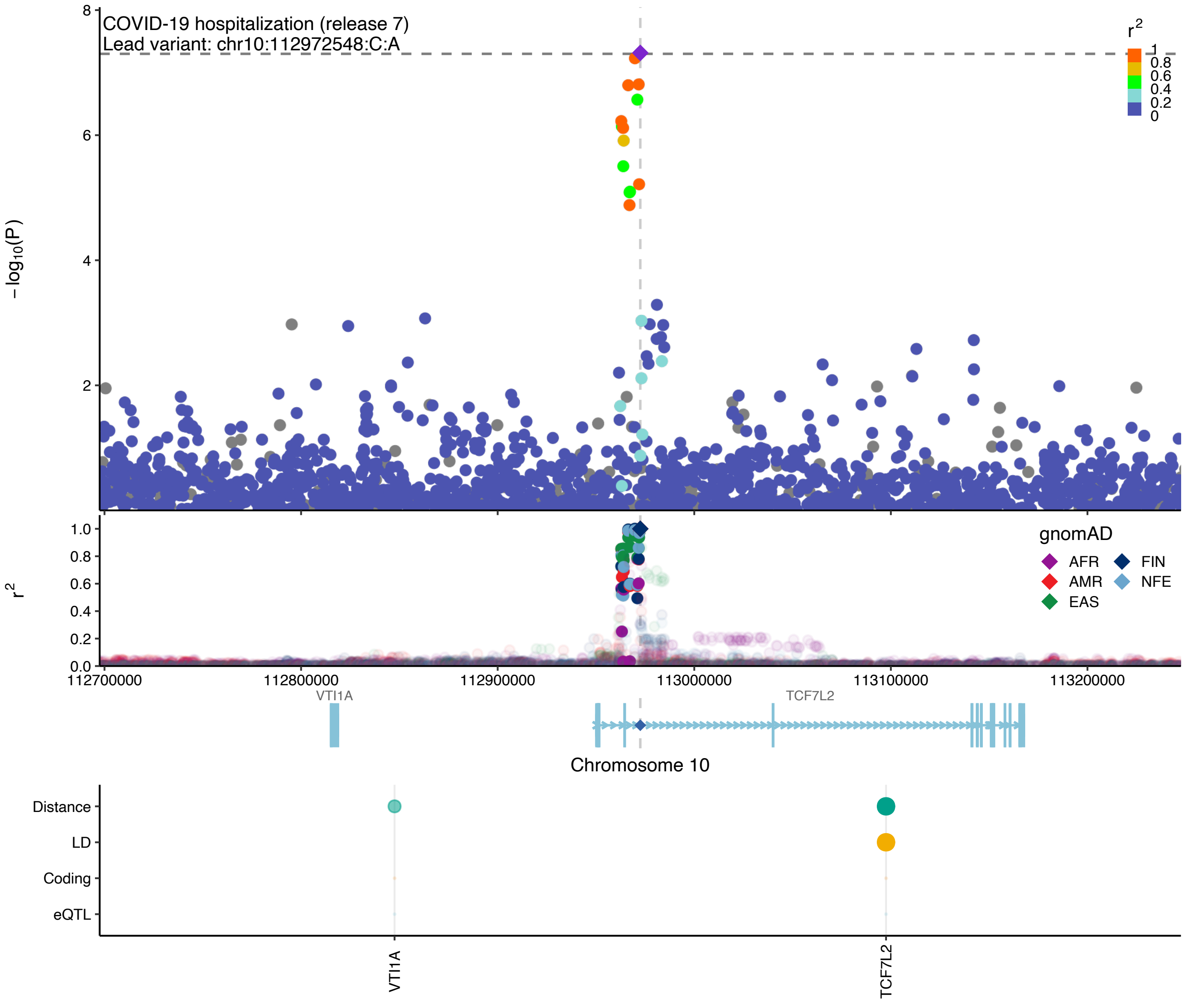

COVID-19 hospitalization (release 7)
Lead variant: chr10:112972548:C:A
−log10(P)
r2
1
0.8
0.6
0.4
0.2
0
gnomAD
AFR
AMR
EAS
FIN
NFE
112700000
112800000
112900000
113000000
113100000
113200000
VTI1A
TCF7L2
Chromosome 10
Distance
LD
Coding
eQTL
VTI1A
TCF7L2

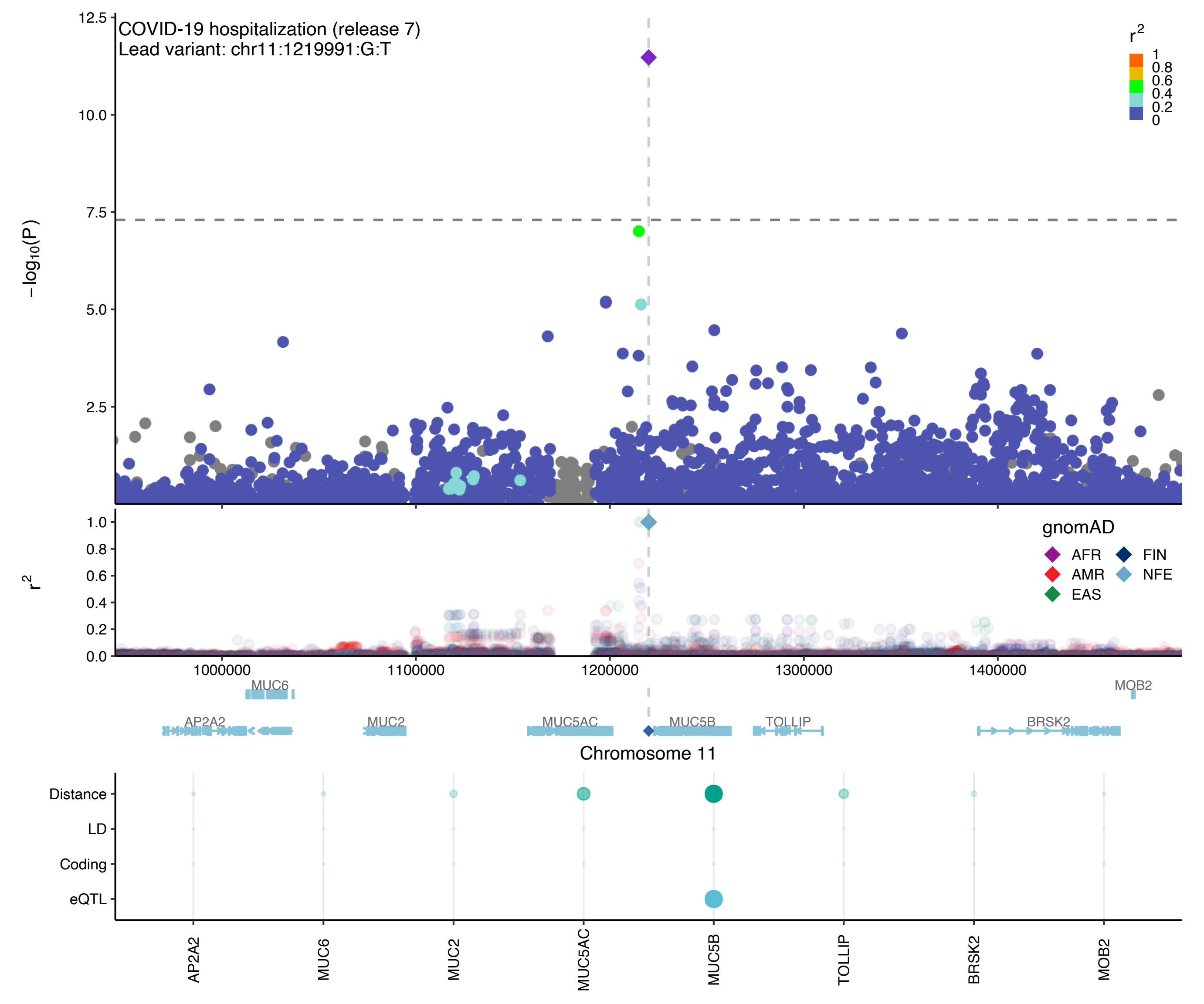
COVID-19 hospitalization (release 7)
Lead variant: chr11:1219991:G:T
−log10(P)
12.5
10.0
7.5
5.0
2.5
r2
1
0.8
0.6
0.4
0.2
0
1.0
0.8
0.6
0.4
0.2
0.0
gnomAD
AFR
FIN
AMR
NFE
EAS
1000000
1100000
1200000
1300000
1400000
MUC6
MOB2
AP2A2
MUC2
MUC5AC
MUC5B
TOLLIP
BRSK2
Chromosome 11
Distance
LD
Coding
eQTL
AP2A2
MUC6
MUC2
MUC5AC
MUC5B
TOLLIP
BRSK2
MOB2

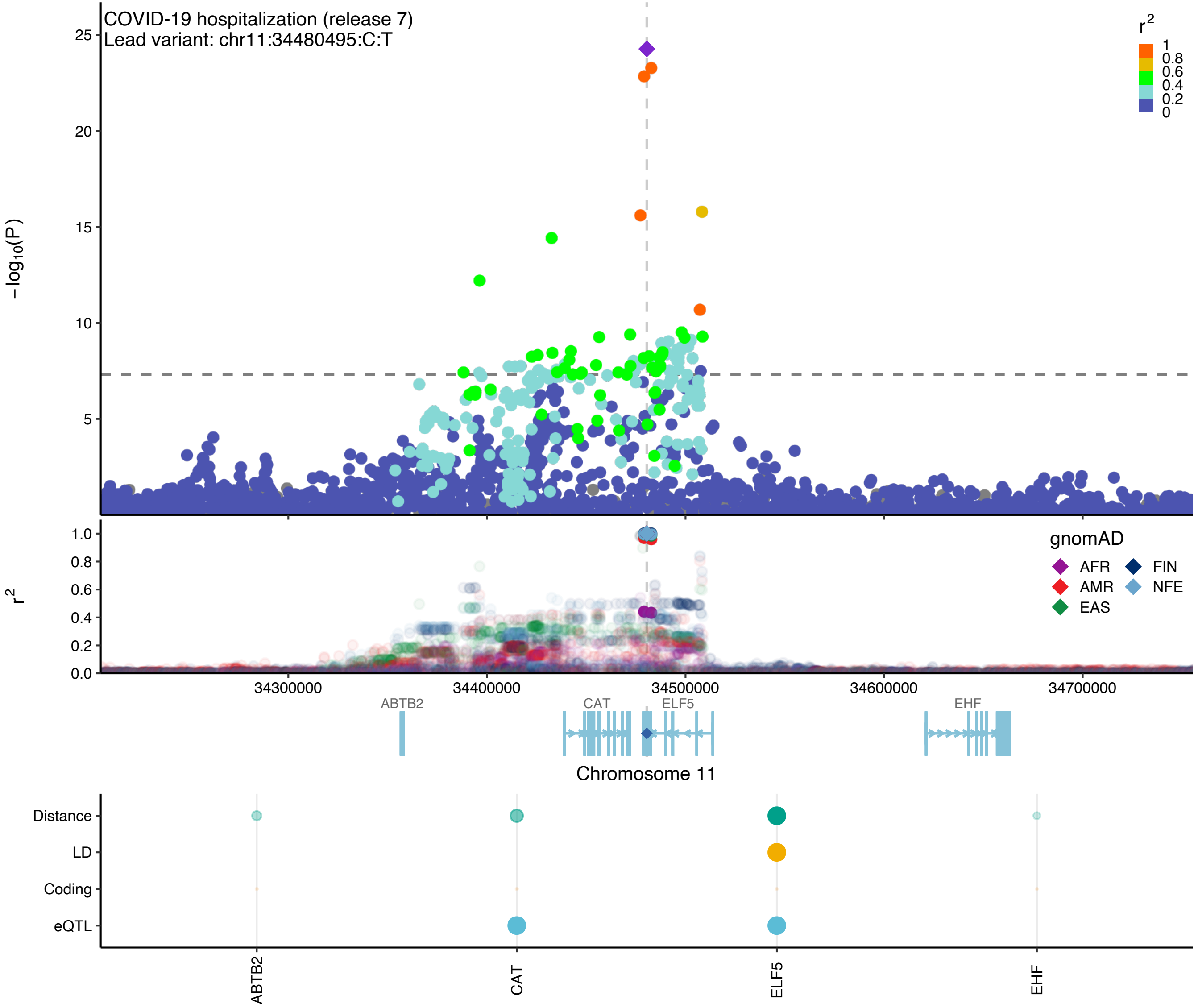

COVID-19 hospitalization (release 7)
Lead variant: chr11:34480495:C:T
−log10(P)
25
20
15
10
5
r2
1
0.8
0.6
0.4
0.2
0
1.0
0.8
0.6
0.4
0.2
0.0
gnomAD
AFR
FIN
AMR
NFE
EAS
34300000
34400000
34500000
34600000
34700000
ABTB2
CAT
ELF5
EHF
Chromosome 11
Distance
LD
Coding
eQTL
ABTB2
CAT
ELF5
EHF

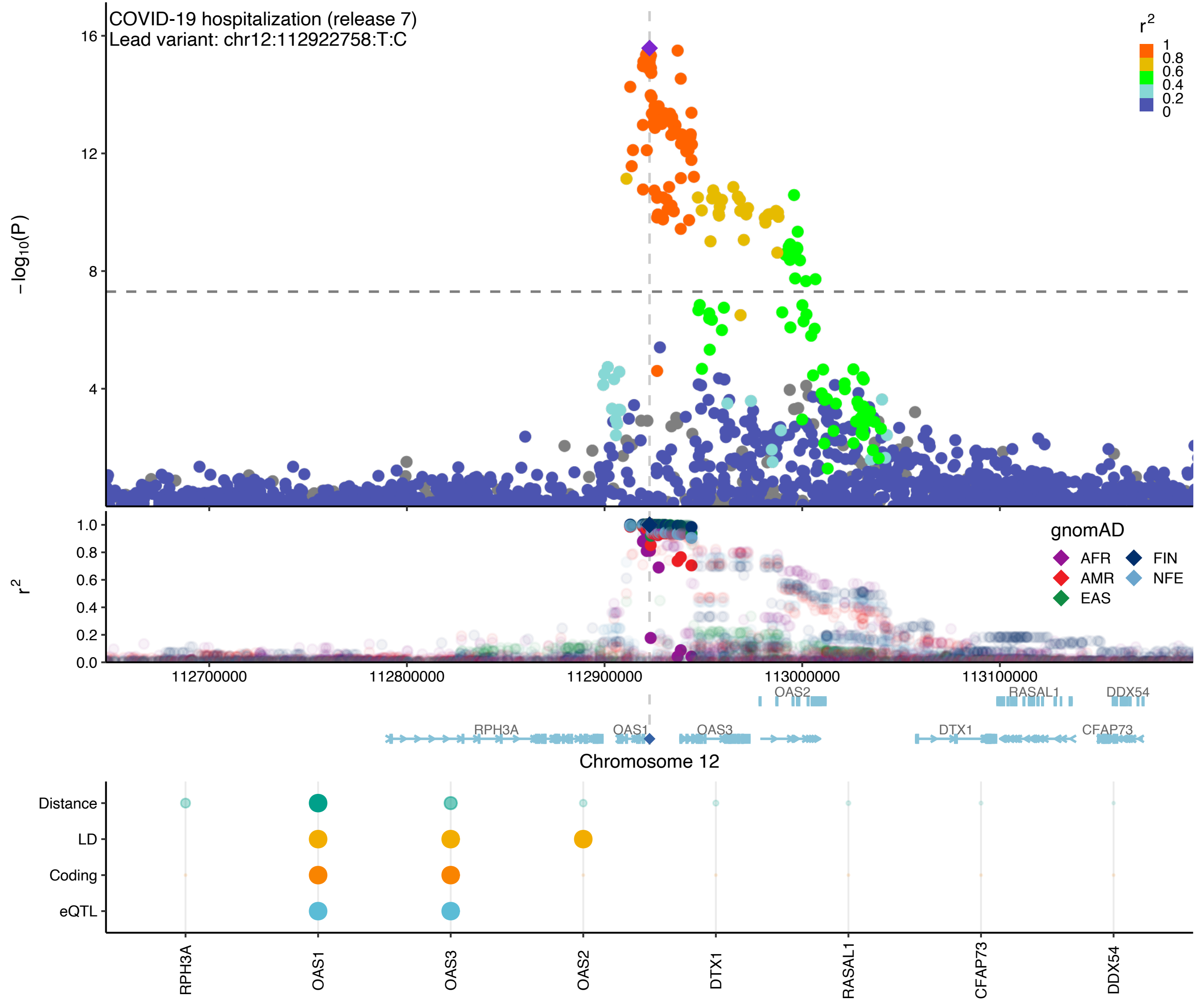

COVID-19 hospitalization (release 7)
Lead variant: chr12:112922758:T:C
$-\log_{10}(P)$
16
12
8
4
$r^2$
1
0.8
0.6
0.4
0.2
0
$r^2$
1.0
0.8
0.6
0.4
0.2
0.0
gnomAD
AFR
AMR
EAS
FIN
NFE
112700000
112800000
112900000
113000000
113100000
OAS2
RASAL1
DDX54
RPH3A
OAS1
OAS3
DTX1
CFAP73
Chromosome 12
Distance
LD
Coding
eQTL
RPH3A
OAS1
OAS3
OAS2
DTX1
RASAL1
CFAP73
DDX54

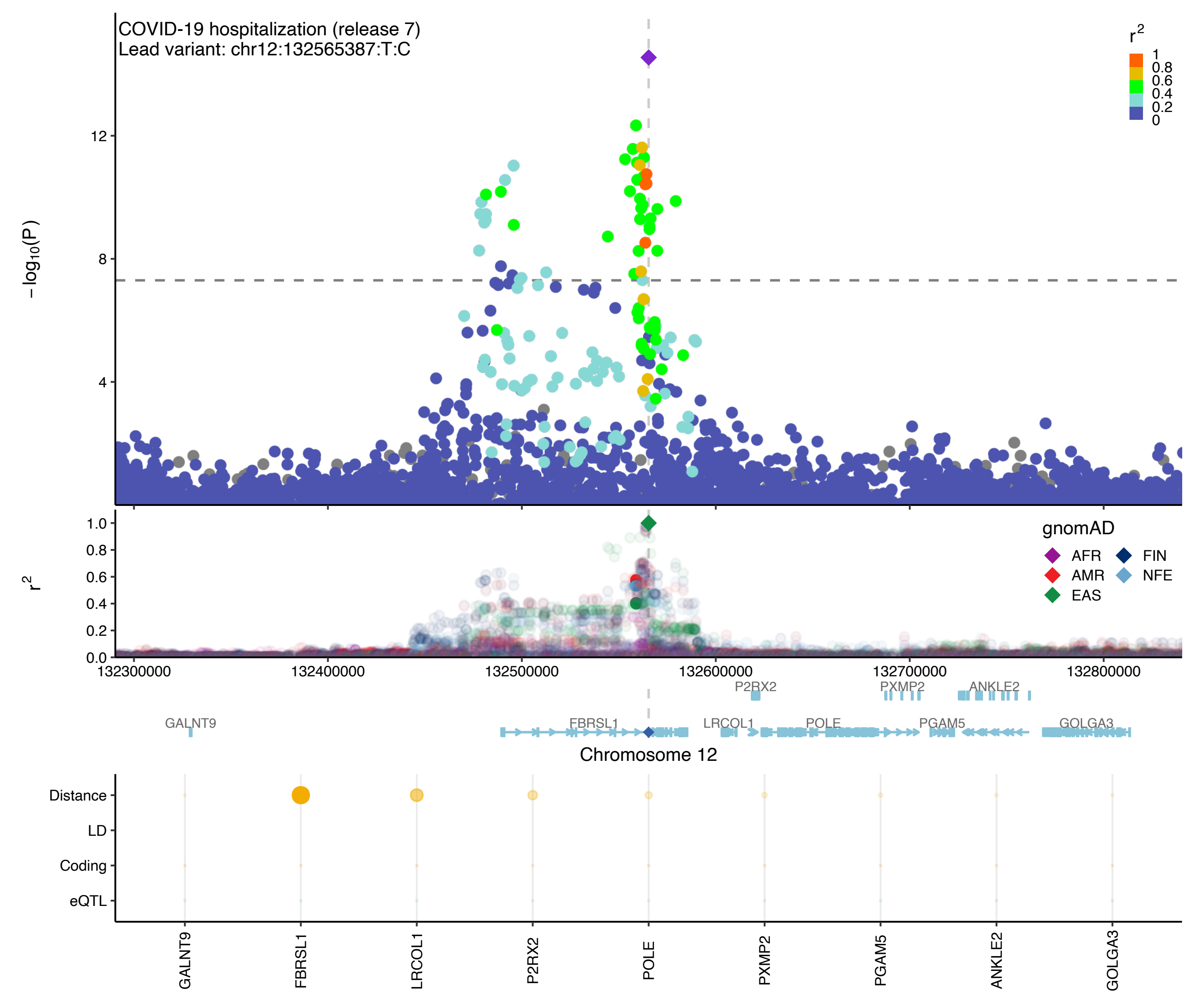

COVID-19 hospitalization (release 7)
Lead variant: chr12:132565387:T:C
r2
1
0.8
0.6
0.4
0.2
0
-log10(P)
12
8
4
r2
1.0
0.8
0.6
0.4
0.2
0.0
gnomAD
AFR
FIN
AMR
NFE
EAS
132300000
132400000
132500000
132600000
132700000
132800000
P2RX2
PXMP2
ANKLE2
GALNT9
FBRSL1
LRCOL1
POLE
PGAM5
GOLGA3
Chromosome 12
Distance
LD
Coding
eQTL
GALNT9
FBRSL1
LRCOL1
P2RX2
POLE
PXMP2
PGAM5
ANKLE2
GOLGA3

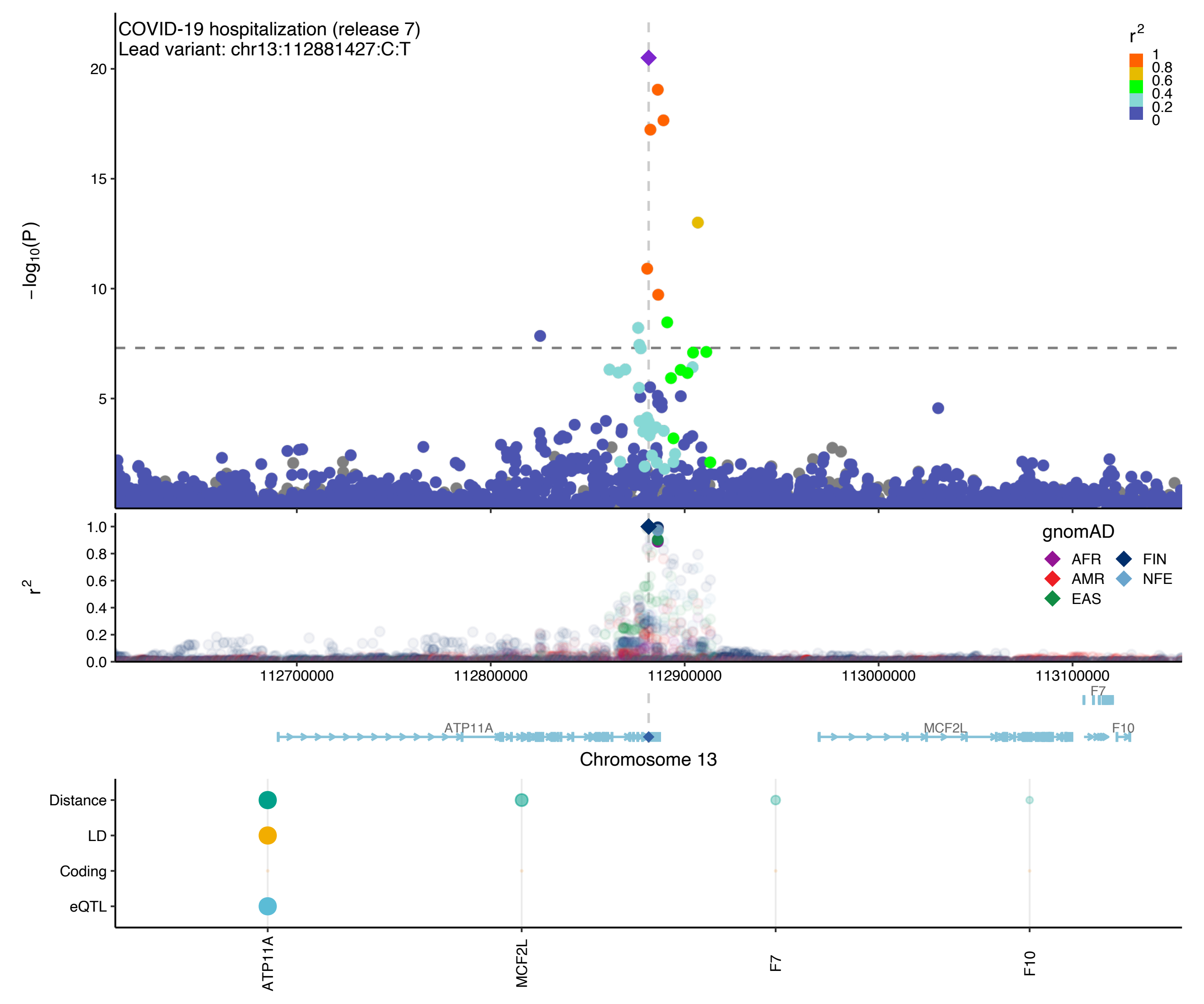
COVID-19 hospitalization (release 7)
Lead variant: chr13:112881427:C:T
−log10(P)
r2
gnomAD
AFR
AMR
EAS
FIN
NFE
112700000
112800000
112900000
113000000
113100000
F7
ATP11A
MCF2L
F10
Chromosome 13
Distance
LD
Coding
eQTL
ATP11A
MCF2L
F7
F10

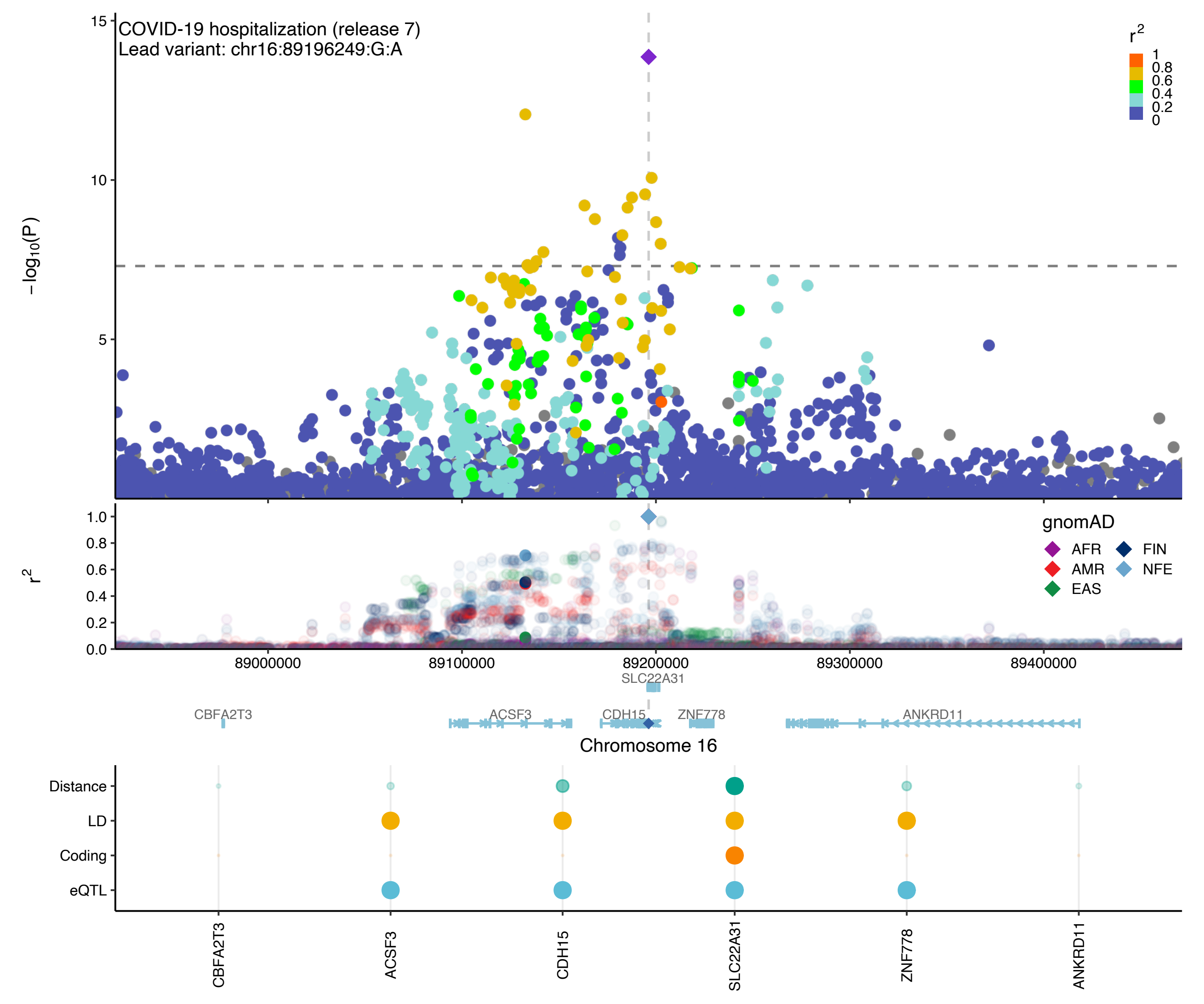
COVID-19 hospitalization (release 7)
Lead variant: chr16:89196249:G:A
$r^2$
1
0.8
0.6
0.4
0.2
0
-log10(P)
15
10
5
gnomAD
AFR
AMR
EAS
FIN
NFE
$r^2$
1.0
0.8
0.6
0.4
0.2
0.0
89000000
89100000
89200000
89300000
89400000
SLC22A31
CBFA2T3
ACSF3
CDH15
ZNF778
ANKRD11
Chromosome 16
Distance
LD
Coding
eQTL
CBFA2T3
ACSF3
CDH15
SLC22A31
ZNF778
ANKRD11

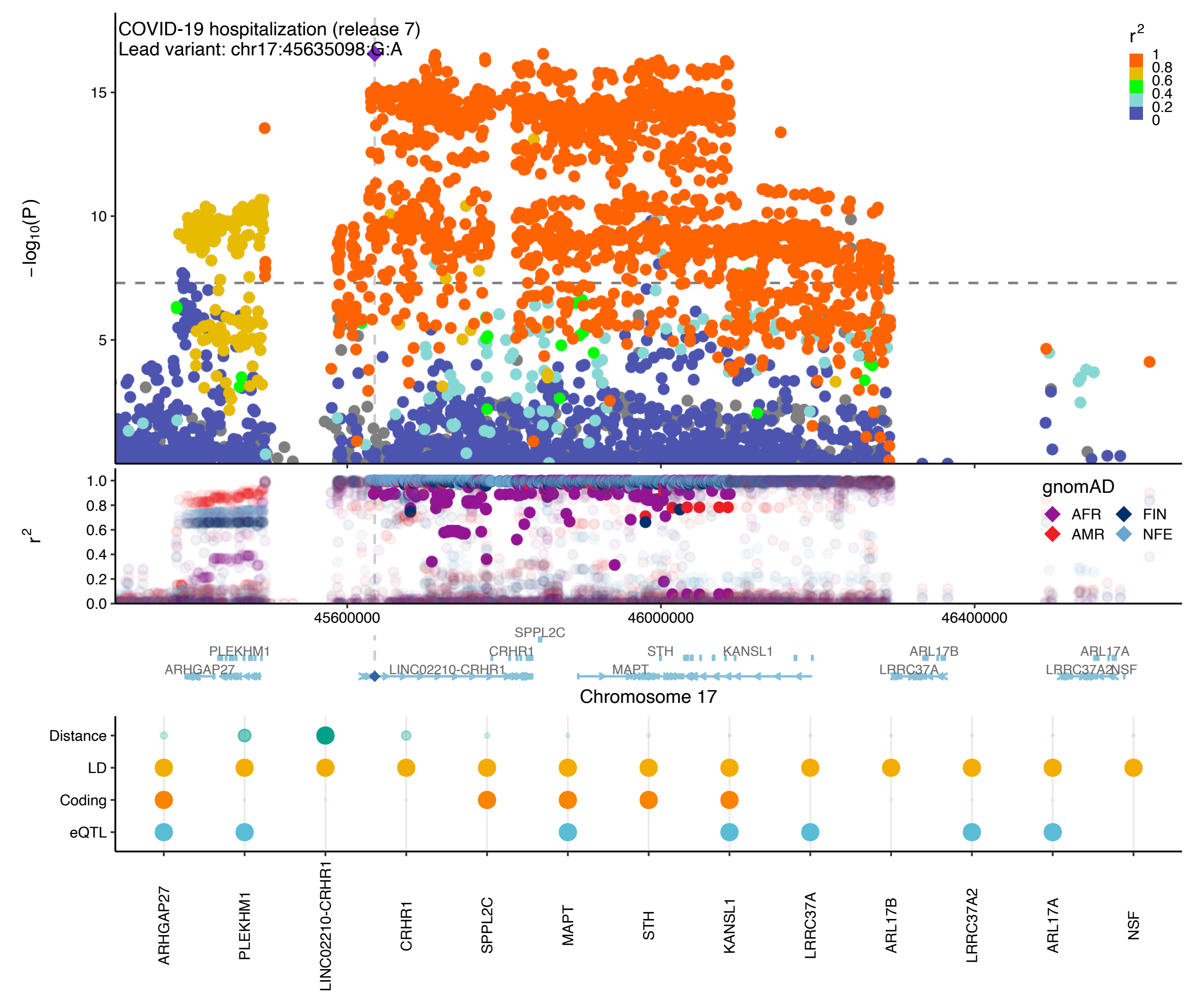

COVID-19 hospitalization (release 7)
Lead variant: chr17:45635098:G:A
−log10(P)
r2
gnomAD
AFR
AMR
FIN
NFE
45600000
46000000
46400000
Chromosome 17
ARHGAP27
PLEKHM1
LINC02210-CRHR1
CRHR1
SPPL2C
MAPT
STH
KANSL1
LRRC37A
ARL17B
LRRC37A2
ARL17A
NSF
Distance
LD
Coding
eQTL

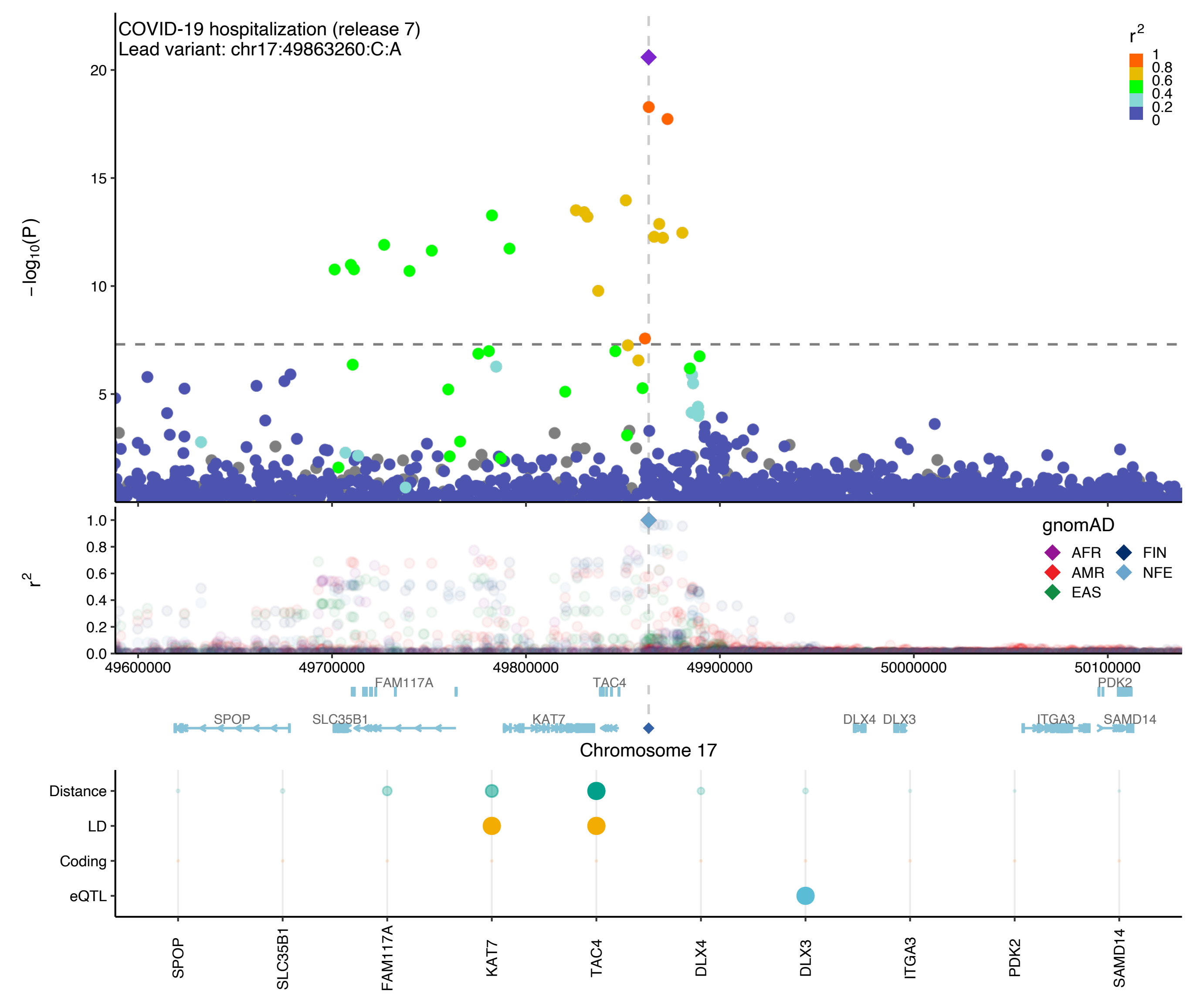

COVID-19 hospitalization (release 7)
Lead variant: chr17:49863260:C:A
−log10(P)
r2
gnomAD
AFR
AMR
EAS
FIN
NFE
49600000
49700000
49800000
49900000
50000000
50100000
FAM117A
TAC4
PDK2
SPOP
SLC35B1
KAT7
DLX4
DLX3
ITGA3
SAMD14
Chromosome 17
Distance
LD
Coding
eQTL
SPOP
SLC35B1
FAM117A
KAT7
TAC4
DLX4
DLX3
ITGA3
PDK2
SAMD14

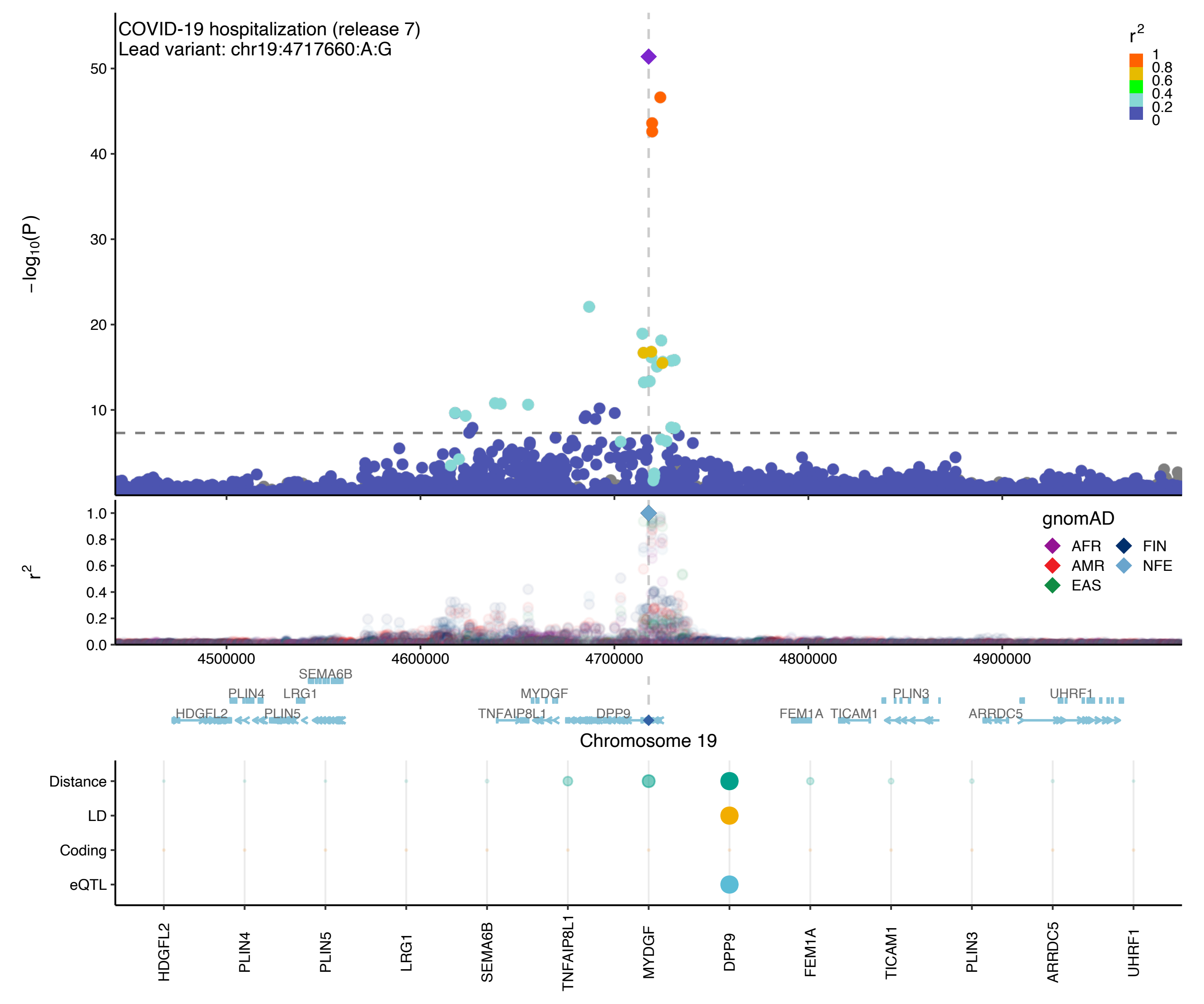
COVID-19 hospitalization (release 7)
Lead variant: chr19:4717660:A:G
−log10(P)
r2
1
0.8
0.6
0.4
0.2
0
gnomAD
AFR
AMR
EAS
FIN
NFE
4500000
4600000
4700000
4800000
4900000
SEMA6B
PLIN4
LRG1
HDGFL2
PLIN5
MYDGF
TNFAIP8L1
DPP9
FEM1A
TICAM1
PLIN3
UHRF1
ARRDC5
Chromosome 19
Distance
LD
Coding
eQTL
HDGFL2
PLIN4
PLIN5
LRG1
SEMA6B
TNFAIP8L1
MYDGF
DPP9
FEM1A
TICAM1
PLIN3
ARRDC5
UHRF1

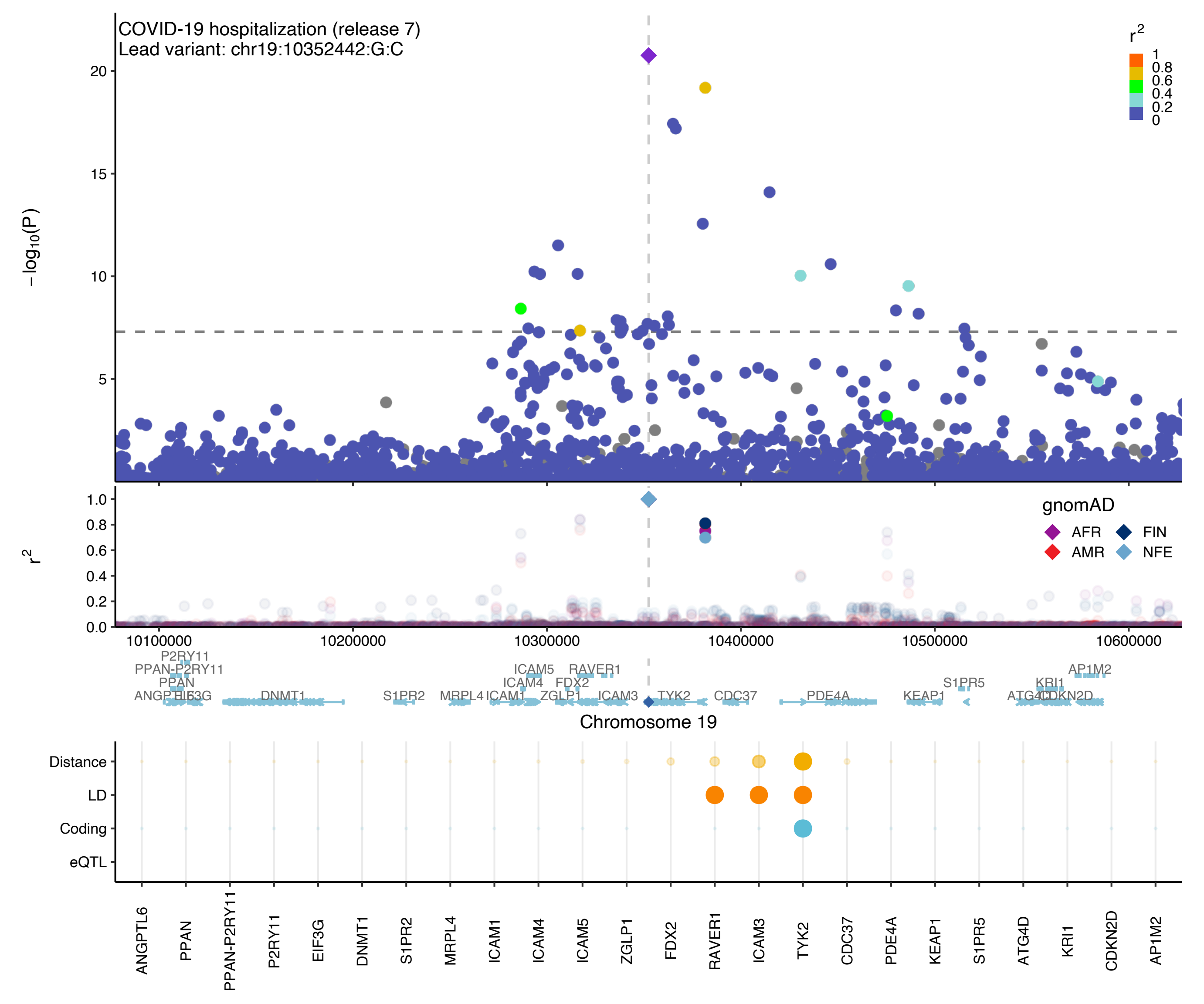

COVID-19 hospitalization (release 7)
Lead variant: chr19:10352442:G:C
−log10(P)
r2
gnomAD
AFR
FIN
AMR
NFE
Chromosome 19
Distance
LD
Coding
eQTL
ANGPTL6
PPAN
PPAN-P2RY11
P2RY11
EIF3G
DNMT1
S1PR2
MRPL4
ICAM1
ICAM4
ICAM5
ZGLP1
FDX2
RAVER1
ICAM3
TYK2
CDC37
PDE4A
KEAP1
S1PR5
ATG4D
KRI1
CDKN2D
AP1M2

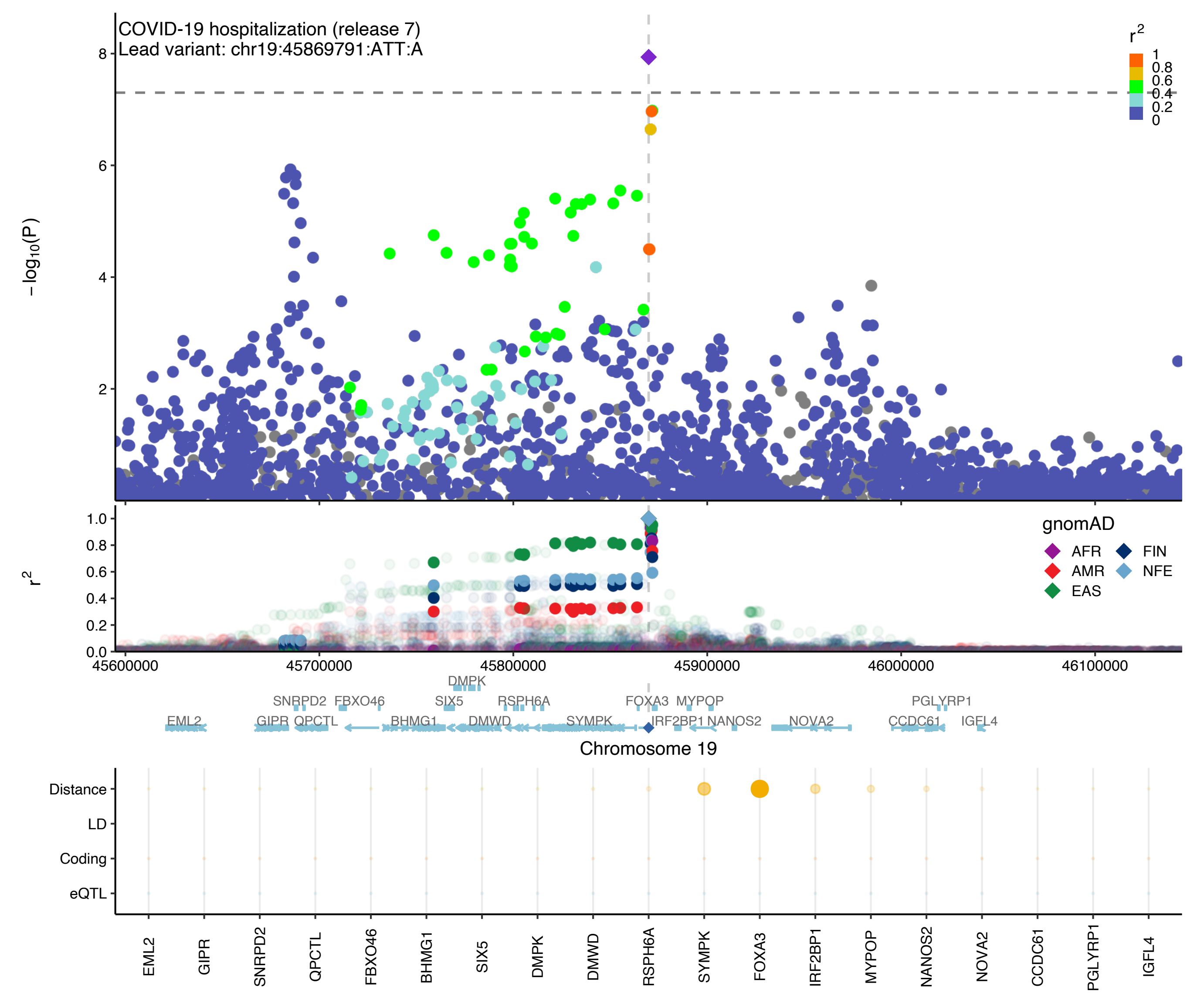

COVID-19 hospitalization (release 7)
Lead variant: chr19:45869791:ATT:A
$-\log_{10}(P)$
$r^2$
1
0.8
0.6
0.4
0.2
0
gnomAD
AFR
AMR
EAS
FIN
NFE
45600000
45700000
45800000
45900000
46000000
46100000
DMPK
SNRPD2
FBXO46
SIX5
RSPH6A
FOXA3
MYPOP
PGLYRP1
EML2
GIPR
QPCTL
BHMG1
DMWD
SYMPK
IRF2BP1
NANOS2
NOVA2
CCDC61
IGFL4
Chromosome 19
Distance
LD
Coding
eQTL
EML2
GIPR
SNRPD2
QPCTL
FBXO46
BHMG1
SIX5
DMPK
DMWD
RSPH6A
SYMPK
FOXA3
IRF2BP1
MYPOP
NANOS2
NOVA2
CCDC61
PGLYRP1
IGFL4

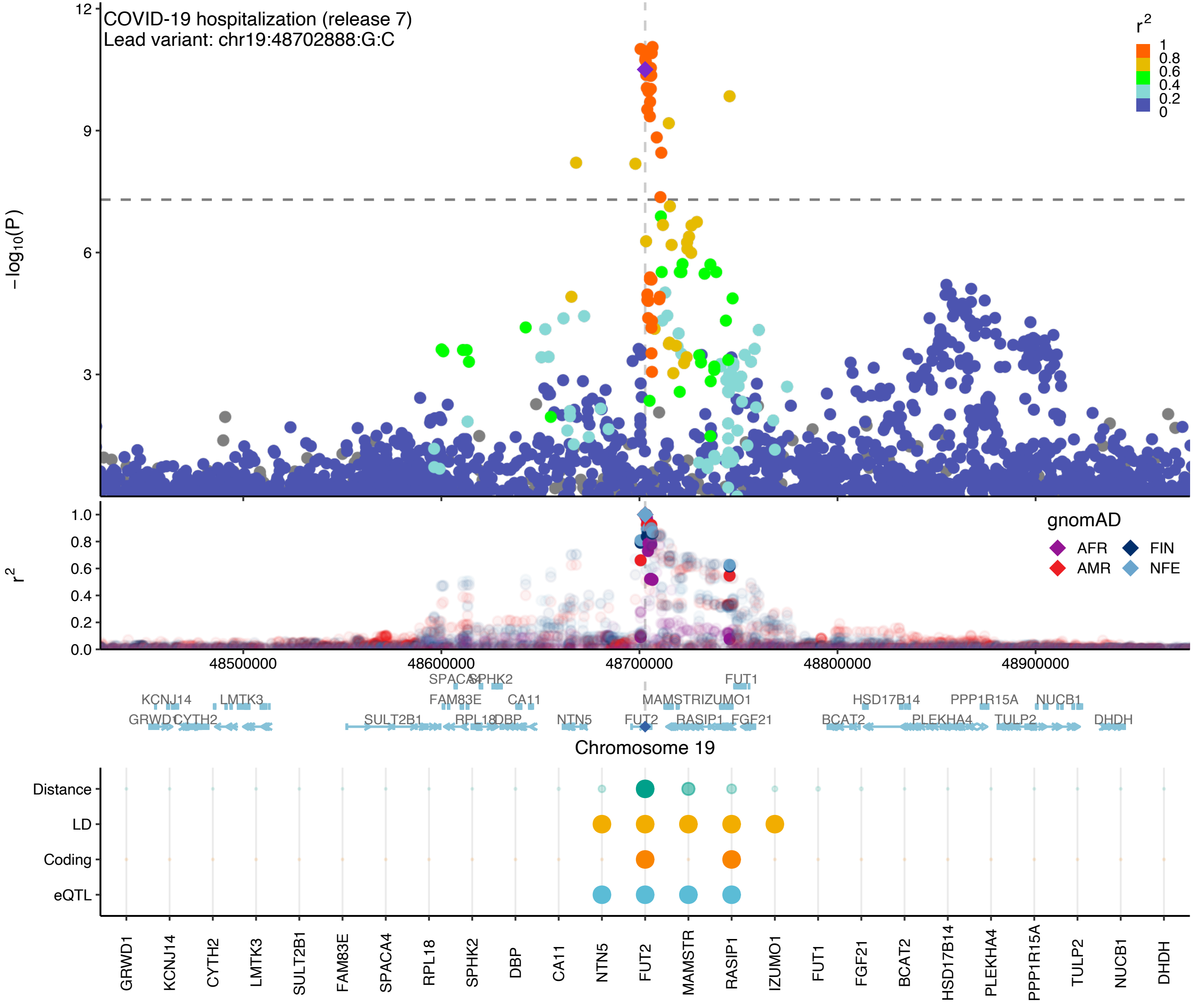

COVID-19 hospitalization (release 7)
Lead variant: chr19:48702888:G:C
−log10(P)
r2
1
0.8
0.6
0.4
0.2
0
gnomAD
AFR
AMR
FIN
NFE
48500000
48600000
48700000
48800000
48900000
Chromosome 19
Distance
LD
Coding
eQTL
GRWD1
KCNJ14
CYTH2
LMTK3
SULT2B1
FAM83E
SPACA4
RPL18
SPHK2
DBP
CA11
NTN5
FUT2
MAMSTR
RASIP1
IZUMO1
FUT1
FGF21
BCAT2
HSD17B14
PLEKHA4
PPP1R15A
TULP2
NUCB1
DHDH

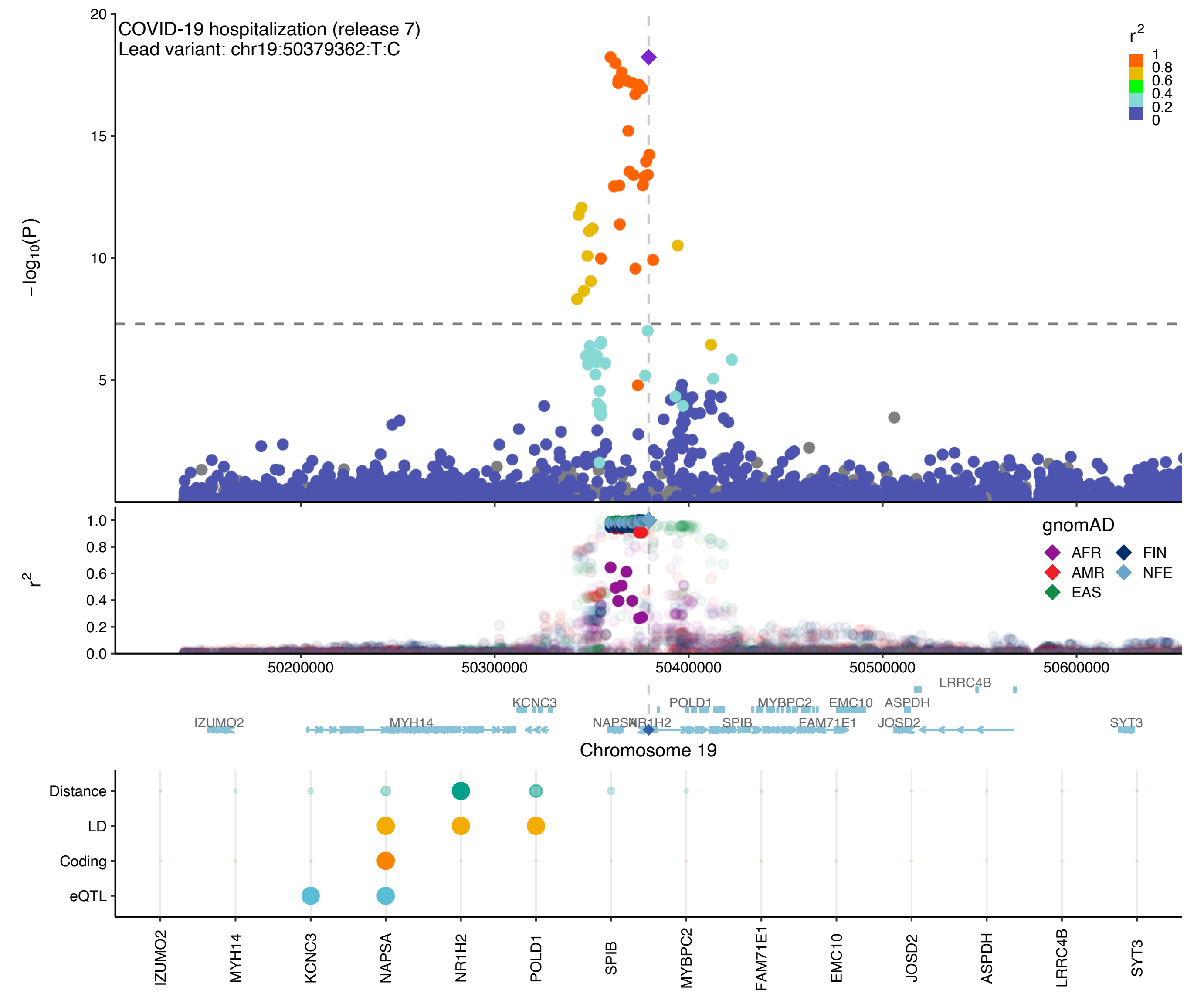

COVID-19 hospitalization (release 7)
Lead variant: chr19:50379362:T:C
−log10(P)
r2
1
0.8
0.6
0.4
0.2
0
20
15
10
5
1.0
0.8
0.6
0.4
0.2
0.0
gnomAD
AFR
AMR
EAS
FIN
NFE
50200000
50300000
50400000
50500000
50600000
LRRC4B
KCNC3
POLD1
MYBPC2
EMC10
ASPDH
IZUMO2
MYH14
NAPSA
NR1H2
SPIB
FAM71E1
JOSD2
SYT3
Chromosome 19
Distance
LD
Coding
eQTL
IZUMO2
MYH14
KCNC3
NAPSA
NR1H2
POLD1
SPIB
MYBPC2
FAM71E1
EMC10
JOSD2
ASPDH
LRRC4B
SYT3

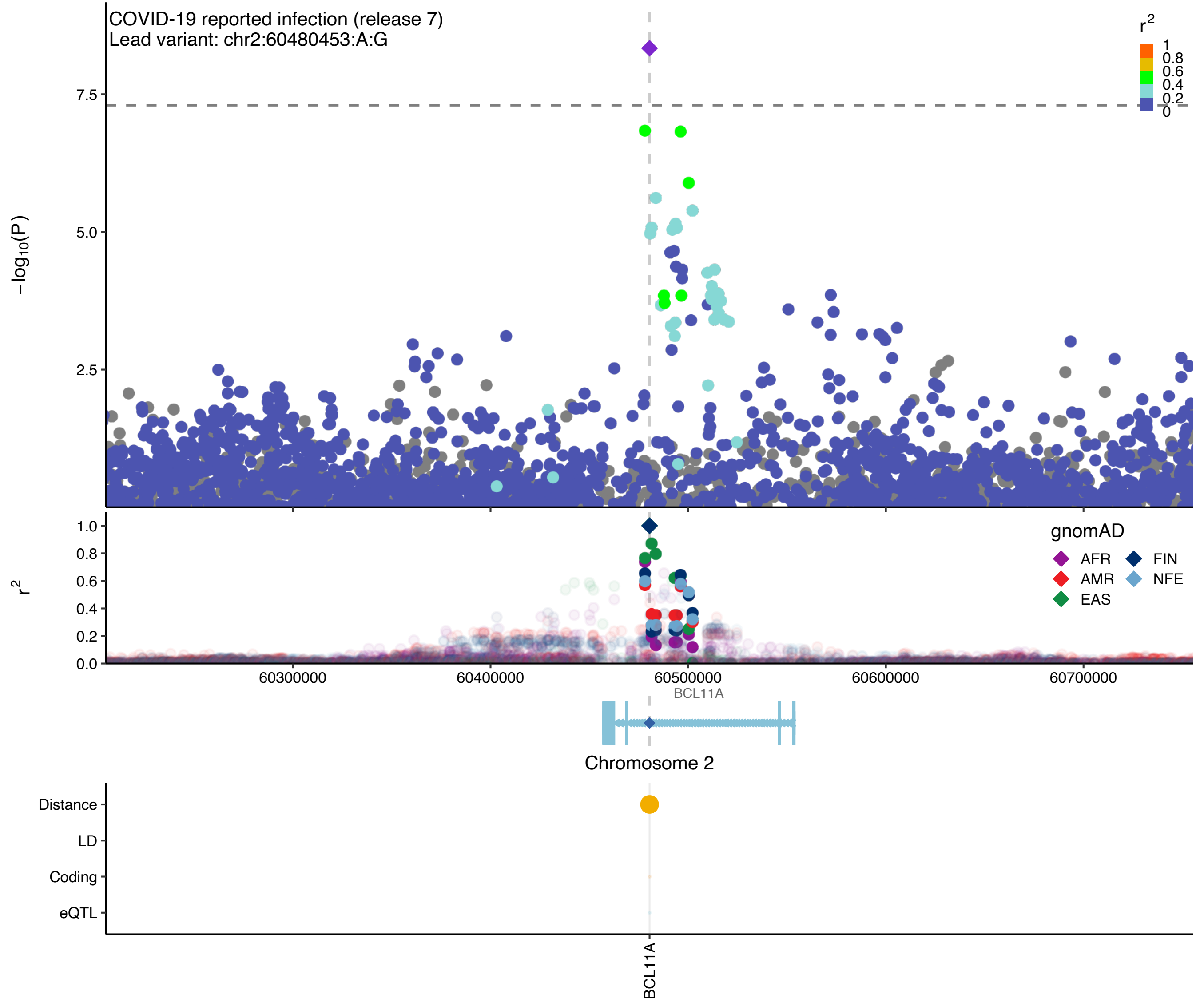
COVID-19 reported infection (release 7)
Lead variant: chr2:60480453:A:G
$-\log_{10}(P)$
7.5
5.0
2.5
$r^2$
1
0.8
0.6
0.4
0.2
0
1.0
0.8
0.6
0.4
0.2
0.0
gnomAD
AFR
AMR
EAS
FIN
NFE
60300000
60400000
60500000
60600000
60700000
BCL11A
Chromosome 2
Distance
LD
Coding
eQTL
BCL11A

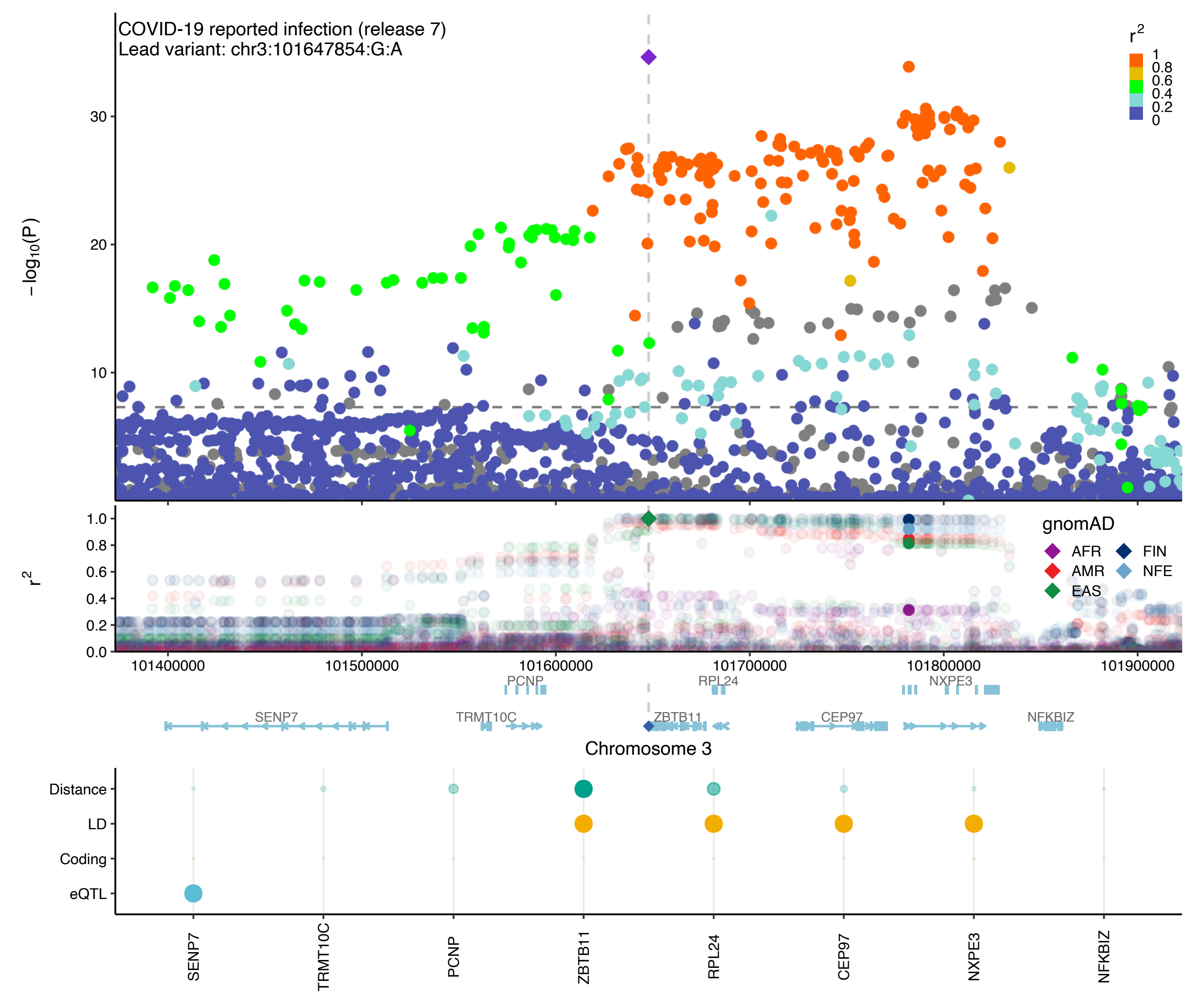

COVID-19 reported infection (release 7)
Lead variant: chr3:101647854:G:A
r2
1
0.8
0.6
0.4
0.2
0
−log10(P)
30
20
10
r2
1.0
0.8
0.6
0.4
0.2
0.0
gnomAD
AFR
FIN
AMR
NFE
EAS
101400000
101500000
101600000
101700000
101800000
101900000
PCNP
RPL24
NXPE3
SENP7
TRMT10C
ZBTB11
CEP97
NFKBIZ
Chromosome 3
Distance
LD
Coding
eQTL
SENP7
TRMT10C
PCNP
ZBTB11
RPL24
CEP97
NXPE3
NFKBIZ

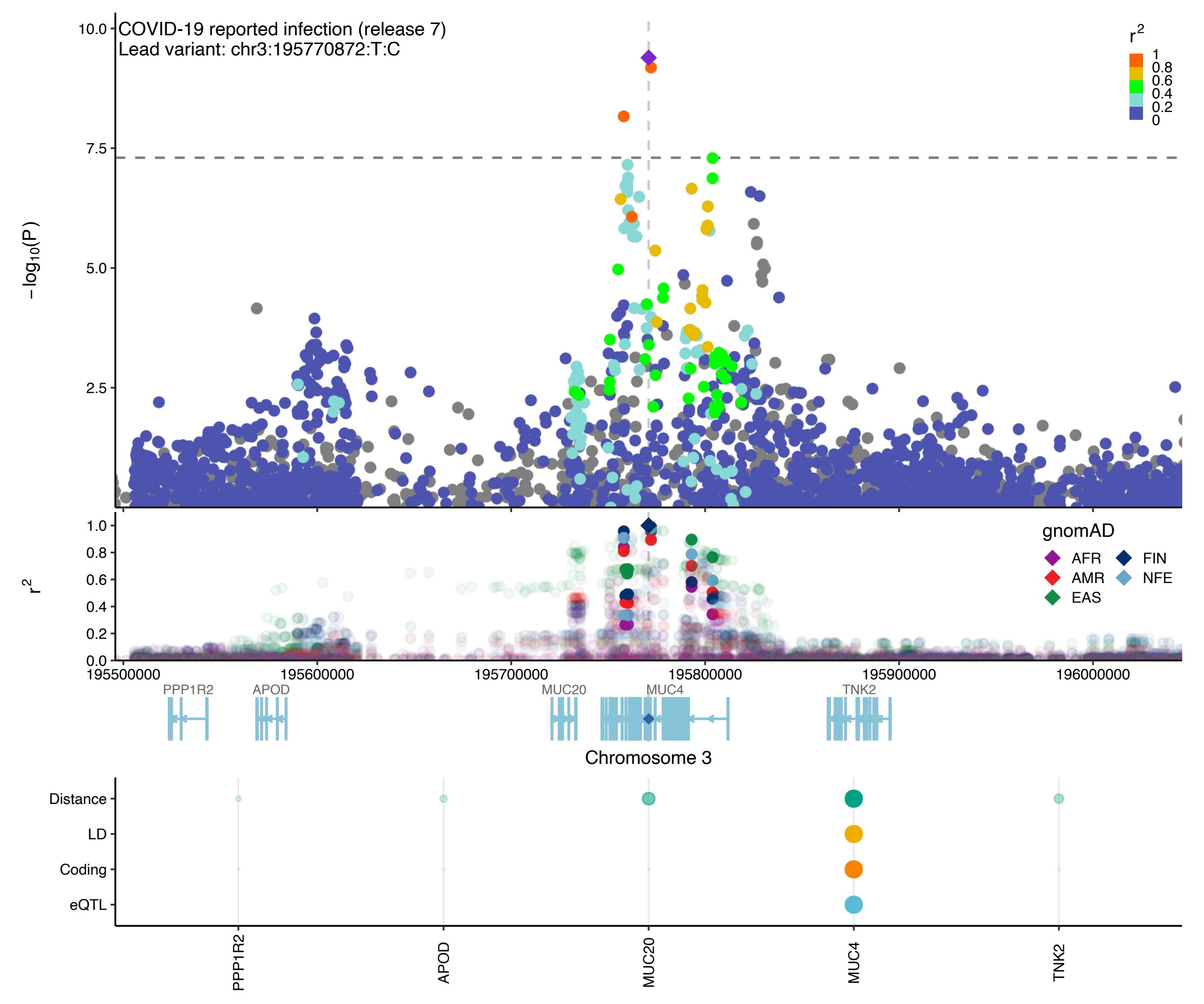
COVID-19 reported infection (release 7)
Lead variant: chr3:195770872:T:C
−log10(P)
r2
1
0.8
0.6
0.4
0.2
0
gnomAD
AFR
AMR
EAS
FIN
NFE
195500000
195600000
195700000
195800000
195900000
196000000
PPP1R2
APOD
MUC20
MUC4
TNK2
Chromosome 3
Distance
LD
Coding
eQTL
PPP1R2
APOD
MUC20
MUC4
TNK2

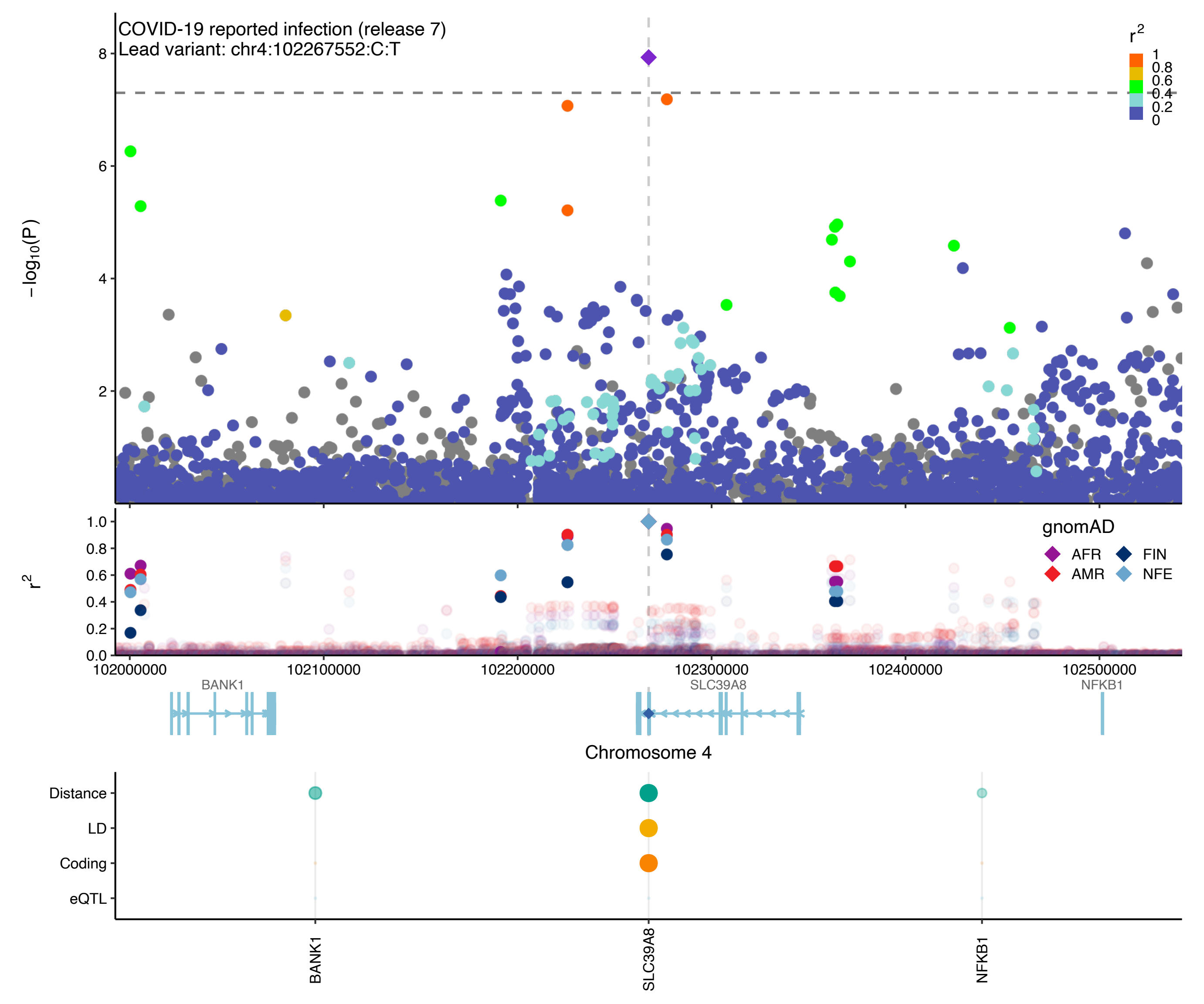

COVID-19 reported infection (release 7)
Lead variant: chr4:102267552:C:T
−log10(P)
r2
1
0.8
0.6
0.4
0.2
0
gnomAD
AFR
FIN
AMR
NFE
102000000
102100000
102200000
102300000
102400000
102500000
BANK1
SLC39A8
NFKB1
Chromosome 4
Distance
LD
Coding
eQTL
BANK1
SLC39A8
NFKB1

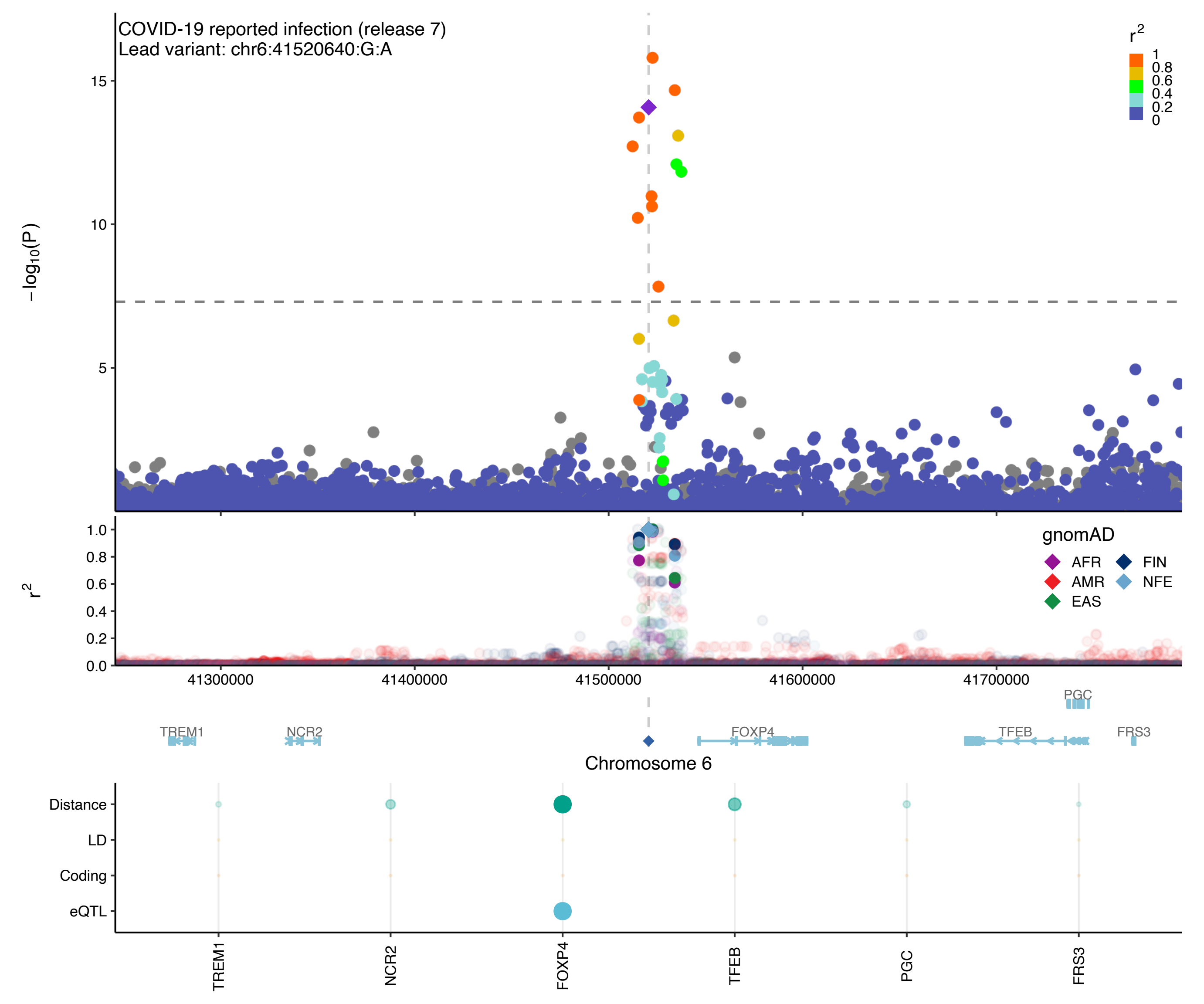
COVID-19 reported infection (release 7)
Lead variant: chr6:41520640:G:A
−log10(P)
15
10
5
r2
1
0.8
0.6
0.4
0.2
0
1.0
0.8
0.6
0.4
0.2
0.0
gnomAD
AFR
AMR
EAS
FIN
NFE
41300000
41400000
41500000
41600000
41700000
PGC
TREM1
NCR2
FOXP4
TFEB
FRS3
Chromosome 6
Distance
LD
Coding
eQTL
TREM1
NCR2
FOXP4
TFEB
PGC
FRS3

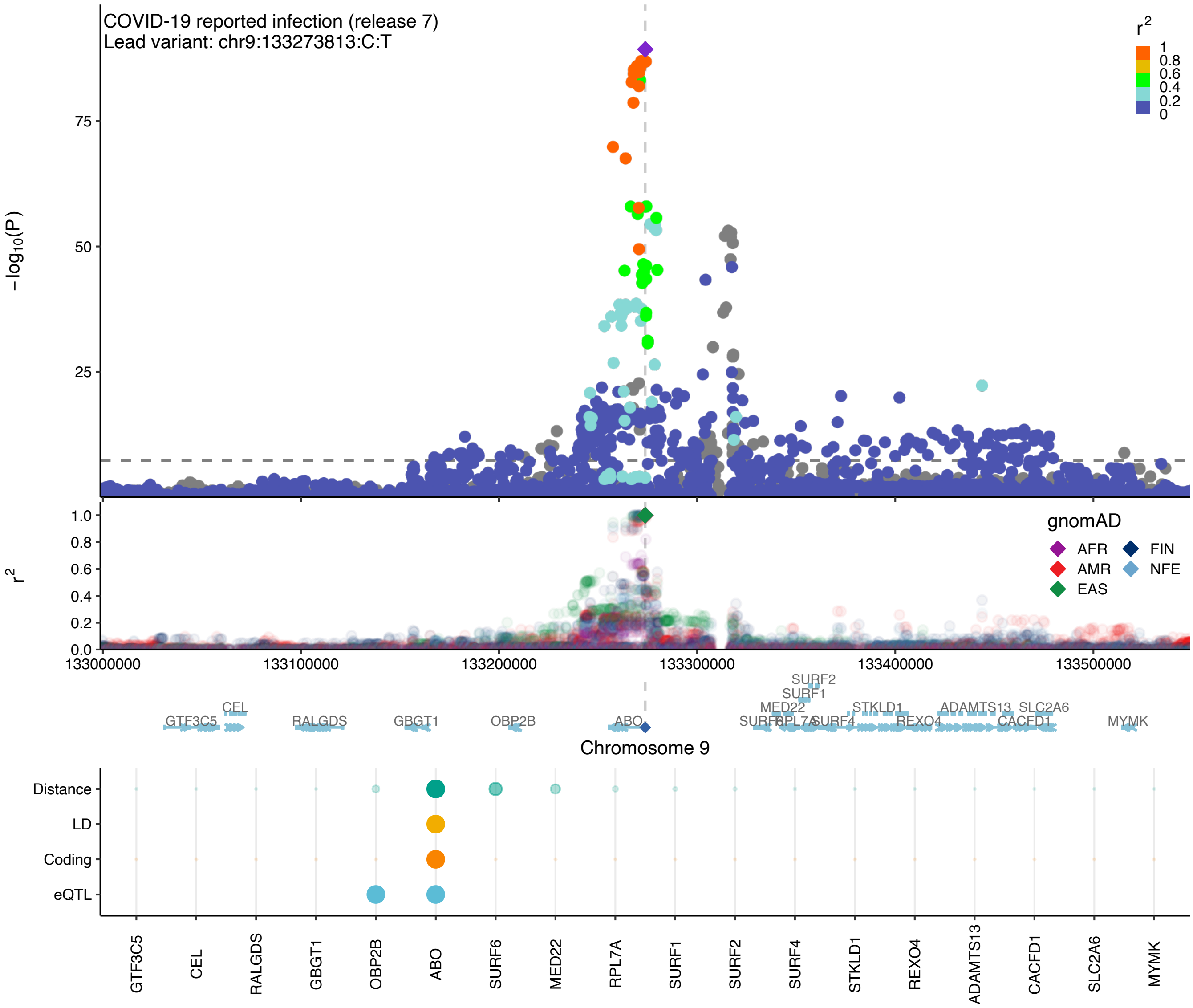

COVID-19 reported infection (release 7)
Lead variant: chr9:133273813:C:T
−log10(P)
75
50
25
r2
1
0.8
0.6
0.4
0.2
0
r2
1.0
0.8
0.6
0.4
0.2
0.0
gnomAD
AFR
FIN
AMR
NFE
EAS
133000000
133100000
133200000
133300000
133400000
133500000
SURF2
SURF1
MED22
STKLD1
ADAMTS13
SLC2A6
CEL
GTF3C5
RALGDS
GBGT1
OBP2B
ABO
SURF6
RPL7A
SURF4
REXO4
CACFD1
MYMK
Chromosome 9
Distance
LD
Coding
eQTL
GTF3C5
CEL
RALGDS
GBGT1
OBP2B
ABO
SURF6
MED22
RPL7A
SURF1
SURF2
SURF4
STKLD1
REXO4
ADAMTS13
CACFD1
SLC2A6
MYMK

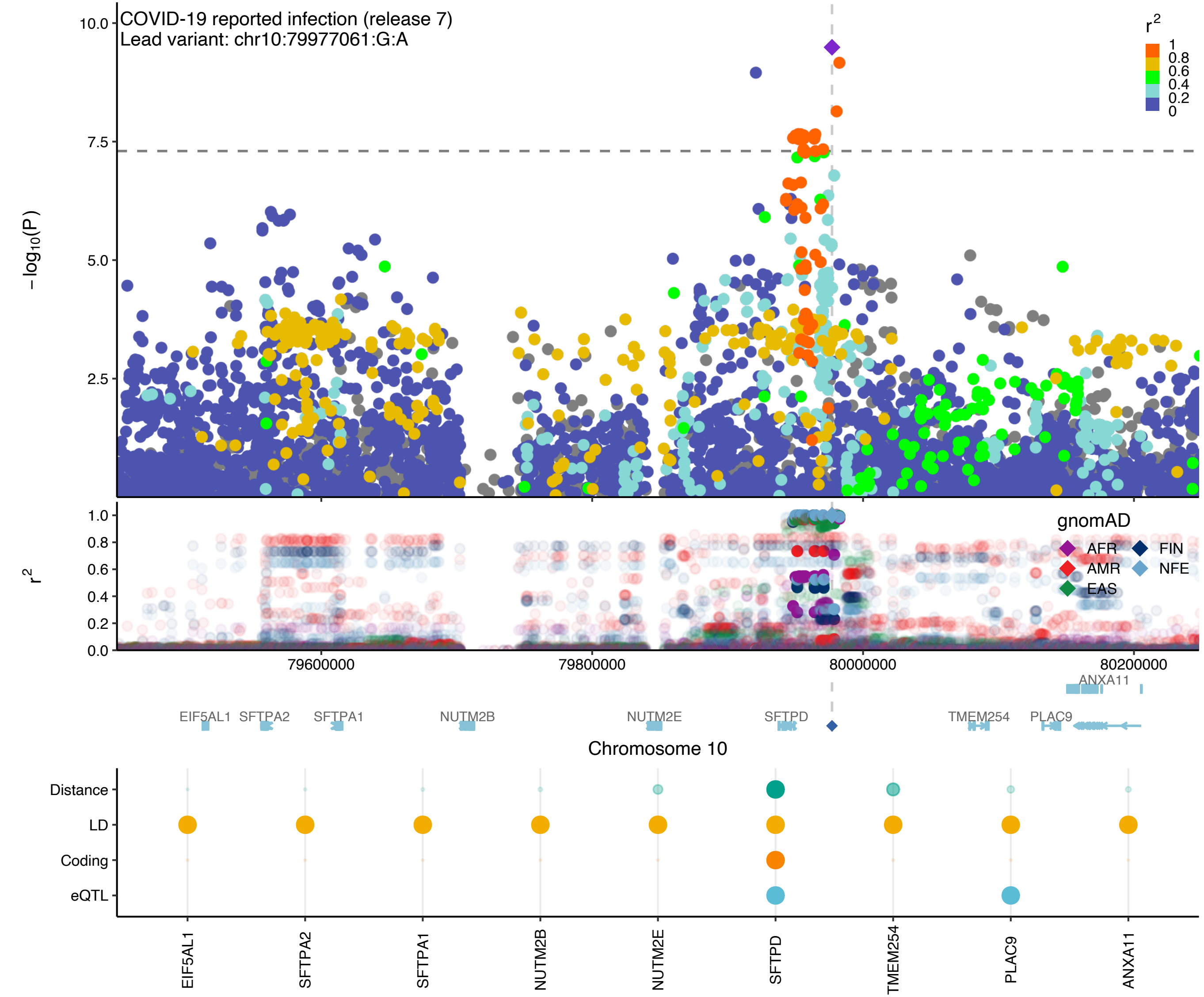

COVID-19 reported infection (release 7)
Lead variant: chr10:79977061:G:A
r2
1
0.8
0.6
0.4
0.2
0
−log10(P)
10.0
7.5
5.0
2.5
r2
1.0
0.8
0.6
0.4
0.2
0.0
gnomAD
AFR
FIN
AMR
NFE
EAS
79600000
79800000
80000000
80200000
ANXA11
EIF5AL1
SFTPA2
SFTPA1
NUTM2B
NUTM2E
SFTPD
TMEM254
PLAC9
Chromosome 10
Distance
LD
Coding
eQTL
EIF5AL1
SFTPA2
SFTPA1
NUTM2B
NUTM2E
SFTPD
TMEM254
PLAC9
ANXA11

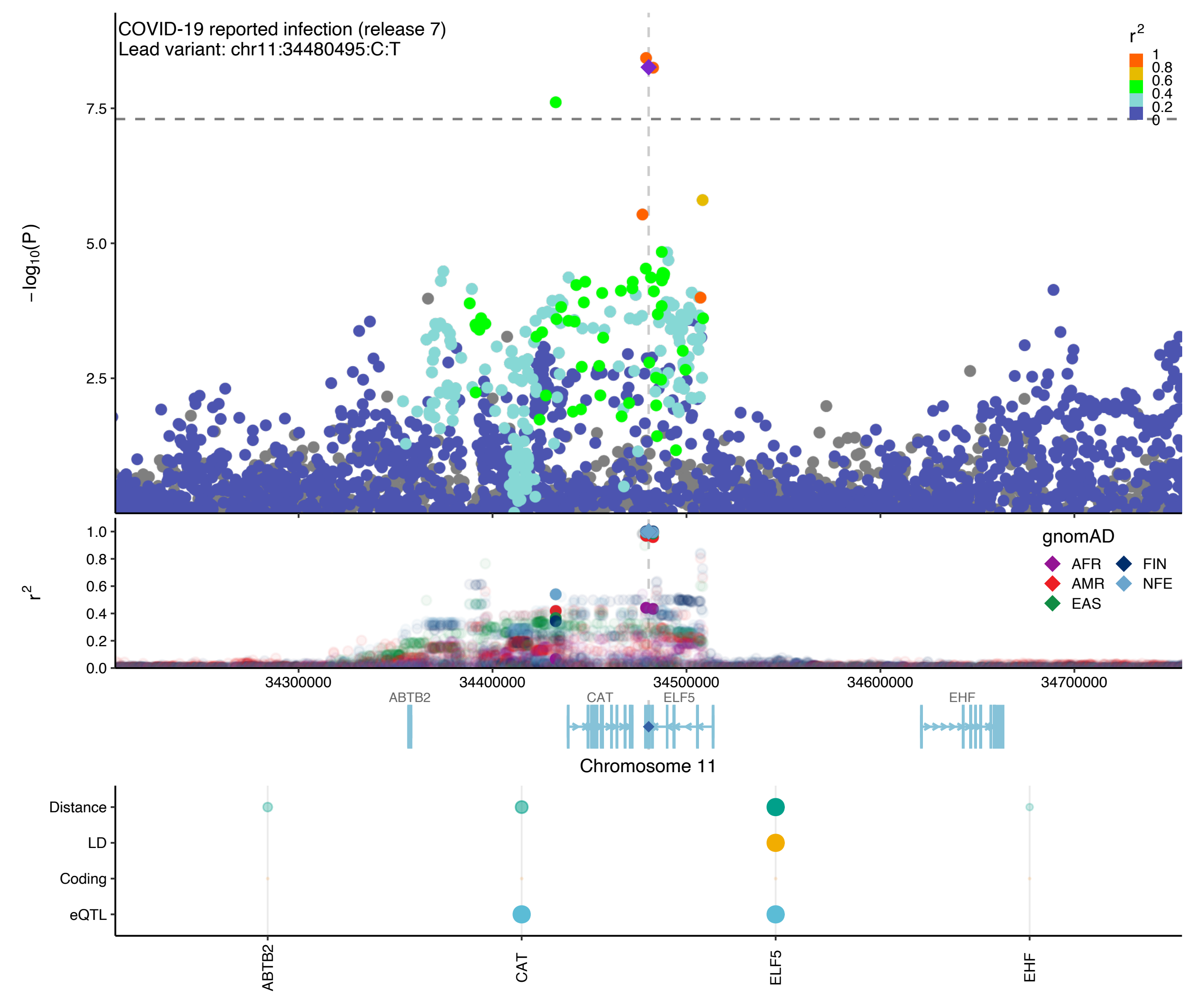

COVID-19 reported infection (release 7)
Lead variant: chr11:34480495:C:T
-log10(P)
7.5
5.0
2.5
r2
1
0.8
0.6
0.4
0.2
0
1.0
0.8
0.6
0.4
0.2
0.0
gnomAD
AFR
AMR
EAS
FIN
NFE
34300000
34400000
34500000
34600000
34700000
ABTB2
CAT
ELF5
EHF
Chromosome 11
Distance
LD
Coding
eQTL
ABTB2
CAT
ELF5
EHF

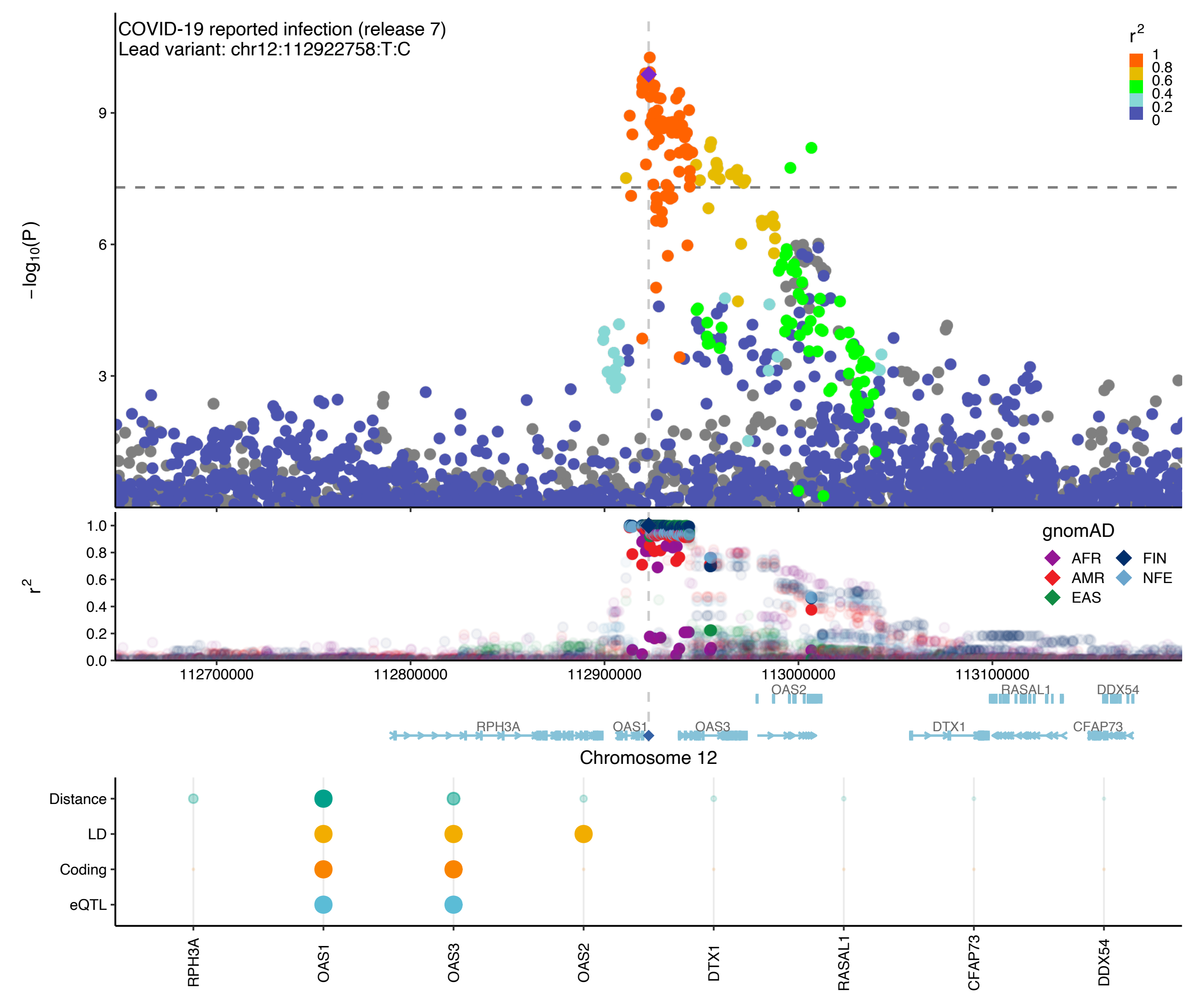

COVID-19 reported infection (release 7)
Lead variant: chr12:112922758:T:C
−log10(P)
r2
gnomAD
AFR
AMR
EAS
FIN
NFE
112700000
112800000
112900000
113000000
113100000
OAS2
RASAL1
DDX54
RPH3A
OAS1
OAS3
DTX1
CFAP73
Chromosome 12
Distance
LD
Coding
eQTL

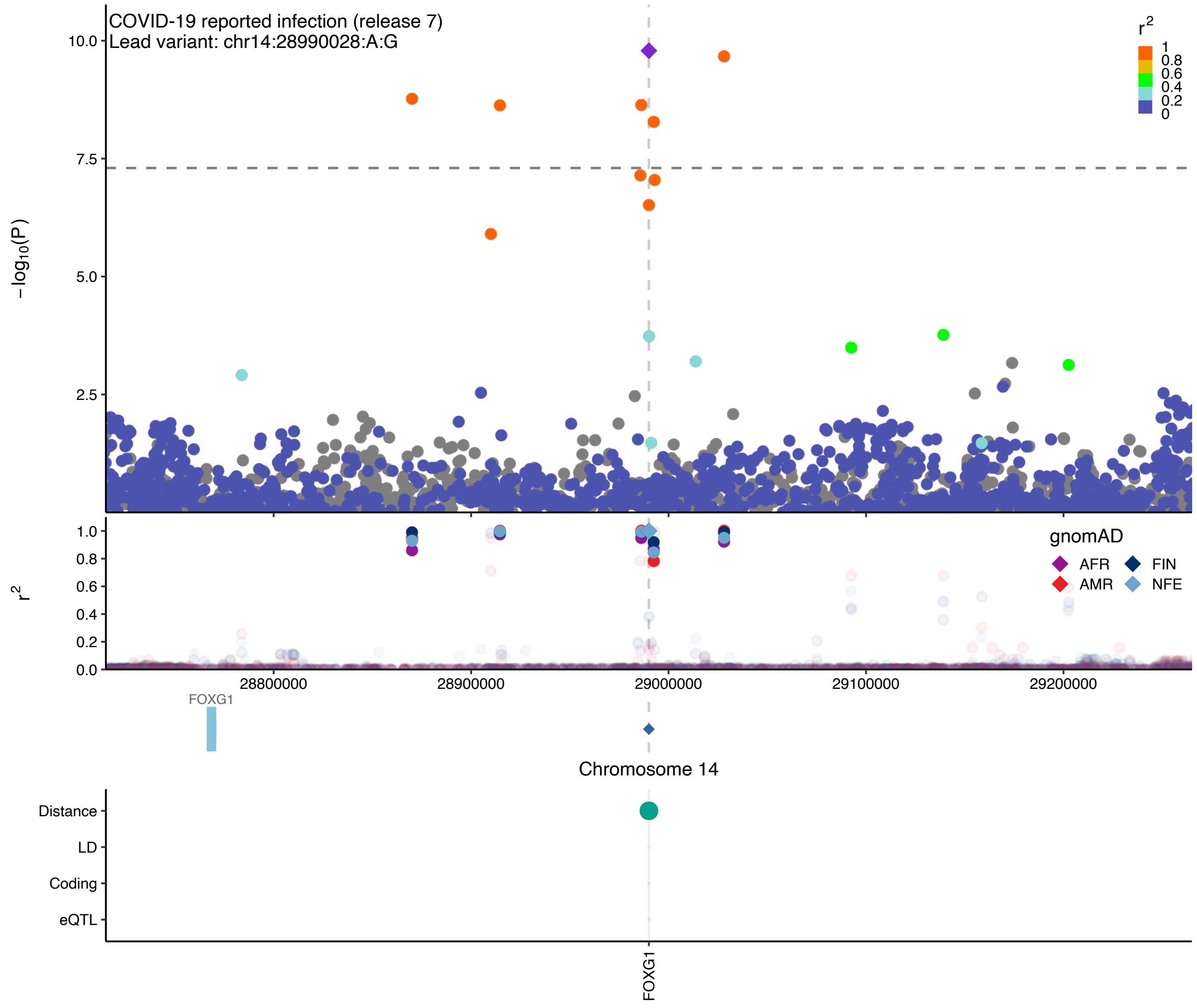

COVID-19 reported infection (release 7)
Lead variant: chr14:28990028:A:G
−log10(P)
10.0
7.5
5.0
2.5
r2
1
0.8
0.6
0.4
0.2
0
1.0
0.8
0.6
0.4
0.2
0.0
gnomAD
AFR
FIN
AMR
NFE
28800000
28900000
29000000
29100000
29200000
FOXG1
Chromosome 14
Distance
LD
Coding
eQTL
FOXG1

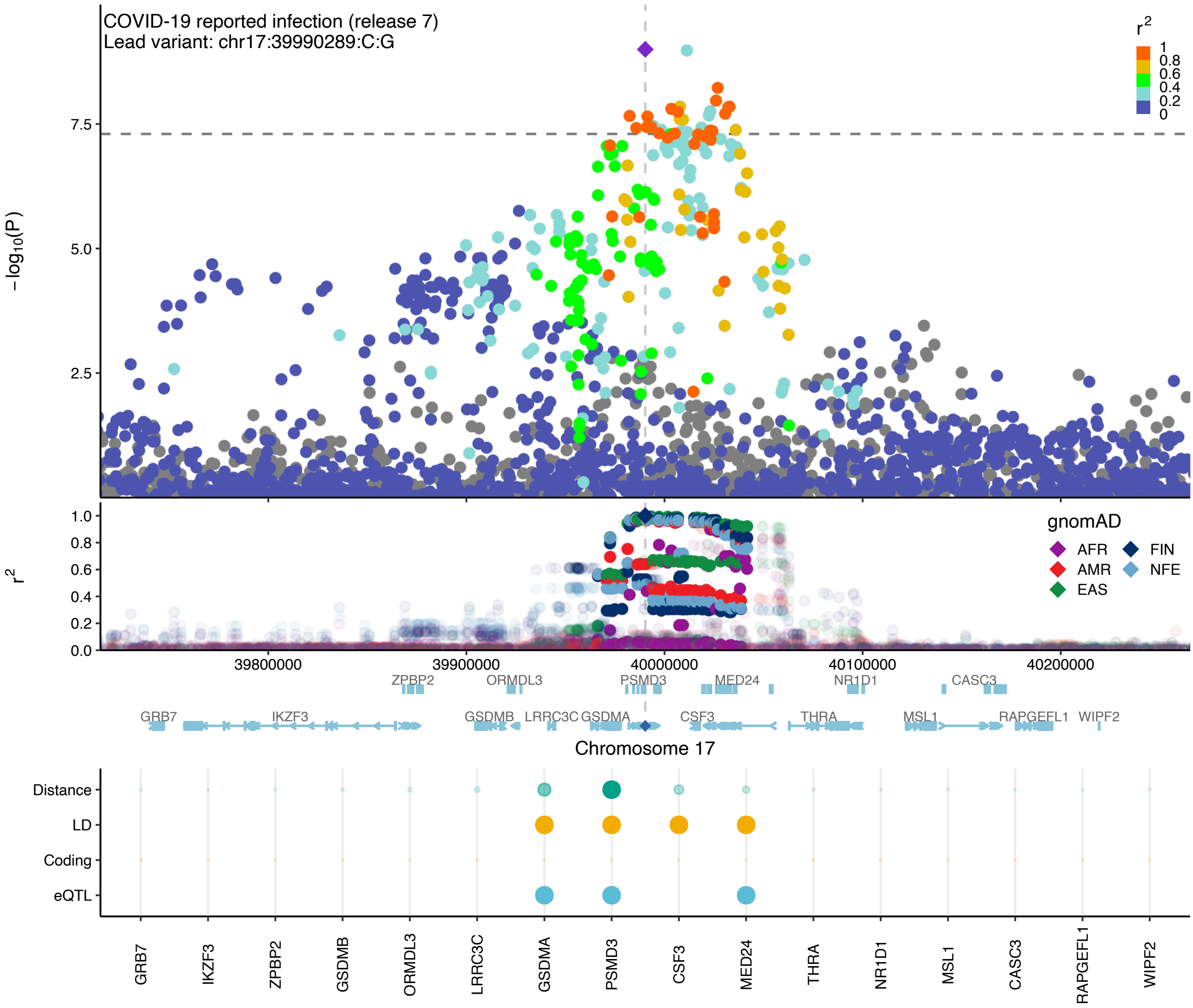

COVID-19 reported infection (release 7)
Lead variant: chr17:39990289:C:G
r²
1
0.8
0.6
0.4
0.2
0
−log10(P)
7.5
5.0
2.5
r²
1.0
0.8
0.6
0.4
0.2
0.0
gnomAD
AFR
FIN
AMR
NFE
EAS
39800000
39900000
40000000
40100000
40200000
ZPBP2
ORMDL3
PSMD3
MED24
NR1D1
CASC3
GRB7
IKZF3
GSDMB
LRRC3C
GSDMA
CSF3
THRA
MSL1
RAPGEFL1
WIPF2
Chromosome 17
Distance
LD
Coding
eQTL
GRB7
IKZF3
ZPBP2
GSDMB
ORMDL3
LRRC3C
GSDMA
PSMD3
CSF3
MED24
THRA
NR1D1
MSL1
CASC3
RAPGEFL1
WIPF2

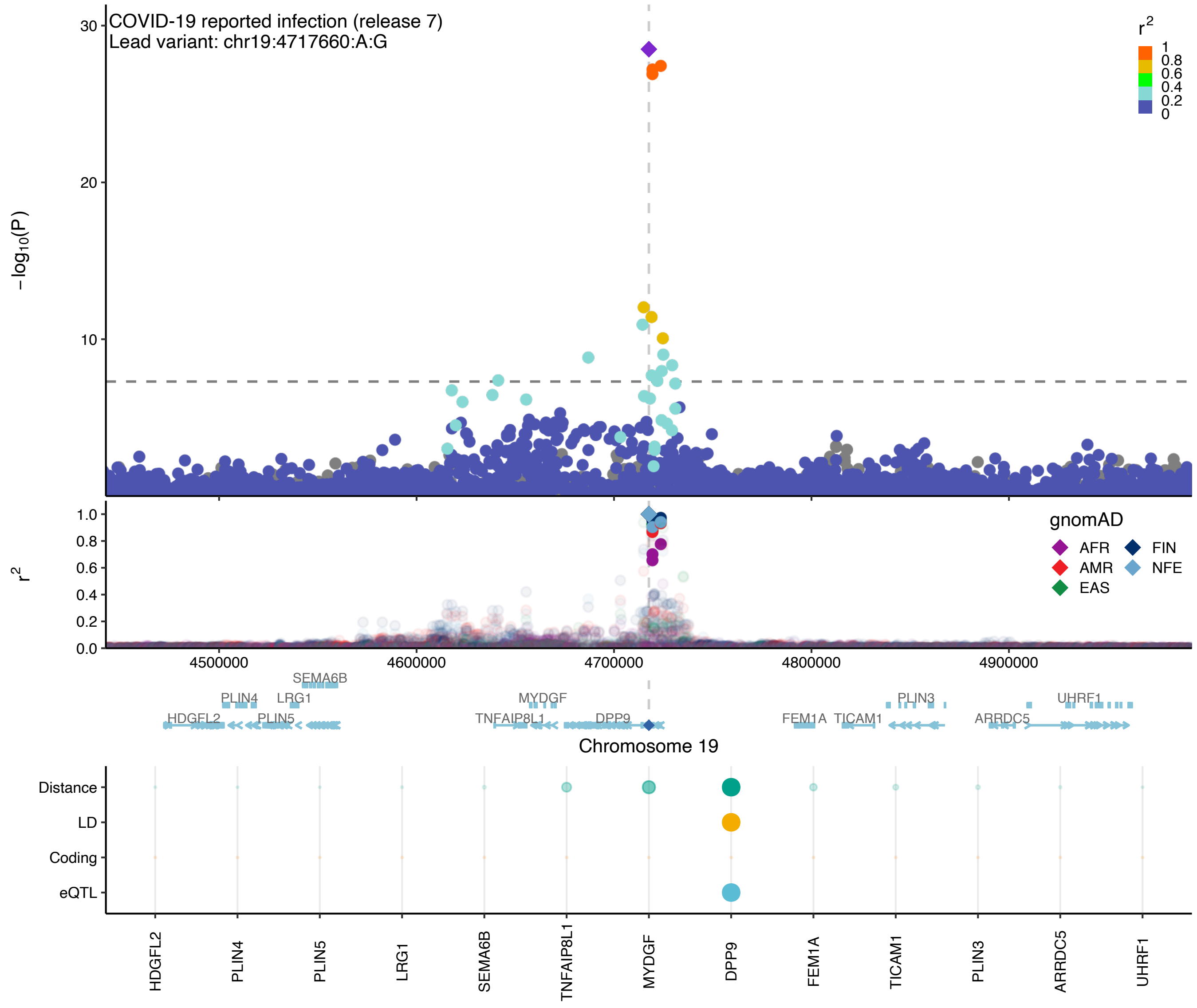

COVID-19 reported infection (release 7)
Lead variant: chr19:4717660:A:G
r2
1
0.8
0.6
0.4
0.2
0
-log10(P)
30
20
10
gnomAD
AFR
FIN
AMR
NFE
EAS
r2
1.0
0.8
0.6
0.4
0.2
0.0
4500000
4600000
4700000
4800000
4900000
SEMA6B
PLIN4
LRG1
MYDGF
PLIN3
UHRF1
HDGFL2
PLIN5
TNFAIP8L1
DPP9
FEM1A
TICAM1
ARRDC5
Chromosome 19
Distance
LD
Coding
eQTL
HDGFL2
PLIN4
PLIN5
LRG1
SEMA6B
TNFAIP8L1
MYDGF
DPP9
FEM1A
TICAM1
PLIN3
ARRDC5
UHRF1

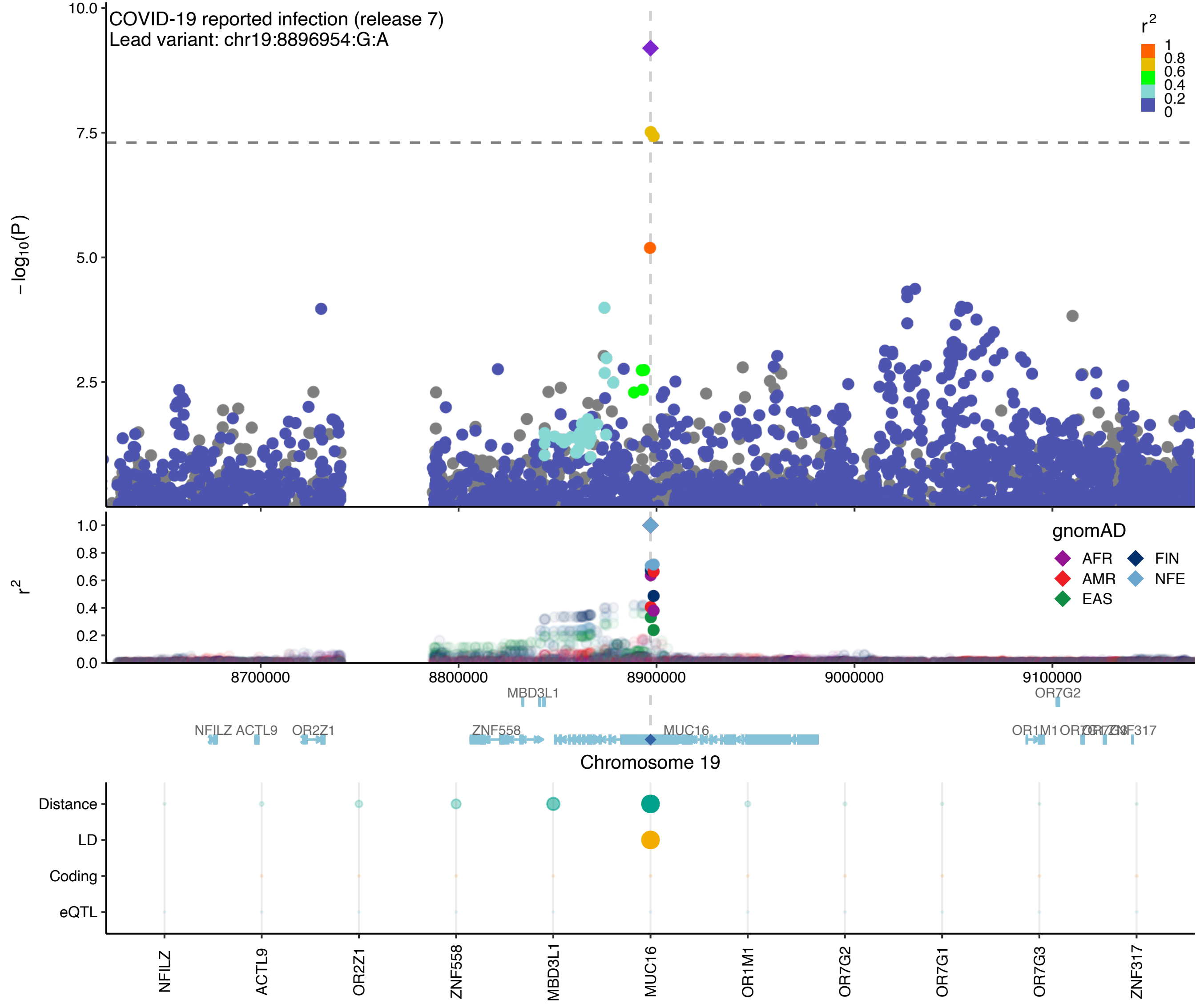

COVID-19 reported infection (release 7)
Lead variant: chr19:8896954:G:A
r2
1
0.8
0.6
0.4
0.2
0
10.0
7.5
5.0
2.5
−log10(P)
1.0
0.8
0.6
0.4
0.2
0.0
r2
gnomAD
AFR
FIN
AMR
NFE
EAS
8700000
8800000
8900000
9000000
9100000
MBD3L1
OR7G2
NFILZ
ACTL9
OR2Z1
ZNF558
MUC16
OR1M1
ZNF317
Chromosome 19
Distance
LD
Coding
eQTL
NFILZ
ACTL9
OR2Z1
ZNF558
MBD3L1
MUC16
OR1M1
OR7G2
OR7G1
OR7G3
ZNF317

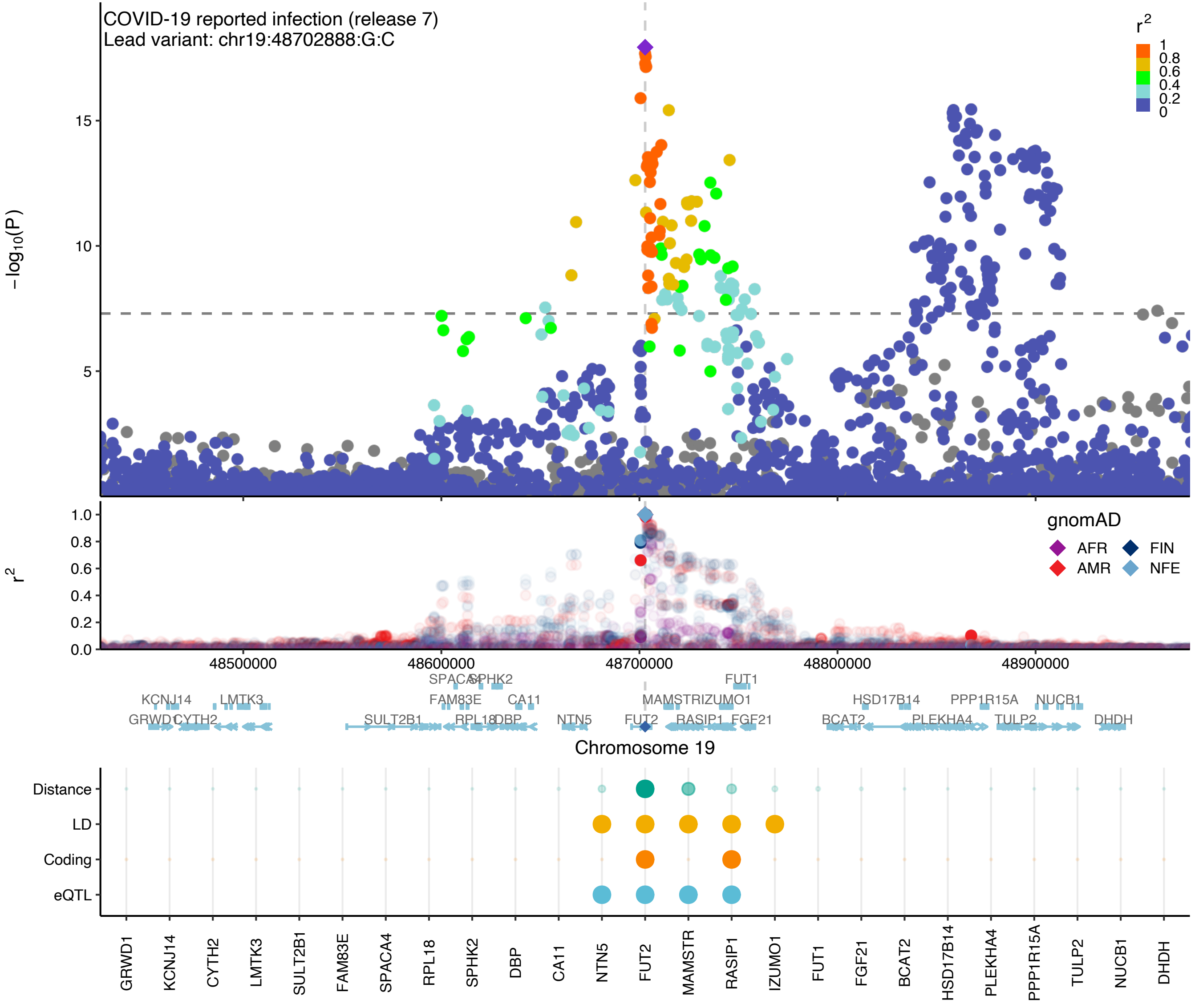

COVID-19 reported infection (release 7)
Lead variant: chr19:48702888:G:C
−log10(P)
r2
gnomAD
AFR
AMR
FIN
NFE
48500000
48600000
48700000
48800000
48900000
Chromosome 19
Distance
LD
Coding
eQTL
GRWD1
KCNJ14
CYTH2
LMTK3
SULT2B1
FAM83E
SPACA4
RPL18
SPHK2
DBP
CA11
NTN5
FUT2
MAMSTR
RASIP1
IZUMO1
FUT1
FGF21
BCAT2
HSD17B14
PLEKHA4
PPP1R15A
TULP2
NUCB1
DHDH

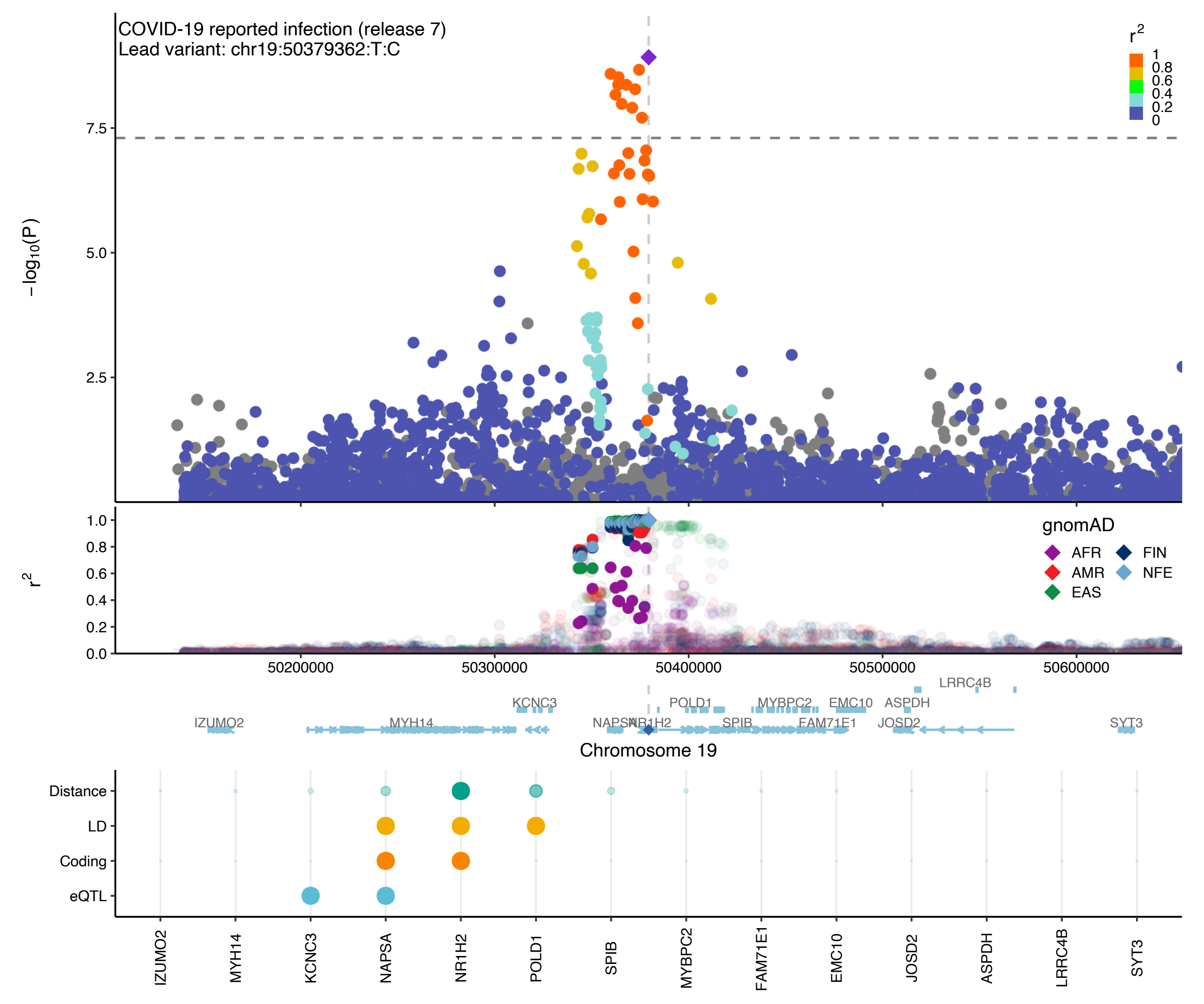
COVID-19 reported infection (release 7)
Lead variant: chr19:50379362:T:C
$r^2$
1
0.8
0.6
0.4
0.2
0
$-\log_{10}(P)$
7.5
5.0
2.5
$r^2$
1.0
0.8
0.6
0.4
0.2
0.0
gnomAD
AFR
FIN
AMR
NFE
EAS
50200000
50300000
50400000
50500000
50600000
LRRC4B
KCNC3
POLD1
MYBPC2
EMC10
ASPDH
IZUMO2
MYH14
NAPSA
NR1H2
SPIB
FAM71E1
JOSD2
SYT3
Chromosome 19
Distance
LD
Coding
eQTL
IZUMO2
MYH14
KCNC3
NAPSA
NR1H2
POLD1
SPIB
MYBPC2
FAM71E1
EMC10
JOSD2
ASPDH
LRRC4B
SYT3

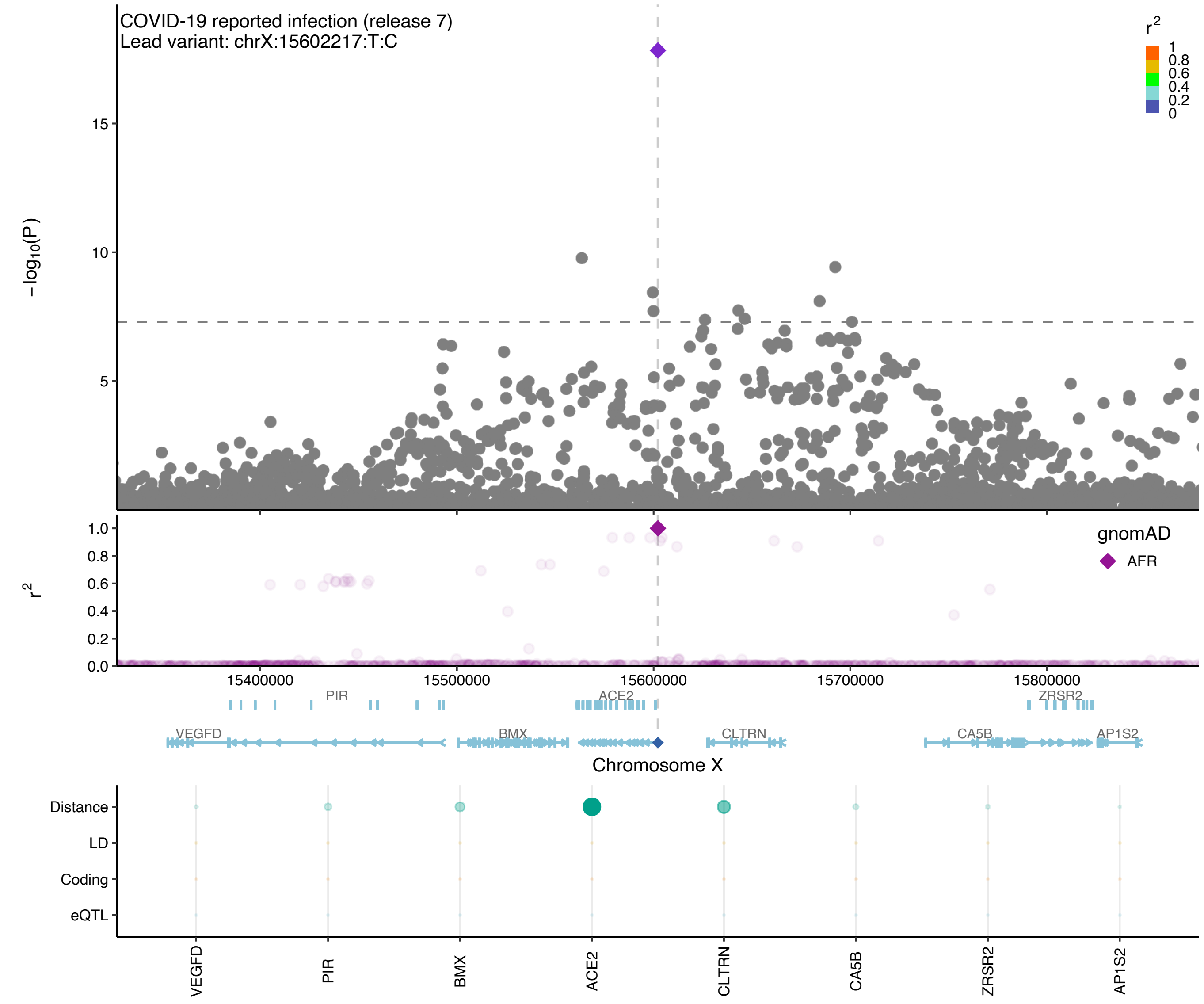

COVID-19 reported infection (release 7)
Lead variant: chrX:15602217:T:C
$-\log_{10}(P)$
$r^2$
1
0.8
0.6
0.4
0.2
0
15
10
5
1.0
0.8
0.6
0.4
0.2
0.0
gnomAD
AFR
15400000
15500000
15600000
15700000
15800000
PIR
ACE2
ZRSR2
VEGFD
BMX
CLTRN
CA5B
AP1S2
Chromosome X
Distance
LD
Coding
eQTL
VEGFD
PIR
BMX
ACE2
CLTRN
CA5B
ZRSR2
AP1S2
